# Quantitative proteomic analysis reveals different functional subtypes among IDH-wildtype glioblastoma

**DOI:** 10.1101/2025.03.09.25323141

**Authors:** Michèle Amer Salem, Jean-Louis Boulay, Marie-Françoise Ritz, Florian Samuel Halbeisen, Viviane J. Tschan, Alexander Schmidt, Katarzyna Buczak, Gregor Hutter, Severina Leu

**Author notes:** These authors contributed equally (Michèle Amer Salem, Jean-Louis Boulay, Marie-Françoise Ritz). **Corresponding author**: Severina Leu, Department of Neurosurgery, University Hospital Basel, University of Basel, Spitalstrasse 21, 4031 Basel, Switzerland.

## Abstract

**Background:** Proteomics of glioma have not yet provided biomarkers and pathways that would clearly discriminate glioma subgroups.

**Methods:** 82 glioma biopsies were prospectively collected and classified into six subgroups defined by methylomic classification: two *Isocitrate dehydrogenase (IDH)-*mutated low-grade (LGG) and four high-grade glioma (HGG) subgroups: *IDH-*mutated HGG (*IDH* HGG), proneural (GB PN), classical (GB CL), and mesenchymal glioblastoma (GB MES). Proteins were extracted and processed for liquid chromatography mass spectrometry (LC-MS). Differentially expressed proteins (DEPs) between subgroups were annotated using Gene Ontology (GO) function analysis, Kyoto Encyclopedia of Genes and Genomes (KEGG) signaling pathway analysis, and Protein-protein interaction (PPI). Functional validation was performed using inhibitor response assays in subtyped, non-cultured patient derived GB single cell suspensions.

**Results:** 5057 proteins were quantified for each sample. Tumor grading and *IDH* mutation status were the strongest discriminators for differential expression patterns. The *IDH*-wildtype GB subgroups showed diverse patterns of pathways and functions enriched with overexpressed DEP (compared to LGG): Translation and cell cycle/telomere regulation in GB PN (linked to cell proliferation), actin cytoskeleton, cell adhesion and apoptosis regulation in GB CL (linked to migration and invasion), and mitochondrial ATP synthesis in GB MES (linked to metabolism). The most overexpressed and underexpressed proteins were correlated with survival and mRNA expression data. *In vitro*, inhibition of those proteins led to reduced cell viability that differed among subgroups, albeit in a small patient-derived exploratory cohort.

**Conclusion:** This mainly descriptive study on proteomics in glioma provides insights into subgroup metabolism, and potential biomarkers for further experimental testing.

## Introduction

Gliomas are the most common primary intracranial tumors, with prognosis varying between 15 months in glioblastoma (GB) and up to several decades in diffuse low-grade glioma (LGG). Cure is not possible with current therapeutic options^1,2^. Regarding LGG, molecular genetic approaches have led to the identification of biomarkers, such as *Isocitrate dehydrogenase* (*IDH) 1* and *2* mutations, associated with chromosome arms 1p and 19q co-deletion, or *Tumor protein 53* (*TP53)* mutations. These markers are being used in the World Health Organisation (WHO) integrated classification of Central Nervous system (CNS) tumors together with histopathological features, since 2016^3^. This allowed refinement of diagnosis and management of diffuse gliomas.

Earlier glioma proteomic studies have led to the identification of three molecular subgroups among *IDH*-wildtype GB, in agreement with genomic and transcriptomic studies: proneural (GB PN), classical (GB CL) and mesenchymal GB (GB MES)^4–6^. DNA methylation profiling confirmed the subclassifications of *IDH-*wildtype as well as *IDH-*mutant gliomas into six major subgroups. The *IDH*-wildtype gliomas are GB that commonly carry chromosome 7 gain together with chromosome 10 loss^7^. Those have been further subclassified into three main subgroups: (1) subgroup of receptor tyrosine kinase (RTK) I or proneural, (2) subgroup of RTK II or classical, and (3) the mesenchymal subgroup^8^. Gliomas carrying *IDH* mutations can be subdivided into (4) those with additional 1p/19q co-deletion or low-grade oligodendroglioma, (5) those without 1p/19q co-deletion, *i.e.,* low-grade astrocytoma, and (6) high-grade astrocytoma^8^. Altogether, methylomic analysis leads to a revision of diagnosis of 12% of all central nervous system tumors^8^.

The understanding of the GB subgroups has been refined by Neftel *et. al.* in 2019^9^. They described four different GB cellular states (mesenchymal-like (MES-like), astrocyte-like (AC-like), oligodendrocyte-progenitor-like (OPC-like), and neural progenitor-like (NPC-like) states) that co-exist within one GB in certain combinations and proportions, and drive GB heterogeneity. Genetics and the microenvironment seem to influence the frequency of cells in each state, and *in vivo* single-cell lineage tracing supports plasticity between these four states. The *IDH-*wildtype GB subtypes reflect the highest frequency-states of each tumor and their associated genotypes: While the GB-CL and GB-MES subtypes correspond to tumors enriched for the AC-like and MES-like states, respectively, the GB-PN subtype corresponds to the combination of two distinct cellular states, OPC-like and NPC-like^9^. However, it is important to note that the GB states or subtypes are not part of the current WHO classification and do currently not possess any clinical relevance implying treatment decisions.

Despite the progress made in glioma classification, no single specific biomarker that clearly distinguishes HGG subgroups between one another has been found yet. One of the reasons that could be invoked is that most studies have been detecting nucleic acid abnormalities (*i.e.,* chromosomal or local aneuploidies, translocations, gain/loss-of-function or neomorphic mutations, epigenetic dysregulation, or inappropriate RNA expression). Indeed, while genetic and transcriptional abnormalities are predictors of molecular events taking place in tumor cells, protein analysis quantifies effective protein levels that are more likely to reflect underlying oncogenic mechanisms.

Quantitative proteomic analysis aims at the identification of the protein spectrum present in each tissue sample and can therefore provide valuable insights into potentially active proteins and pathways. Many aspects of the biochemical relevance of the glioma proteome remain to be investigated, to be further potentially integrated into the routine diagnostic work-up. To this end, we performed a large-scale quantitative proteomic analysis across cohorts of the six major subgroups of diffuse glioma from patients operated in our institution, with the aim to identify specific proteins and pathways characterizing each glioma subgroup.

## Material and Methods

### Patient identification

We screened all patients undergoing surgery on a diffuse glioma between July 2017 and February 2020 in the Department of Neurosurgery of the University Hospital Basel for eligibility for this study. Patients were included in the study after histological confirmation of the presumed diagnosis and if the molecular classifier revealed a clear affiliation to one of the beforehand defined six glioma subgroups. All patients gave written informed consent, and ethical approval was obtained from the Ethikkommission Nordwest-und Zentralschweiz (EKNZ, Basel, Switzerland; reference: EKNZ 02019-02358).

### Tumor tissue collection

Tumor tissue samples of patients undergoing surgery for a diffuse glioma at our institution were directly collected during surgery. Tissue was snap frozen with liquid nitrogen and stored unembedded at –80° until further use.

Since the purpose of our study was a relative comparison of the proteome inventory of individual glioma subgroups against each other, non-neoplastic control tissue samples were not included.

### Data collection

Demographic and clinical data such as age, sex assigned at birth, tumor location, clinical presentation, surgery details, adjuvant treatments and survival data were extracted from our glioma tumor database or retrospectively from patient records from the University Hospital Basel. Histologic and basic molecular data was obtained from neuropathology report.

### Tumor subclassification

Tumors were further subclassified using genome-wide DNA methylation profile data^8^ produced at Deutsches Krebsforschungszentrum (DKFZ), as following: ‘*IDH*-wildtype GB, subtype RTK I (proneural = GB PN)’; ‘*IDH*-wildtype GB, subtype RTK II (classical = GB CL)’; ‘*IDH*-wildtype GB, subtype mesenchymal = GB MES’; ‘*IDH*-mutant high-grade glioma = *IDH* HGG’; ‘*IDH*-mutant glioma = *IDH* LGG’; and ‘*IDH*-mutant, 1p/19q co-deleted oligodendroglioma = *IDH* CODEL’. As the main aim of this study was the identification of protein markers specific for individual HGG subgroups, the samples from the two LGG subgroups (*IDH* LGG and *IDH* CODEL) were pooled into one single group (= LGG) used as comparator.

### Tissue quality control

The quality of the tumor tissue was controlled histologically by cutting several 10 µm sections from each biopsy sample with a cryostat. Sections were stained with haematoxylin and examined under a light microscope to ensure enough vital tumor cells and to exclude samples with necrosis and bleeding. Tumor biopsies from 86 patients were useable for further proteomic analysis.

### Protein extraction and digestion

After quality control, a piece of 20mg of tissue was cut from the biopsy sample. The piece was then transferred into a microtube and grinded with a pellet pestle (Fisher Scientific, Reinach, Switzerland). Tissue was lysed in 8M Urea, 0.1M ammonium bicarbonate, phosphatase inhibitors (Sigma P5726&P0044) by sonication (Bioruptor, 10 cycles, 30 seconds on/off, Diagenode, Belgium) and proteins were digested as described previously ^10^. Shortly, proteins were reduced with 5 mM tris(2-carboxyethyl) phosphine for 60 min at 37 °C and alkylated with 10 mM chloroacetamide for 30 min at 37 °C. After diluting samples with 100 mM ammonium bicarbonate buffer to a final urea concentration of 1.6M, proteins were digested by incubation with sequencing-grade modified trypsin (1/50, w/w; Promega, Madison, Wisconsin) for 12 h at 37°C. After acidification using 5% trifluoroacetic acid (TFA), peptides were desalted using C18 reverse-phase spin columns (Macrospin, Harvard Apparatus) according to the manufacturer’s instructions, dried under vacuum and stored at –20°C until further use.

### Tandem mass tag (TMT) proteomics

Sample aliquots comprising 10 μg of peptides were labeled with isobaric tandem mass tags (TMTpro 16-plex, Thermo Fisher Scientific). To enable the normalization across multiple TMT sets, the representative sample pool was prepared and labelled in each TMT set (for details see the experimental design table (Supplementary Table 1)). Peptides were resuspended in 10 μl labeling buffer (2 M urea, 0.2 M HEPES, pH 8.3) by sonication and 2.5 μL of each TMT reagent was added to the individual peptide samples followed by a 1 h incubation at 25°C shaking at 500 rpm. To quench the labelling reaction, 2 μL aqueous 0.75 M hydroxylamine solution was added, and samples were incubated for another 5 min at 25°C shaking at 500 rpm followed by pooling of all samples. The pH of the sample pool was increased to 12 by adding 1 M phosphate buffer (pH 12) and incubated for 20 min at 25°C shaking at 500 rpm to remove TMT labels linked to peptide hydroxyl groups. Subsequently, the reaction was stopped by adding 2 M hydrochloric acid until a pH < 2 was reached. Finally, peptide samples were further acidified using 5 % TFA, desalted C18 reverse-phase spin columns (Macrospin, Harvard Apparatus) according to the manufacturer’s instructions and dried under vacuum.

### High performance liquid chromatography (HPLC) fractionation

TMT-labeled peptides were fractionated by high-pH reversed phase separation using a XBridge Peptide Ethylene Bridged Hybrid (BEH) C18 column (3,5 µm, 130 Ångstrom (Å), 1 mm x 150 mm, Waters) on an Agilent 1260 Infinity HPLC system. Peptides were loaded on column in buffer A (20 mM ammonium formate in water, pH 10) and eluted using a two-step linear gradient from 2% to 10% in 5 minutes and then to 50% buffer B (20 mM ammonium formate in 90% acetonitrile, pH 10) over 55 minutes at a flow rate of 42 µl/min. Elution of peptides was monitored with an ultraviolet (UV) detector (215 nm, 254 nm) and a total of 36 fractions were collected, pooled into 12 fractions using a post-concatenation strategy as previously described^11^ and dried under vacuum.

### Liquid chromatography (LC)-MS/MS analysis

Dried peptides were resuspended in 0.1% aqueous formic acid and subjected to LC–MS/MS analysis using a Q Exactive HF Mass Spectrometer fitted with an EASY-nLC 1000 (both Thermo Fisher Scientific) and a custom-made column heater set to 60°C. Peptides were resolved using a RP-HPLC column (75μm × 30cm) packed in-house with C18 resin (ReproSil-Pur C18–AQ, 1.9 μm resin; Dr. Maisch GmbH) at a flow rate of 0.2 μLmin^-1^. The following gradient was used for peptide separation: from 5% B to 15% B over 10 min to 30% B over 60 min to 45 % B over 20 min to 95% B over 2 min followed by 18 min at 95% B. Buffer A was 0.1% formic acid in water and buffer B was 80% acetonitrile, 0.1% formic acid in water.

The mass spectrometer was operated in DDA mode with a total cycle time of approximately 1 s. Each MS1 scan was followed by high-collision-dissociation of the 10 most abundant precursor ions with dynamic exclusion set to 30 seconds. For MS1, 3e6 ions were accumulated in the Orbitrap over a maximum time of 100 ms and scanned at a resolution of 120,000 FWHM (at 200 m/z). MS2 scans were acquired at a target setting of 1e5 ions, maximum accumulation time of 100 ms and a resolution of 30,000 FWHM (at 200 m/z). Singly charged ions and ions with unassigned charge state were excluded from triggering MS2 events. The normalized collision energy was set to 30%, the mass isolation window was set to 1.1 m/z and one microscan was acquired for each spectrum.

The acquired raw files were analysed using the SpectroMine software (Biognosis AG, Schlieren, Switzerland).

Spectra were searched against a human database consisting of 20742 protein sequences (downloaded from Uniprot on 20190307^12^) and 392 commonly observed contaminants. The search criteria were set as follows: full tryptic specificity was required; 2 missed cleavages were allowed; carbamidomethylation (C) and TMT16plex (K, N-term) were set as fixed modifications; oxidation (M) and acetyl (protein N-term) were set variable modification. Peptide Spectrum Match (PSM) FDR cut-off was set to 0.01. Raw reporter ions intensities of protein group specific PSMs were exported and used for quantification.

For each TMTpro 16-plex experiment, raw PSMs intensities were summed within a protein group and normalized using quantiles method^13^. The normalization between multiple TMT experiments was performed as described here^14^. In brief, for each protein, the geometric means of reference channel intensities were calculated. For each TMTpro 16-plex, the protein specific scaling was then calculated as ratio of geometric mean to the reference channel intensity. Protein intensities were then multiplied by the calculated scaling factors.

## Quantification and statistical analysis

### Statistical analysis

We performed descriptive data analyses to summarize baseline patient characteristics. Qualitative data were summarized with the number of observations and percentages. Quantitative data were reported as median and range. To assess difference between tumor subgroups, Fisher exact tests for categorical data, and Kruskal-Wallis rank sum tests for quantitative data were used. All statistical analyses were performed using R^15^.

### Identification of differentially expressed proteins (DEPs)

For the analysis of the proteomics data, we only included protein samples where we had complete data. We transformed the absolute protein abundances to log_2_ values and fitted protein-wise linear models to the expression data using the Limma R library. We performed the differential expression analysis to identify DEPs between subgroups and between diffuse LGG and *IDH* HGG, GB PN, GB CL, and GB MES subgroups.

Age, sex, treatments prior to surgery where the tumor sample was taken (chemotherapy, radiotherapy, steroids), and epilepsy were included as confounding factors into the model. As our aim of the study was the identification of ideally subgroup-specific biomarkers and gliomas are known to be contaminated with non-neoplastic cells^16^, we introduced three proteins as potential confounders into the multivariable model to exclude the DEPs significantly associated with these markers: HLA class II histocompatibility antigen gamma chain (HG2A) and Receptor-type tyrosine-protein phosphatase C (PTPRC, CD45) as marker for immune cells^17^, and Neuronal-specific septin-3 (SEPT3) as marker for neurons^18^.

We used the Benjamini-Hochberg’s method to control for the false discovery rate in multiple testing. The threshold of statistical significance was set at adjusted p < 0.05. We then used heatmaps, barplots, and Venn diagrams to visualize the DEPs between our contrast groups. The heatmap displays the expression levels of each DEP, showing the variation in protein expression between the groups. Barplots are used to illustrate the number of overexpressed and underexpressed DEPs. Venn diagrams show the unique and shared DEPs between the contrast groups, showing the overlap in protein expression.

### Dimensionality Reduction: Principal Component Analysis (PCA) and Uniform Manifold Approximation and Projection (UMAP)

We used PCA and UMAP to our dataset of DEPs to both simplify its complexity and visualize the underlying patterns among the DEPs across various glioma subgroup contrasts. PCA effectively captured the main variations among DEPs, while UMAP provided a nuanced mapping of protein expression patterns. These analyses were performed using the FactoMineR and umap packages in R, respectively^15^.

### Gene Ontology (GO) enrichment, Kyoto Encyclopedia of Genes and Genomes (KEGG) pathway analysis and Protein-protein interaction (PPI) using Over-representation analysis (ORA)

We used ORA to perform independent assessments of GO enrichment, KEGG Pathway analysis, and PPI of all DEPs between the glioma subgroups. For the annotation of the GO terms, we used the Uniprot accession numbers^12^ of the Bioconductor annotation data package org.Hs.eg.db. For the annotation of KEGG data, we used the directly download from the KEGG website (https://www.kegg.jp/). For these analyses, we utilized the clusterProfiler package in R^15^, and the String database enrichment tool (Version 12.0)^19^.

### GO enrichment and KEGG Pathway analysis using Gene Set Enrichment Analysis (GSEA)

We used GSEA to perform independent assessments of both GO enrichment and KEGG Pathway analysis. In our GSEA approach, we utilized two distinct outcomes: the logarithmic fold change (log_2_FC) and the signed p-value (which was determined based on the direction of the log_2_FC). To ensure the robustness of our findings, we only considered those terms that were significantly enriched based on both the log_2_FC and the signed p-value outcomes.

Enriched terms were identified by using the highest and lowest Normalized Enrichment Score values as they contain the highest amounts of over– and underexpressed proteins, respectively.

For the annotation of the GO terms, we used the Uniprot accession numbers^12^ of the Bioconductor annotation data package org.Hs.eg.db. For the annotation of KEGG data, we used the directly download from the KEGG website (https://www.kegg.jp/). For these analyses, we utilized the clusterProfiler package in R.

### Correlation of DEPs with survival of our cohort

To clinically validate our data, we performed survival analysis to investigate the relationship between the three most overexpressed and the three most underexpressed differentially expressed proteins (DEPs) and survival. We first split the DEP abundance into high and low expression groups at the median. Using the dichotomized data, we applied Kaplan-Meier survival analysis to estimate survival curves and used the log-rank test to assess differences between these groups.

Further, we performed univariable Cox proportional hazards regression using the continuous DEP abundance to quantify the association between protein expression levels and survival outcomes. For this analysis, we reported hazard ratios (HR) along with 95% confidence intervals (CI) to estimate the risk associated with each unit increase in protein abundance.

### Correlation of the DEPs with mRNA expression and survival data from The Cancer Genome Atlas (TCGA)

To externally validate our proteomics data, we compared them with the publicly available TCGA^20^ glioma mRNA expression and survival data from a composited cohort consisting of 152 HGG and 515 LGG patients (TCGA GBMLGG) available on the GlioVis platform^21^.

## Functional validation using inhibitor response assays

### Dissociation of GB tumors

Freshly resected tumor pieces were manually minced using razor blades and enzymatically dissociated at 37 °C for 30Lmin with 1 mg/mL collagenase type IV (Worthington Biochemical Corporation, USA) and 250 μ/mL DNase 1 (Roche, Switzerland) in a dissociation buffer containing HBSS (with Ca^2+^/Mg^2+^), 1% non-essential amino acids, 1 mM sodium pyruvate, 44 mM sodium bicarbonate, 25 mM HEPES, 1% GlutaMAX-I and 1% antibiotic-antimycotic (all from Gibco, USA). The resulting cell suspensions were filtered through a 70LμM strainer and centrifuged in a 0.9 M sucrose gradient. Erythrocytes were removed using ACK lysing solution (ThermoFisher Scientific, USA), and cell suspensions were frozen in Bambanker^TM^ Standard until further use.

### Inhibitor response assays

To perform a functional validation of the identified targets, cells were plated at a concentration of between 3500 and 15000 cells per well in low-adherence 384 well plates. 24 h after plating, 7 inhibitors targeting the identified DEPs were selected (Supplementary Table 2) and an 8 to 9-point dilution series of each compound was dispensed in four replicates using a Tecan Digital Dispenser D300e (Tecan). Drug concentrations spanned from 50 pM to 500 μM, depending on the drug. Cell viability was measured by CellTiter-Glo 3D assay following 3 days of drug incubation, and results were normalized to untreated controls. Data analyses were performed using GraphPad Prism, applying nonlinear regression (curve fit) and the equation [inhibitor] vs. normalized response (variable slope).

To assess differences in cell viability across different patients for each compound and concentration, the Kruskal-Wallis rank sum test was used as a global test for differences, followed by Dunn’s post-hoc test with Bonferroni correction for pairwise comparisons to assess differences in cell viability across different patients for each compound and concentration. The threshold of statistical significance was set at p < 0.05. All statistical analyses were performed using R.

### Reference data

Some reference data shown in this study are in whole or part based upon data generated by the TCGA Research Network (https://www.cancer.gov/tcga)^20^, by the STRING consortium 2023 (https://string-db.org/)^19^, by Uniprot (https://www.uniprot.org)^12^, and by GlioVis (http://gliovis.bioinfo.cnio.es/)^21^.

## Results

### Features of the glioma subgroups

A total of 122 glioma patients undergoing surgery on a diffuse glioma between July 2017 and February 2020 at the Department of Neurosurgery of the University Hospital Basel were initially screened for eligibility for this study. After confirmation of histologic and molecular diagnosis, subgroup assignment was defined by DNA methylation-based classification^8^. We performed proteomic analysis on tumors from 86 patients. After exclusion of 4 patients (GB CL_061, GB MES_072, and GBMES_073, IDHLGG_027) due to poor or missing proteomic data, 82 patients remained for final analysis.

Mean age at diagnosis was significantly lower in the *IDH* mutants compared to the *IDH*-wildtype subgroups (p < 0.001). Patients from all subgroups had similar mean Karnofsky index (p = 0.642) and sex ratio (p = 0.692). Patients in the LGG subgroup had epilepsy in 85.3% of cases compared to lower values in the HGG subgroups (range from 50 to 69.2%), even though this difference did not reach statistical significance (p = 0.139) (Baseline characteristics, Table 1). Patients with LGG had a significantly longer median survival than patients with HGG. *IDH* HGG did show a significantly longer median survival compared to the other HGG subgroups, whereas there was no difference between the median survival of the three *IDH*-wildtype GB subgroups (Figure 1).

**Figure 1:**
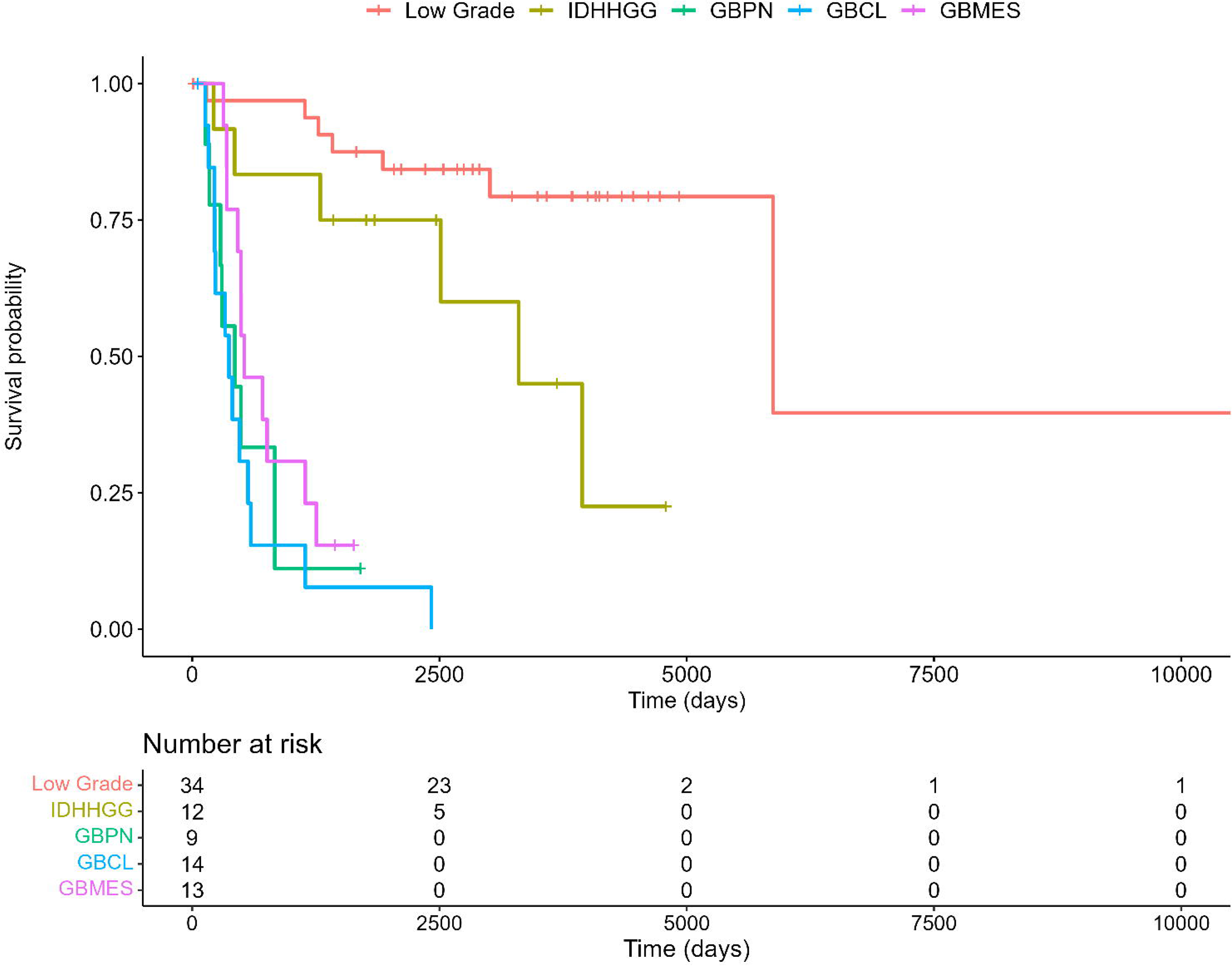
Kaplan-Meier curve showing the overall survival of the 6 glioma subgroups (Note: the two LGG subgroups are given as one pooled group (Low grade)).

**Table 1:** Baseline characteristics of the glioma subgroups (Note: the two low grade glioma subgroups are pooled as one group (diffuse LGG))

| Baseline Characteristics | Diffuse LGG<br>(N = 34) | IDH HGG<br>(N = 12) | GB PN<br>(N = 9) | GB CL<br>(N = 14) | GB MES<br>(N = 13) | P-value |
| --- | --- | --- | --- | --- | --- | --- |
| Male sex (%) | 24/34 (70.6%) | 6/12 (50%) | 7/9 (77.8%) | 9/14<br>(64.3%) | 8/13<br>(61.5%) | 0.692<sub>a</sub> |
| Epilepsy (%) | 29/34 (85.3%) | 6/12 (50%) | 6/9 (66.7%) | 9/14<br>(64.3%) | 9/13<br>(69.2%) | 0.139<sub>a</sub> |
| Median age (range) | 44.5 (20 to 79) | 38 (27 to 73) | 60 (55 to 78) | 66 (48 to<br>76) | 59 (52 to<br>79) | <0.001<sub>b</sub> |
| Mean Karnofsky index<br>(range) | 80 (70 to 100) | 80 (70 to 100) | 80 (70 to 90) | 80 (60 to<br>90) | 80 (70 to<br>90) | 0.642<sub>b</sub> |
| Surgery type<br>(GTR: Gross<br>total resection, STR:<br>subtotal resection) | 21 GTR | 5 GTR | 5 GTR | 6 GTR | 5 GTR | 0.496<sub>a</sub> |
|  | 13 STR | 7 STR | 4 STR | 8 STR | 7 STR |  |
|  |  |  |  |  | 1 biopsy |  |
| Tumor side | 16 right sided | 5 right sided | 5 right sided | 11 right<br>sided | 4 right<br>sided | 0.379<sub>a</sub> |
|  | 15 left sided | 6 left sided | 4 left sided | 3 left sided | 7 left sided |  |
|  | 3 bilateral | 1 bilateral |  |  | 2 bilateral |  |
| Most common affected<br>lobe | 23/34 frontal | 8/12 frontal | 6/9 temporal | 7/14<br>parietal | 7/13<br>temporal | n/a |

<sub>a</sub> Fisher's Exact Test
<sub>b</sub> Kruskal-Wallis Rank Sum Test

The *IDH* mutated tumors were most commonly located in the frontal lobe, whereas GB PN and GB MES were most commonly found in the temporal, and GB CL in the parietal lobe. 21/34 LGG (61.7%), 5/12 *IDH* HGG (41.7%), 5/9 GB PN (55.6%), 6/14 GB CL (42.9%), and 5/13 GB MES (38.5%) underwent gross total resection. 2/82 patients had chemotherapy and only 1/82 patients had radiotherapy prior to the surgery, at the time the sample for the proteomic analysis has been taken. 46/82 patients had been given steroids prior to surgery. As expected, this rate was higher in the HGG (35/48 patients; 72.9%) than in the LGG (11/34 patients; 32.4%) (Supplementary Table 3. Neither treatment prior to surgery (chemotherapy, radiotherapy, steroids) nor confounding factors (age, epilepsy) was significantly associated with any of the DEPs, although sex impacted the expression of Y chromosome-encoded Adenosine triphosphate (ATP)-dependent RNA helicase DDX3Y (DDX3Y).

Additional collected data comprised local genetic imbalances such as *Epidermal growth factor receptor (EGFR)* amplification (in addition to chromosome 7 triploidy) and *Cyclin-dependent kinase inhibitor 2A* (*CDKN2A)* homozygous deletion statuses; *Telomerase Reverse Transcriptase* (*TERT)* promoter mutations and TP53 immunopositivity (as an indicator for possible *TP53* mutation when higher than 20%^22^), and hemizygous co-deletions at 1p and 19q chromosome arms) as well as methylomic data. Taken together, these data converged to consistent definitions of the subgroups of our cohort (Supplementary Table 1).

By introduction of HG2A, PTPRC/CD45 (reflecting immune cell contents in tumors), and SEPT3 (neuron contents) as non-neoplastic cell markers into the multivariate model, 464 proteins that were significantly different between at least two subgroups before were removed from, and 137 proteins were added to the total amount of DEPs between the subgroups.

### Proteomic profiles primarily segregate LGG from HGG, and *IDH-*mutant from *IDH*-wildtype tumors

After exclusion of samples with missing protein data, 5057 common proteins were identified and quantified in each tumor sample. Profiles of DEPs distinguished the *IDH*-mutated subgroups (LGG and *IDH* HGG) from the three *IDH*-wildtype GB subgroups (Figure 2a), while PCA analysis and UMAP both showed a main separation between LGG and HGG subgroups, followed by *IDH* mutation status (Figure 2b and 2c). The largest differences in DEP numbers were observed between GB CL and LGG (162 DEPs), followed by GB PN and LGG (144 DEPs), GB MES and LGG (28 DEPs), and lastly *IDH* HGG and LGG (1 DEP) (Figure 2d). We did not observe DEPs that were significantly overexpressed in all four HGG subgroups (Figure 2e).

**Figure 2:**
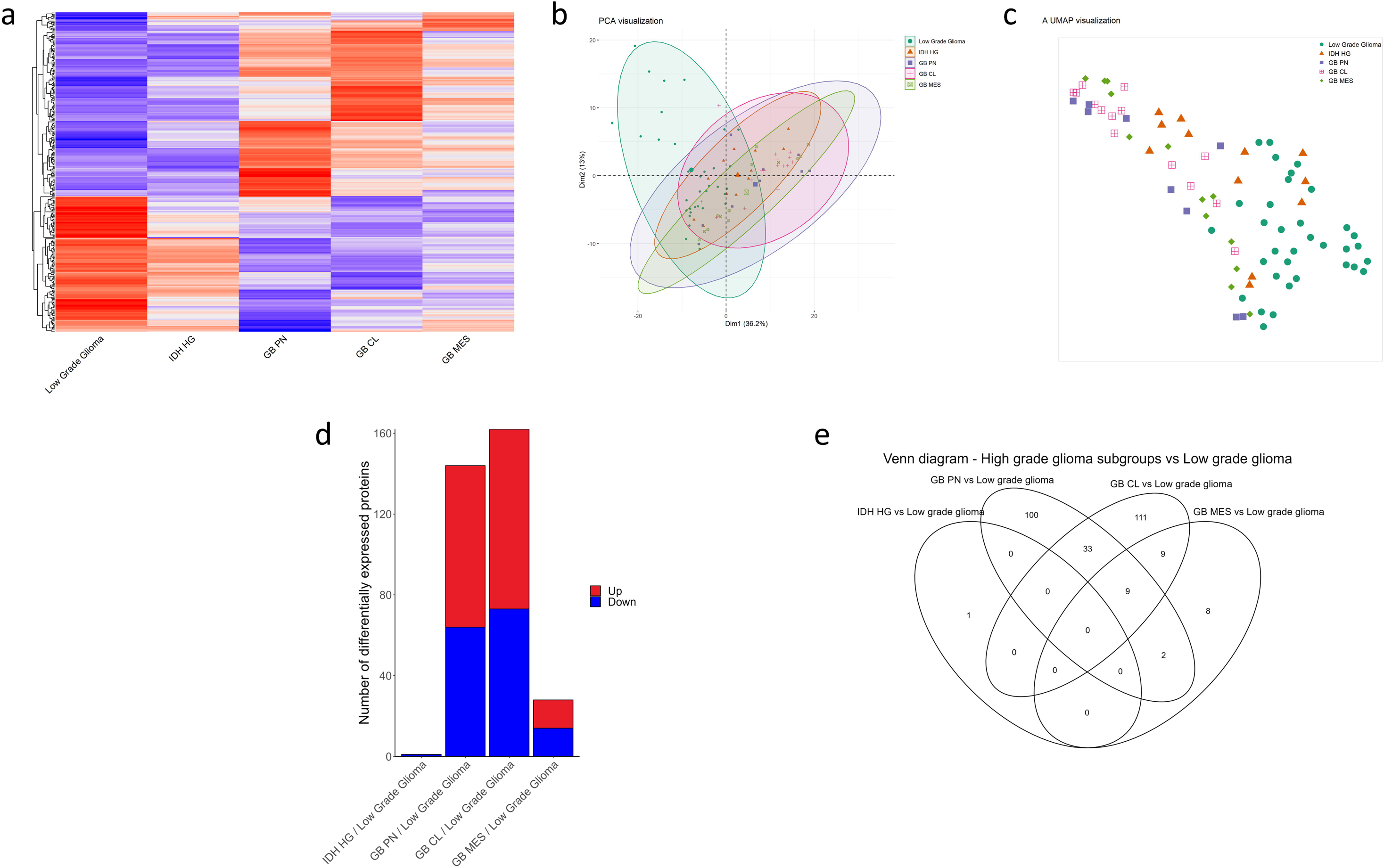
Proteomic profiles segregate LGG from HGG and *IDH-*mutant from *IDH-*wildtype tumors. a) Heatmap of abundance values of DEPs between LGG and HGG subgroups. Overexpressed proteins in red, underexpressed proteins in blue (this applies also to B). b) PCA analysis showing clear separation and some overlap between the LGG and all four HGG subgroups. c) UMAP showing the clustering of the DEPs of the LGG and all four HGG subgroups. d) Number of DEPs if comparing the four HGG subgroups to the pooled LGG subgroups. e) Venn diagram showing the DEPs that are common between the LGG and each HGG subgroups. There was no common protein significantly differentially expressed between the LGG and all four HGG subgroups.

### GB PN did show enrichment in functions important for cell proliferation

Among the 144 DEPs between GB PN and pooled LGG (Figure 3a), 80 proteins were overexpressed in GB PN, and 64 underexpressed in GB PN (compared to LGG) (Supplementary Tables 4a and 4b).

**Figure 3:**
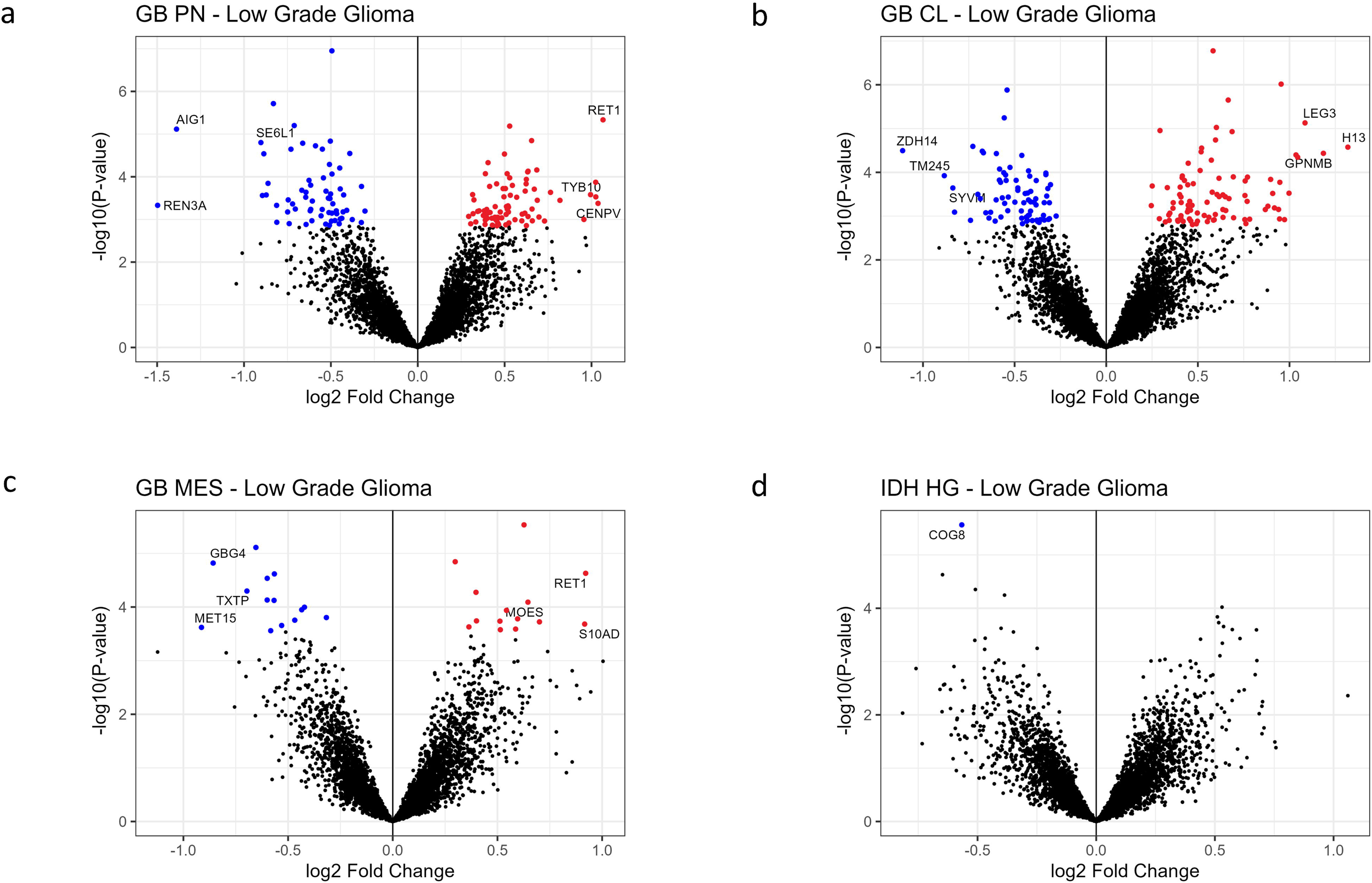
Volcano plots of DEPs between each HGG subgroup and LGG. a) Volcano plot of 144 DEPs between the GB PN and LGG; Red and blue dots indicate significantly over– and underexpressed proteins, respectively (this applies also to Figures 3b to 3d). b) Volcano plot of 162 DEPs between the GB CL and LGG. c) Volcano plot of 28 DEPs between the GB MES and LGG. d) Volcano plot of 1 DEPs between the *IDH* HGG and LGG.

RalA-binding protein 1 (RBP1), Centromere protein V (CENPV), and Thymosin Beta 10 (TYB10) were the proteins showing the highest log_2_FC values in GB PN (compared to LGG). Translation and cell cycle/telomere regulation were dominantly associated with overexpressed DEPs in GB PN, and the most enriched GO cellular compartment was the ribosome (Table 2, Figure 4a, Supplementary Table 5).

**Figure 4:**
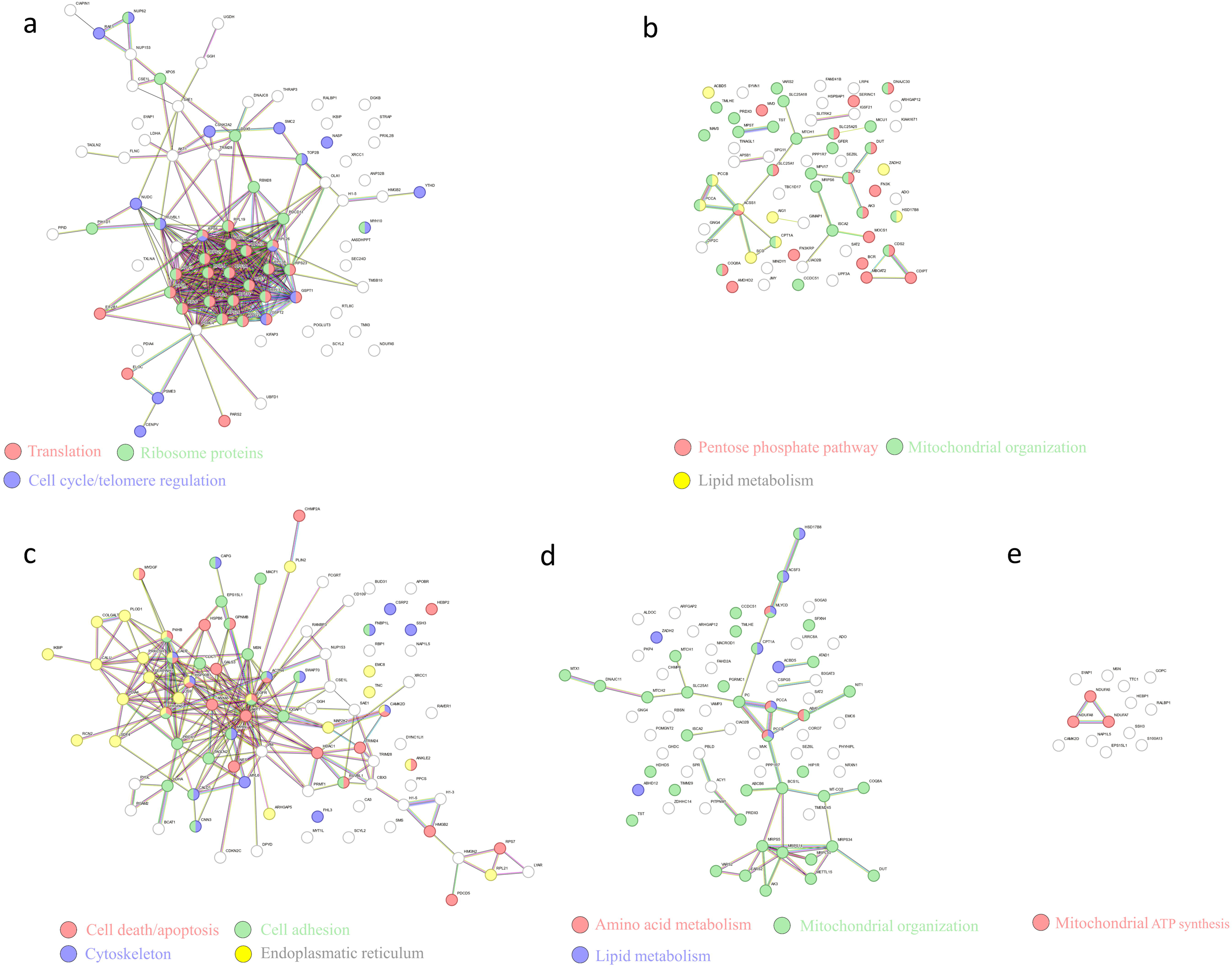
ORA (over-representation analysis) of the DEPs over– and underexpressed in the IDH-wildtype GB subgroups (compared to LGG) using the String functional enrichment tool (KEGG, GO, and PPI) a) Translation (network nodes in red), ribosome proteins (green) and cell cycle/telomere regulation (blue) were dominantly increased in the 80 DEPs overexpressed in GB PN. The most enriched GO cellular compartment was the ribosome. b) Functions involved in mitochondrial organization (network nodes in green), lipid metabolism (yellow), and pentose phosphate pathway (red) were enriched in the 64 DEPs underexpressed in GB PN. The most enriched GO cellular compartment was the mitochondrion (green). c) Functional enrichment was seen in cell adhesion (network nodes in green), actin cytoskeleton (blue) and cell death/apoptosis (red) among the 89 DEPs overexpressed in GB CL. The most enriched GO cellular compartment was the endoplasmic reticulum (yellow). d) Functional enrichment was seen in mitochondrial organization (network nodes in green), lipid metabolism (blue), and amino acid metabolism (red) among the 73 proteins underexpressed in GB CL. The most enriched GO cellular compartment was the mitochondrion (green). e) Functional enrichment was seen in mitochondrial ATP synthesis (red) among the 14 overexpressed DEPs in GB MES. Note: Results for the 14 DEPs underexpressed in GB MES are not shown, as these were only seen in GSEA (and not in ORA).

**Table 2:** Functional enrichment detected using the String functional enrichment tool (https://www.string-db.org)^19^ including GO and KEGG annotation among the over– and underexpressed proteins in the HGG subgroups (compared to LGG). (Note: if not otherwise specified, enrichment was found in Over-representation analysis (ORA) and Gene Set Enrichment Analysis (GSEA) analysis)

| HGG subgroup<br>(compared to LGG) | Pathways and functions containing over- and underexpressed DEP, respectively: |  |
| --- | --- | --- |
| **GB PN** | Containing overexpressed DEP | Translation (ORA and GSEA)<br>Cell cycle/telomere regulation (only GSEA) |
|  | Containing underexpressed DEP | Mitochondrial organisation<br>Lipid metabolism<br>Pentose phosphate pathway |
| **GB CL** | Containing overexpressed DEP | Cell adhesion<br>Actin cytoskeleton<br>Cell death/apoptosis regulation |
|  | Containing underexpressed DEP | Mitochondrial organisation<br>Lipid metabolism<br>Protein metabolism |
| **GB MES** | Containing overexpressed DEP | Mitochondrial ATP synthesis (ORA and GSEA)<br>Actin cytoskeleton (only GSEA) |
|  | Containing underexpressed DEP | Translation (only GSEA)<br>Lipid metabolism (only GSEA) |
| **IDH HGG** | Containing overexpressed DEP | Mitochondrial ATP synthesis (only GSEA) |
|  | Containing underexpressed DEP | Golgi organization (ORA and GSEA)<br>Translation (only GSEA) |

Regulator of nonsense transcripts 3A (REN3A), Androgen-induced gene 1 protein (AIG1), and Seizure 6-like protein (SE6L1) showed the highest log_2_FC values in LGG (compared to GB PN). Functions in mitochondrial organization, lipid metabolism and pentose phosphate pathway appeared enriched with underexpressed DEPs, and the mitochondrion was the most enriched GO cellular compartment among the underexpressed DEPs in GB PN (compared to LGG) (Table 2, Figure 4b, Supplementary Table 5).

### Overexpressed DEPs in GB CL exhibited functions required for cell migration and invasion

Among the 162 DEPs between GB CL and pooled LGG (Figure 3b), 89 proteins were overexpressed in GB CL, and 73 underexpressed in GB CL (compared to LGG) (Supplementary Tables 6a and 6b).

Histone H1.3 (H13), Glycoprotein NMB (GPNMB), and Galectin-3 (LEG3) were the proteins that showed the highest log_2_FC values in GB CL (compared to LGG). In GB CL, functional enrichment was seen in cell adhesion, actin cytoskeleton and cell death/apoptosis, The most enriched GO cellular compartment was the endoplasmatic reticulum (Table 2, Figure 4c, Supplementary Table 5).

Palmitoyltransferase ZDHHC14 (ZDH14), Transmembrane protein 245 (TM245), and Valine-tRNA ligase, mitochondrial (SYVM) showed the highest log_2_FC values in LGG (compared to GB CL). DEPs underexpressed in GB CL did show functional enrichment in mitochondrial organization and lipid as well as protein metabolism. The mitochondrion was the most common GO cellular compartment among the underexpressed DEPs in GB CL (Table 2, Figure 4d, Supplementary Table 5).

### GB MES displayed sigs of activated metabolism

Among the 28 DEPs between GB MES and pooled LGG (Figure 3c), 14 proteins were overexpressed in GB MES, and 14 underexpressed in GB MES (compared to LGG) (Supplementary Tables 7a and 7b).

RBP1, Protein S100-A13 (S10AD), and Moesin (MOES) were the proteins that showed the highest log_2_FC values in GB MES (compared to LGG). Overexpressed DEPs in GB MES showed functional enrichment in mitochondrial ATP synthesis and actin cytoskeleton (Table 2, Figure 4e, Supplementary Table 5).

12S rRNA N4-methylcytidine methyltransferase (MET15), Guanine nucleotide-binding protein G(I)/G(S)/G(O) subunit gamma-4 (GBG4), and Tricarboxylate transport protein, mitochondrial (TXTP) did show the highest log_2_FC values in LGG (compared to GB MES). Underexpressed DEPs in GB MES (compared to LGG) did show functional enrichment in translation and lipid metabolism (Table 2).

### *IDH* HGG and LGG were characterized by similar proteomic features

Only one protein (Conserved oligomeric Golgi complex subunit 8 (COG8)) was significantly overexpressed in pooled LGG compared to *IDH* HGG with a log_2_FC of 0.56 (Figure 3d, Supplementary Tables 8a and 8b).

Mitochondrial ATP synthesis was associated with overexpressed DEPs in *IDH* HGG in the GSEA, while the underexpressed DEPs in *IDH* HGG were associated with Golgi organization (Table 2, Supplementary Table 5).

### Different protein expression between HGG subgroups

Individual comparisons revealed 22 DEPs between GB CL and *IDH* HGG (Supplementary Table 9a), whereas there were no DEPs between the other individual HGG subgroups. Ras GTPase-activating-like protein IQGAP1 (IQGA1) was the sole overexpressed protein in GB CL compared to the other pooled HGG subgroups (Supplementary Table 9b), a protein that plays a crucial role in regulating the dynamics and assembly of the actin cytoskeleton.

### Correlation of the most over– and underexpressed DEPs with overall survival data from our cohort

We analyzed the three most overexpressed and three most underexpressed DEPs from each subgroup in relation to overall survival within our cohort. High expression of TYB10, H13, GPNMB, LEG3, S10AD, and MOES was significantly associated with shorter overall survival, whereas high expression of SE6L1, AIG1, REN3A, SYVM, TM245, TXTP, GBG4 and MET15 was significantly associated with longer overall survival (Supplementary Figure 1). However, two of the most underexpressed proteins (ZDH14 (p=0.18), and COG8 (p=0.74)) and two of the strongest overexpressed proteins (RBP1 (p=0.89) and CENPV (p=0.86)) did not show a statistically significant association with survival (Supplementary Figure 1, Supplementary Table 10).

### Correlation of our results with transcriptomic and survival data from TCGA

We compared our proteomics data with the publicly available TCGA^20^ glioma mRNA expression and survival data from a composited cohort consisting of 152 HGG and 515 LGG patients (TCGA GBMLGG) available on the GlioVis platform^21^. Most of the proteins significantly overexpressed in our HGG subgroups showed higher mRNA levels in GB compared to LGG, and vice versa for the proteins that were underexpressed in the HGG subgroups (Supplementary Figures 2 – 5). Higher mRNA expression levels of all the proteins overexpressed in our HGG subgroups were associated with worse survival (compared to lower mRNA expression levels) (Supplementary Figures 2 – 4). However, the TCGA survival data was less consistent for the mRNA expression coding for proteins that were significantly underexpressed in our HGG subgroups (Supplementary Figures 2 – 5).

### Functional validation with inhibition of the most overexpressed DEPs led to reduced cell viability at higher concentrations that was different between *IDH*-wildtype GB subgroups

To assess the functional relevance of the DEPs in each subgroup and their potential subgroup-specificity, we tested the efficacy of protein inhibitors in a viability assay, using patient-derived, non-cultured, subtyped^8^ GB single cell suspensions with a sample from each subgroup and one sample defined as a “mixed” sample expressing GB PN and CL characteristics. Inhibition of those most overexpressed DEPs reduced the cell viability, partly depending on GB subgroup.

Inhibition of LEG3 (overexpressed in GB CL) using GB1107^23^ resulted in reduced cell viability in samples of all subgroups, most pronounced in the GB CL sample (significant difference to the GB MES sample at 500 μM). Thus, overexpression of LEG3 in GB CL might lead to higher susceptibility against inhibition (Supplementary Figure 6).

Inhibition of GPNMB (overexpressed in GB CL) using the antibody drug conjugate Glembatumumab Vedotin^24,25^ resulted in only a moderate cell viability reduction across all samples, even at the highest concentrations. However, it impacted the mixed sample at higher concentrations significantly compared to the GB PN sample (Supplementary Figure 7).

Increasing concentrations of the histone deacetylase inhibitor Vorinostat^26^ (inhibiting H13 that is overexpressed in GB CL) led to stepwise reduced cell viability in samples of all subgroups. The GB CL sample preserved the highest levels of cell viability up to a dose range of 100 μM, but the GB MES sample outperformed the GB CL sample at 500 μM. The cell viability of the GB PN sample was stable at lower concentrations but decreased rapidly with higher concentrations. Hence, overexpression of H13 might lead to more robustness against inhibition (Supplementary Figure 8).

Inhibition of CENPV (overexpressed in GB PN) using GSK-923295^27,28^ reduced cell viability sharply in samples of all subgroups at dose ranges between 50 to 100 μM. At 50 μM, significant differences between the mixed sample and the GB CL and GB PN samples could be observed. However, GSK-923295 is specifically inhibiting CENPE, and thus the effect on CENPV could be unspecific (Supplementary Figure 9).

Inhibition of S10AD (overexpressed in GB MES) using Amlexanox^29^ reduced cell viability in samples of all subgroups at higher concentrations and most in the GB MES sample, even though without statistically significant difference at 500 μM (Supplementary Figure 10).

Inhibition of MOES (overexpressed in GB MES) was performed using NSC668394^30^ and Fasudil HCl^31^ (indirect inhibitor). Inhibition of MOES with NSC668394 led to a sudden reduction in cell viability at concentration ranges of 50 and 100 μM with the least impact in the GB MES sample at 50 μM (Supplementary Figure 11). Indirect inhibition of MOES with Fasudil HCl led to reduced cell viability in samples of all subgroups at increasing concentrations, but also less in the GB MES sample at 500 μM (Supplementary Figure 12). Hence, overexpression of MOES in GB MES might be protective against inhibition.

## Discussion

### Definition of glioma subgroups and introduction of non-neoplastic cell markers

We provide a mainly descriptive comparative proteome inventory of glioma subgroups sharply defined using the methylomic classification^8^. Thereby, we showed that glioma grade and *IDH* mutation status are the major discriminators for protein expression levels between glioma subgroups.

Gliomas are heterogenous tumors containing not only neoplastic cells but also a network of infiltrating immune, and other cell types. Thus, the observed proteins level variations may also reflect tumor-associated cell composition, which varies from one glioma subtype to another^16^ and whose proportion can make up to 40%^32^ with the highest values of macrophages and microglia infiltration in GB MES^16^. In addition, within each subtype collection, biopsy cell type composition potentially varies from one resection event to another. To exclude proteins mainly associated with immune cells and neurons, we introduced the markers HG2A and PTPRC (CD45) for immune cells, and SEPT3 for neurons into our model as “non-neoplastic cell markers”.

One could argue, that overexpressed proteins in HGG subgroups may simply result from higher cellularity of these tumors. However, from a total of 5057 analyzed proteins only 162 proteins (3%) were differentially expressed, and therefore 97% of all proteins did show similar expression levels.

### The *IDH*-wildtype GB subgroup definition and its evolution over time

An early proteomic study on GB in 2009^5^ delineated three subclasses of GB characterized by distinct patterns of protein expression and activation within glioma-relevant signaling pathways. One subgroup exhibited a predominance of Platelet derived growth factor receptor (PDGFR) activation (reminiscent of GB PN), another displayed a predominance of EGFR activation (reminiscent of GB CL), and a third demonstrated loss of the Rat sarcoma (RAS) regulator Neurofibromin 1 (NF1) (reminiscent of GB MES)^5^. The understanding of the GB subgroups and heterogeneity has been refined in 2019 as Neftel *et. al.* described the GB to exist in four different cellular states^9^. They as well as previous studies have shown that various states or subtypes do coexist within the same GB^9,33,34^. Shift in predominant state or subtype occurs in patients^35^ and can be induced experimentally^4^. Tumor recurrences in GB typically maintain the same state or subtype domination, although studies have shown that chemoradiation holds the potential to induce a transition from proneural to mesenchymal (PMT), akin to the epithelial-mesenchymal transition (EMT) observed in carcinomas, which is associated with therapy resistance^36^. The mechanisms underlying PMT remain poorly understood; however, the tumor microenvironment appears to play a crucial role^,37^. In cancer, the evolutionary trajectory and coexistence of distinct subpopulations of cancer cells within the same tumor likely underlie therapy failure, development of treatment resistance, and eventual recurrence of the malignancy^38^. Therefore, comprehending the composition of GB subpopulations is crucial for effective disease management, and targeted therapy.

### Novelty of our study compared to recent proteomic studies in glioma

Yanovich *et. al.* compared in 2021 proteogenomics vs. transcriptomics (RNA sequencing) in 87 GB samples. They found genes and relevant products that highly correlated with short (common-short cluster) and long survival (common long cluster) in both RNA and protein analysis. With a rather modest correlation between both methods, they could show that proteomics correlated better with survival than transcriptomics. Ultimately, they identified a unique protein-based classification that was distinct from the established RNA-based classification. However, in contrast to our study, they only included GB patients in their study and did not compare the proteomes of the different glioma subgroups with each other^39^.

Duhamel *et al.* presented in 2022 a spatial proteomic characterization in clinical samples of glioblastoma. A non-targeted matrix-assisted laser desorption/ionization mass spectrometry imaging (MALDI-MSI) analysis followed by spatial segmentation using different algorithms allowed to highlight molecular heterogeneity among these tumors. They were able to identify three sub-regions with differing protein expression (Group A: proteins associated with neurogenesis, Group B: immune system activation, and Group C: tumorigenesis) with high intratumoral heterogeneity. Additionally, they found 5 proteins that were significantly associated with short or long survival in GB patients^40^. This study also only included GB patients and like our study, they detected GB sub-regions or subtypes with differing protein expression. However, the proteins of two of their detected patterns (Group A: proteins associated with neurogenesis, and Group B: proteins associated with immune system activation) would not have been detected using our approach.

Lam et. al. leveraged in 2022 mass spectrometry to spatially align abundance levels of 4,794 proteins to distinct histologic tumor niches across 20 GB patients. They defined two distinct proteogenomic programs: MYC– and KRAS-axis hereon, that cooperate with hypoxia to produce a tri-dimensional model of intra-tumoral heterogeneity. Interestingly, high KRAS targets activity was associated with invasion and epithelial-to-mesenchymal transition processes (similar to our results in GB CL and GB MES subgroups), whereas samples enriched for the MYC axis were associated with cell cycle progression (similar to our results in GB PN subgroup)^41^. Contrary to our approach, they did not beforehand define the dominant GB subtype of the individual samples.

To conclude, several studies have recently attempted at defining the proteome and understanding the heterogeneity of glial tumors, and especially GB. Each of these studies used a slightly different approach, and all of those are of complementary value. Our study is different from those studies as we 1) also included LGG and *IDH*-mutated HGG subgroups, 2) used methylomic classification^8^ to obtain a sharp subgroup definition before proteomic analysis, 3) included factors potentially impacting protein expression levels such as age, sex, treatments (chemotherapy, radiotherapy, steroids), and epilepsy as confounding factors into the model and 4) included “non-neoplastic protein markers” (HG2A, PTPRC and SEPT3) prior to analysis to exclude proteins primarily associated with non-neoplastic cells such as immune cells and neurons. We analysed intraoperatively collected high quality proteomic data, selected the most overexpressed DEPs from each subgroup and correlated them with survival data from our glioma cohort, as well as with survival and mRNA data from the TCGA cohort. Functional validation using inhibitor response assays on dissociated tumor cells did show that inhibition of the most overexpressed DEPs and their associated pathways in the I*DH*-wiltype GB subgroups (except RBP1 and TYB10, for which no inhibitor was available) led to reduced cell viability. Thus, these proteins and pathways seem to be important for cell functionality and survival. We could also observe significant differences in cell survival between samples of different GB subgroups. We provide diverse functions and pathways enriched with overexpressed DEPs for each *IDH*-wildtype GB subgroup: translation and cell cycle/telomere regulation in GB PN (linked to proliferation), actin cytoskeleton, cell adhesion and apoptosis regulation in GB CL (linked to migration and invasion), and mitochondrial ATP synthesis in GB MES (linked to metabolism), a novel finding that has not been reported in this way previously.

For all these reasons, our study adds complementary value and substantially amends the current knowledge of the proteome of glial tumors.

### Pathway analyses revealed different biological processes and molecular functions in each glioma subgroup

The major discriminators for protein expression levels in our study were glioma grade and *IDH* mutation status, as shown previously in a study comparing glioma subgroups of various grades and *IDH* mutation status^42^.

Many of the overexpressed DEPs in GB PN were associated with functions related to translation and protein synthesis. GB may potentially boost inherent ribosome production by dysregulation of ribosome biogenesis and assimilating external ribosomes that encourage reprogramming within GB cells^43^. The utilization of ribosome biogenesis inhibition as therapeutic approach has shown promising outcomes including decreased viability of glioma cells, induction of apoptosis, and hindered growth of transplanted glioma cells in zebrafish^44^. This therapeutic option would therefore be potentially applicable to tumors of this GB subgroup.

Several studies have previously identified enrichment of DNA replication and cell cycle gene transcripts in the GB PN subtype^45^. Sustaining telomere length, either via increased *TERT* expression resulting from *TERT* promoter mutations, or through alternative lengthening of telomeres (ALT) resulting from *Alpha-thalassemia/mental retardation X-linked* (*ATRX)* mutation, appears to be a crucial aspect of GB development^32^. A study from 2010 showed that GB patients with tumors with ALT had longer survival that was independent of age, surgery, and other treatments. This could be attributed to the fact that most ALT+ tumors are less aggressive GB PN subtypes^46^, whereas *TERT* promoter mutations appear to be less common in GB PN than in GB CL or GB MES (29.7%, 41.8%, and 41.6%, respectively)^47^.

The ‘regulation of the actin cytoskeleton’ emerged as a prominent function among the proteins significantly overexpressed in GB CL. The dynamic processes of GB cell migration and invasion necessitate the restructuring of the actin cytoskeleton to alter cell shape and propel cell movement. Recent research suggests that remodeling of the actin cytoskeleton might enables cancer cell resistance to antitumor immunity^48^. A study has unveiled a potential association between elevated expression of Transgelin-2 (TAGL2), invasion, and unfavorable patient outcomes in *IDH*-wildtype gliomas^49^. Interestingly, several proteins linked with invasion were significantly overexpressed in the GB CL: MOES, EGFR, Annexin A5 (ANXA5), Vimentin (VIM), Tenascin C (TENA), and Cysteine and glycine-rich protein 2 (CSRP2). Elevated MOES expression in GB cells leads to increased cell proliferation, invasion, and migration in vitro and in vivo^50^. High levels of *EGFR* amplification serve as a marker for GB CL^5,6,51^. ANXA5 promotes invasion and chemoresistance to temozolomide in GB cells, correlating with worse prognosis^52^. High VIM expression signifies epithelial-mesenchymal transition and is associated with poor survival in gliomas and particularly in the GB CL subtype^51^. TENA is linked to tumor progression and poor prognosis, and promotes migration^53^.

Another enriched function among the DEPs overexpressed in GB CL is regulation of apoptosis and cell death. Among mediators of these processes are EGFR, ANXA5, LEG3, GPNMB, Nestin (NES), Heat shock protein beta-6 (HSPB6), and Transcription intermediary factor 1-alpha (TRIM24). The phosphatidylinositol-3 kinase (PI3K)/protein kinase B (AKT)/mammalian target of rapamycin (mTOR) pathway, downstream of receptor tyrosine kinase pathway to which EGFR belongs, plays a pivotal role in cell survival, proliferation, motility, angiogenesis, and apoptosis, and is deregulated in approximately 80% of all GB cases^54^. Phosphatase and TENsin homolog (PTEN) counteracts the PI3K/AKT/mTOR pathway, and its loss is a hallmark of GB. *EGFR* amplification and *PTEN* loss particularly co-occur in GB^55^. TRIM24, as activator of PI3K/AKT signaling pathway, promotes glioma progression and enhances chemoresistance, while also promoting stemness and invasiveness of GB cells via Sox2 expression^56^. Silencing of Histone H1.5 (H15) has been shown to trigger glioma cell apoptosis^57^.

IQGA1, the only protein significantly overexpressed in GB CL compared to the pool of other HGG, is involved in cell adhesion and migration of glioma cells and its expression correlates with poor tumor grade and survival^58^. It is a known scaffold protein in glioma stem cell niches^59^.

Citrate synthase and oxidative phosphorylation complexes gradually decrease during progression from LGG to HGG^60^. The S10AD overexpression we found in GB MES compared to LGG refines S10AD overexpression previously observed in HGG, correlating with tumor grade and micro-vessel density^61^.

### Strengths and limitations of our study

This study provides comprehensive proteomic insights by analyzing and quantifying 5057 proteins for each tumor, with only minimal missing values per protein The strengths of our study can be seen in the prospective sample collection and sharp subgroup definition before proteomic analysis, using the methylomic classification. Gliomas are known for their heterogeneity, encompassing up to 40% of non-neoplastic cells, such as infiltrating immune cells, and resident neurons ^32^. Three protein markers (HG2A and PTPRC (CD45) for immune cells, and SEPT3 for neurons) were introduced into the model to exclude the over– and underexpressed proteins mainly associated with those non-neoplastic cells, to rather focus on the proteome of neoplastic cells of the defined glioma subtypes. However, this offers only an approximation of reality and possibly obviates the detection of interesting results, as there were many DEPs between *IDH* HGG and LGG in the uncorrected model (results not shown), and only one DEP remaining after accounting for these confounders. While CD45 has been utilized as a marker in proteomic studies before and has shown good correlation with CD45 immunohistochemistry^62^, there is limited data available regarding the use of SEPT3 and HG2A as biomarkers in proteomic and other studies.

We could observe distinct patterns of pathways and functions containing overexpressed DEPs in all three *IDH-* wildtype GB subtypes: Translation and cell cycle/telomere regulation in GB PN (essential for cell proliferation), actin cytoskeleton, cell adhesion, and cell death/apoptosis regulation in GB CL (essential for cell migration and invasion), and mitochondrial ATP synthesis in GB MES (essential for metabolism). We provided potential novel biomarkers for each of the glioma subgroups and correlated them with survival data of our own cohort as well as with mRNA expression and survival data from the TCGA network. We performed functional validation of those DEPs with inhibitor response assays in patient-derived, non-cultured and subgroup defined GB samples. These assays showed effective, dose-dependent responses in all samples, and trended towards identification of subgroup specific responses in a subset of the assessed signature protein inhibitors. There were specific inhibitors available for some of the proteins, for others we had to use more broad or indirect inhibition of protein effect or associated pathways. However, these are preliminary results that should be interpreted cautiously. Larger cohorts of subtyped single cell suspensions and more stringent dose-response curves assessing the subtype-defining DEPs from our dataset might in the future be used to stratify patients and use these screens to personalize GB treatment.

The results are focusing on the HGG, and especially the *IDH-*wildtype GB subgroups, with the aim of a better understanding of the heterogeneity and metabolism of these highly malignant tumors. However, as described above, we are just at the beginning of understanding the composition and heterogeneity of these tumors. To date, there is no clinical utility of the GB subtypes, and they are also not part of any WHO classification. Therefore, the results of this study are mainly of explorative and descriptive nature.

The constrained sample sizes within subgroups may have impacted the statistical power of differential protein expression between groups. If comparing *IDH* HGG to LGG, only one protein was found to be significantly differentially expressed, likely reflecting the common *IDH*-mutant background of these two subgroups and its major impact on tumor metabolism and protein expression patterns^42^.

Some of the clinical and demographic data was collected retrospectively, thus inheriting the limitations associated with such data collection.

## Conclusion

Quantitative proteomic analysis across glioma subgroups highlighted tumor grade and *IDH* mutation status as the primary determinants of protein level variations. Delving deeper into the expression profiles of the three *IDH*-wildtype GB subgroups compared to LGG revealed distinct patterns of overexpressed proteins associated with glioma progression-related pathways: Cell proliferation in GB PN, cell migration and invasion in GB CL, and metabolism in GB MES. Functional validation using inhibitor response assays did show that inhibition of the most overexpressed DEPs and their associated pathways led to reduced cell viability, hence these proteins seem to be relevant for the functionality and survival of the tumor cells.

This mainly descriptive study provides valuable insights into tumor subgroup metabolism and potential biomarkers for further experimental testing. Better understanding of the metabolic dynamics of GB and its heterogeneity and subgroups is crucial for informing disease management strategies and facilitating future targeted therapy for this lethal malignancy.

## Author contributions

**Amer Salem M:** Data Curation, Investigation, Methodology, Visualization; **Boulay JL:** Conceptualization, Data curation, Investigation, Methodology, Writing – original draft**; Ritz MF:** Data curation, Investigation, Methodology, Resources, Writing – Original Draft; **Halbeisen FS:** Formal analysis, Validation, Visualization, Writing – Review and Editing; **Tschan VJ:** Investigation, Validation, Visualization, Formal analysis, Writing – Review and Editing; **Schmidt A:** Investigation, Methodology; **Buczak K:** Data curation, Formal analysis, Investigation, Methodology, Validation; **Hutter G:** Funding acquisition, Resources, Writing – Review and Editing; **Leu S:** Conceptualization, Data curation, Funding acquisition, Investigation, Methodology, Supervision, Validation, Visualization, Writing – original draft.

## Data Statement

The generated proteomic glioma data inventory was uploaded to the public MassIVE database repository (project accession number: PXD041647), and the raw data from the inhibitor response assays was uploaded to Mendeley Data (10.17632/msr3c3rnt6.1).

## Data Availability

All data produced in the present study are available upon reasonable request to the authors.
The generated proteomic glioma data inventory was uploaded to the public MassIVE database repository (project accession number: PXD041647), and the raw data from the inhibitor response assays was uploaded to Mendeley Data (10.17632/msr3c3rnt6.1).

https://massive.ucsd.edu/ProteoSAFe/private-dataset.jsp?task=07d1eb41382b48dd8dd03b77af856077

https://data.mendeley.com/datasets/msr3c3rnt6/1

## Acknowledgements

We thank Heike Neddersen for collecting demographic data, Cristòbal Tostado for *TERT* promotor sequencing, Jürgen Hench and Stephan Frank for providing EPIC methylomics data and CNV data, Wandrille Duchemin and Sabrina Hogan for bioinformatic input, and for the statistical analysis of the functional validation data, and Luigi Mariani and Raphael Guzman for constant support.

## Abbreviations and glossary

Å: Ångstrom
AC-like: Astrocyte-like
AKT: Protein kinase B
ALT: Alternative lengthening of telomere
ATP: Adenosine triphosphate
*ATRX*: Alpha-thalassemia/mental retardation X-linked
*CDKN2A*: *Cyclin-dependent kinase inhibitor 2A*
CNS: Central nervous system
CNV: Copy number variations
DEP: Differentially expressed protein
DKFZ: Deutsches Krebsforschungszentrum
EKNZ: Ethikkommission Nordwest-und Zentralschweiz
EMT: Epithelial-to-mesenchymal transition
GB: Glioblastoma
GB CL: *IDH*-wildtype glioblastoma, subtype RTK II (classical)
GB MES: *IDH*-wildtype glioblastoma, subtype mesenchymal
GB PN: *IDH*-wildtype glioblastoma, subtype RTK I (proneural)
GO: Gene ontology
GSEA: Gene Set Enrichment Analysis
HGG: High grade glioma
HPLC: High performance liquid chromatography
*IDH 1/2*: *Isocitrate dehydrogenase 1/2*
*IDH* CODEL: *IDH*-mutant, 1p/19q co-deleted oligodendroglioma
*IDH* HGG: *IDH*-mutant high-grade glioma
*IDH* LGG: *IDH*-mutant glioma
KEGG: Kyoto Encyclopedia of Genes and Genomes
LC: Liquid chromatography
LGG: Low grade glioma
log_2_FC: log_2_ fold change
M: Molarity
MALDI-MSI: Matrix-assisted laser desorption/ionization mass spectrometry imaging
MES-like: Mesenchymal-like
MS: Mass spectrometry
mTOR: Mammalian target of rapamycin
NF1: Neurofibromin 1
NPC-like: Neural progenitor-like
OPC-like: Oligodendrocyte-progenitor-like
ORA: Over-representation analysis
PCA: Principal component analysis
PI3K: Phosphatidylinositol-3 kinase
PMT: Proneural-to-mesenchymal transition
PDGFR: Platelet-derived growth factor receptor
PPI: Protein-protein interaction
PTEN: Phosphatase and TENsin homolog
RAS: Rat sarcoma
RTK I and II: Receptor-tyrosine kinase I and I
TCGA: The Cancer Genome Atlas
TERT: Telomerase Reverse Transcriptase
TFA: Trifluoroacetic acid
TMT: Tandem mass tag
TP53: Tumor protein 53
UMAP: Uniform Manifold Approximation and Projection
WHO: World Health Organisation

## Proteins and short names

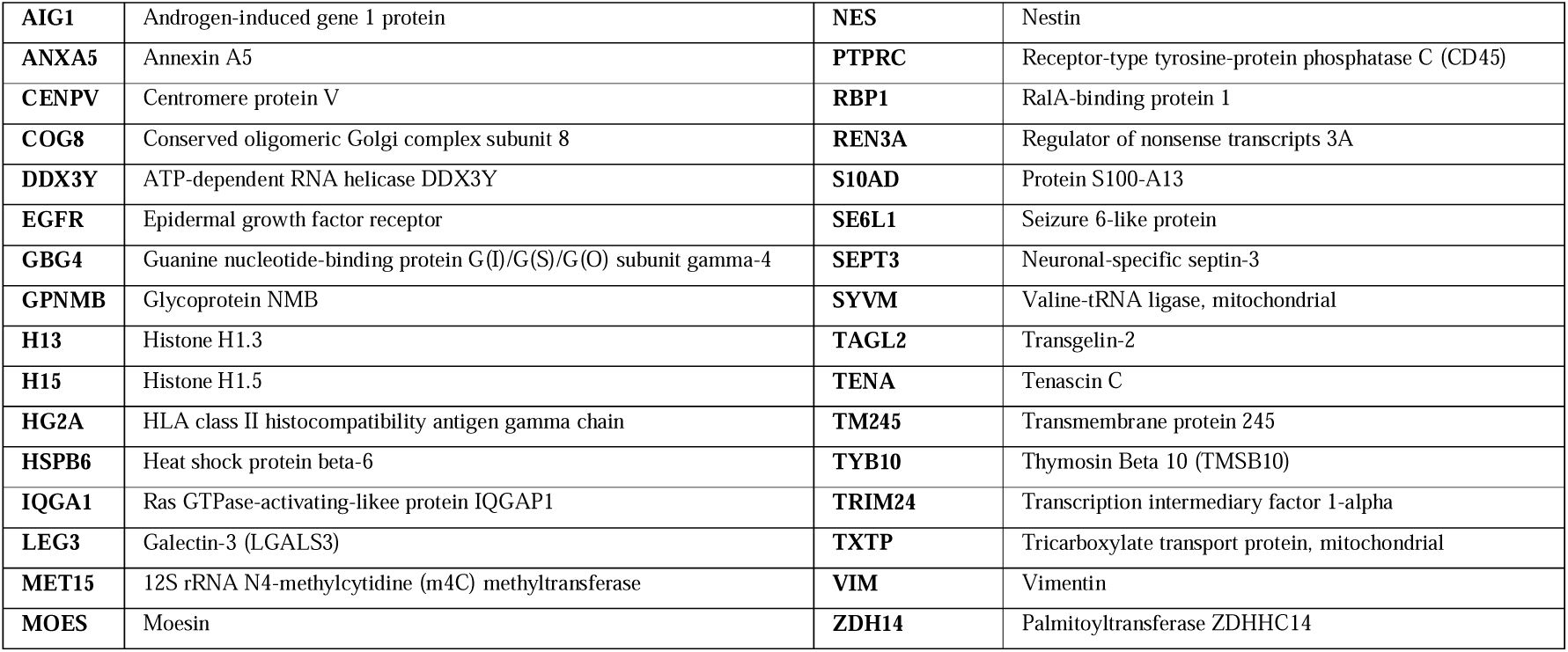

|  |  |  |  |
| --- | --- | --- | --- |
| **AIG1** | Androgen-induced gene 1 protein | **NES** | Nestin |
| **ANXA5** | Annexin A5 | **PTPRC** | Receptor-type tyrosine-protein phosphatase C (CD45) |
| **CENPV** | Centromere protein V | **RBP1** | RalA-binding protein 1 |
| **COG8** | Conserved oligomeric Golgi complex subunit 8 | **REN3A** | Regulator of nonsense transcripts 3A |
| **DDX3Y** | ATP-dependent RNA helicase DDX3Y | **S10AD** | Protein S100-A13 |
| **EGFR** | Epidermal growth factor receptor | **SE6L1** | Seizure 6-like protein |
| **GBG4** | Guanine nucleotide-binding protein G(I)/G(S)/G(O) subunit gamma-4 | **SEPT3** | Neuronal-specific septin-3 |
| **GPNMB** | Glycoprotein NMB | **SYVM** | Valine-tRNA ligase, mitochondrial |
| **H13** | Histone H1.3 | **TAGL2** | Transgelin-2 |
| **H15** | Histone H1.5 | **TENA** | Tenascin C |
| **HG2A** | HLA class II histocompatibility antigen gamma chain | **TM245** | Transmembrane protein 245 |
| **HSPB6** | Heat shock protein beta-6 | **TYB10** | Thymosin Beta 10 (TMSB10) |
| **IQGA1** | Ras GTPase-activating-likee protein IQGAP1 | **TRIM24** | Transcription intermediary factor 1-alpha |
| **LEG3** | Galectin-3 (LGALS3) | **TXTP** | Tricarboxylate transport protein, mitochondrial |
| **MET15** | 12S rRNA N4-methylcytidine (m4C) methyltransferase | **VIM** | Vimentin |
| **MOES** | Moesin | **ZDH14** | Palmitoyltransferase ZDHHC14 |

