## Supplementary Figure 1 for "Quantitative proteomic analysis reveals different functional subtypes among IDH-wildtype glioblastoma"

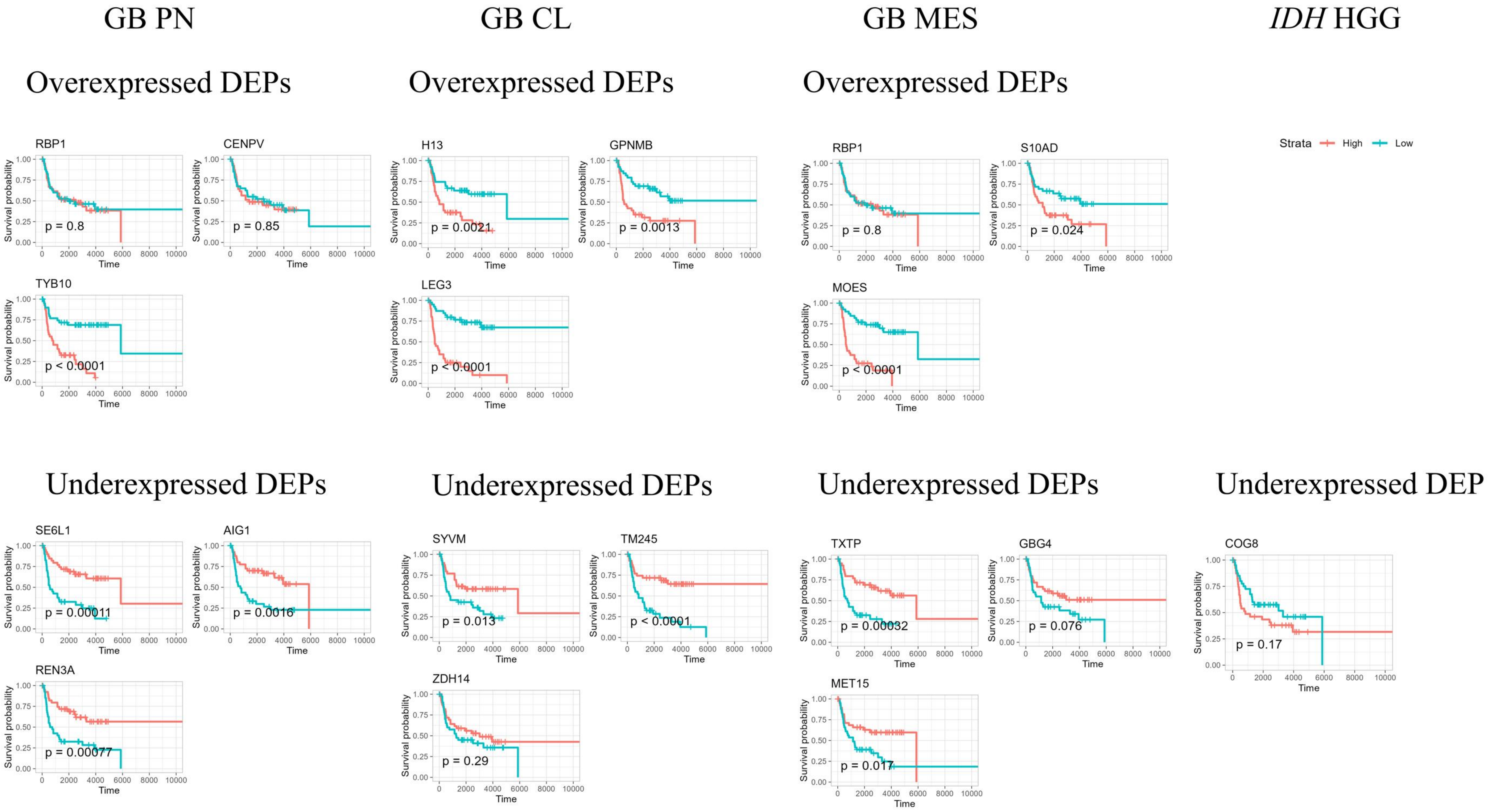


**Supplementary Figure 1:** Association of the three most overexpressed and three most underexpressed DEPs from each subgroup with overall survival. (DEP abundance was split into high and low expression groups at the median, Kaplan-Meier survival analysis was done to estimate survival curves, and the log-rank test was used to assess differences between the groups).
