## Supplementary Figure 2 for "Quantitative proteomic analysis reveals different functional subtypes among IDH-wildtype glioblastoma"

DEPs overexpressed in GB PN vs. LGG

DEPs underexpressed in GB PN vs. LGG

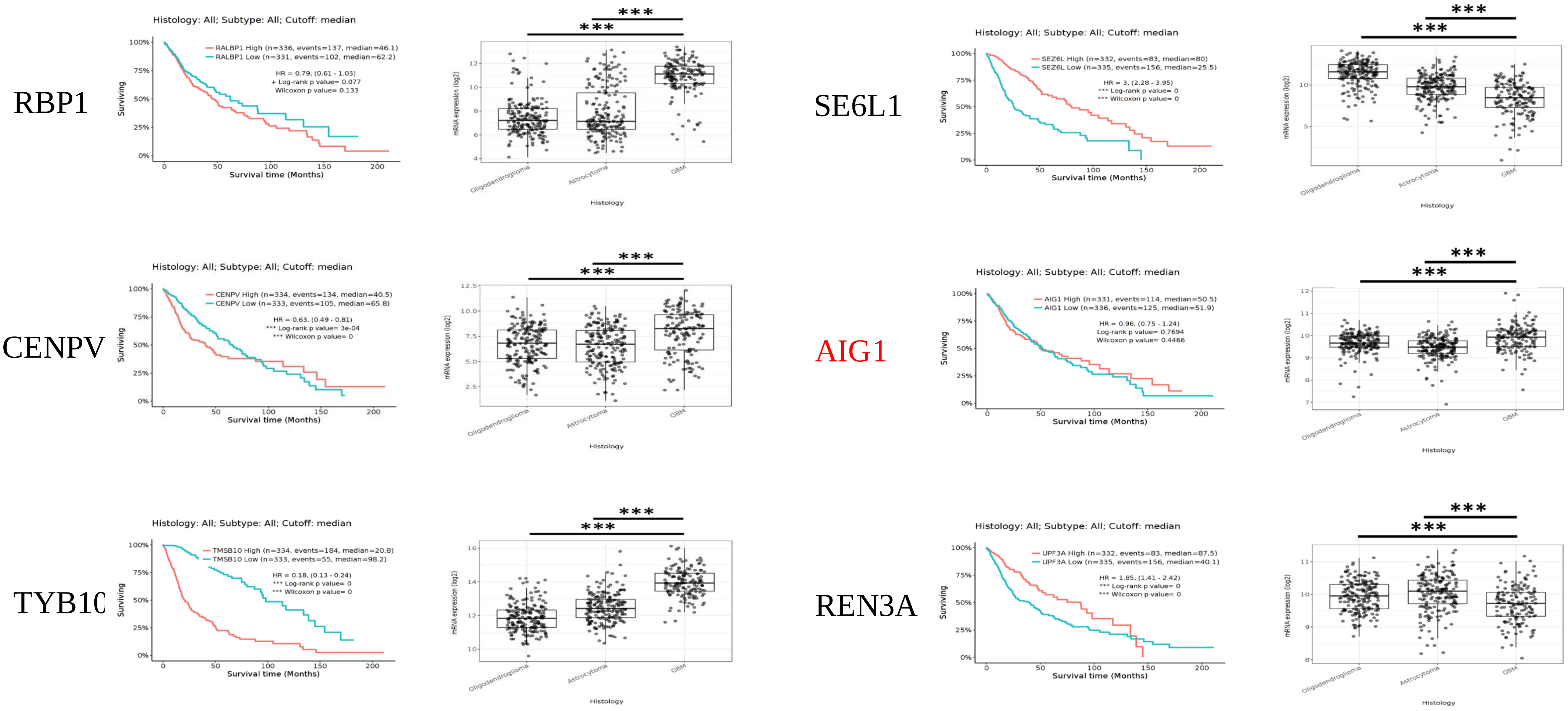


**Supplementary Figure 2:** TCGA data using the TCGA GBM-LGG cohort showing the mRNA expression levels of the DEPs most over- and underexpressed in GB PN, and association of high and low mRNA levels of these DEPs with survival. Note: in red listed are the proteins that have contradicting expression levels and association with survival in the TCGA GBM-LGG database compared to our data (www.tcga.gov).
