## Supplementary Figure 3 for "Quantitative proteomic analysis reveals different functional subtypes among IDH-wildtype glioblastoma"

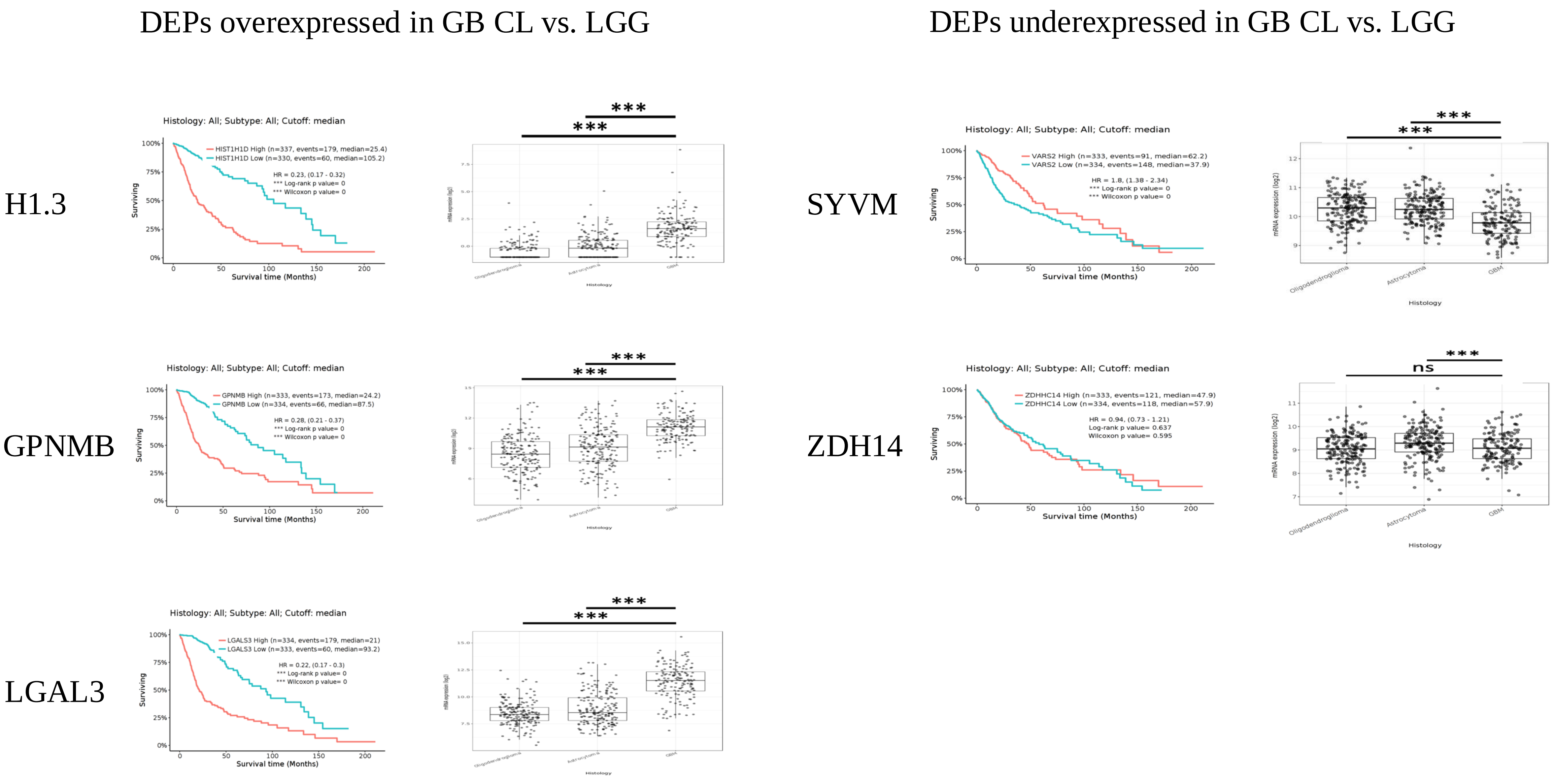


**Supplementary Figure 3:** TCGA data using the TCGA GBM-LGG cohort showing the mRNA expression levels of the DEPs most over- and underexpressed in GB CL, and association of high and low mRNA levels of these DEPs with survival. There were no results available for mRNA expression of TM245 in glioma (www.tcga.gov).
