## Supplementary Figure 4 for "Quantitative proteomic analysis reveals different functional subtypes among IDH-wildtype glioblastoma"

### DEPs overexpressed in GB MES vs. LGG

### DEPs underexpressed in GB MES vs. LGG

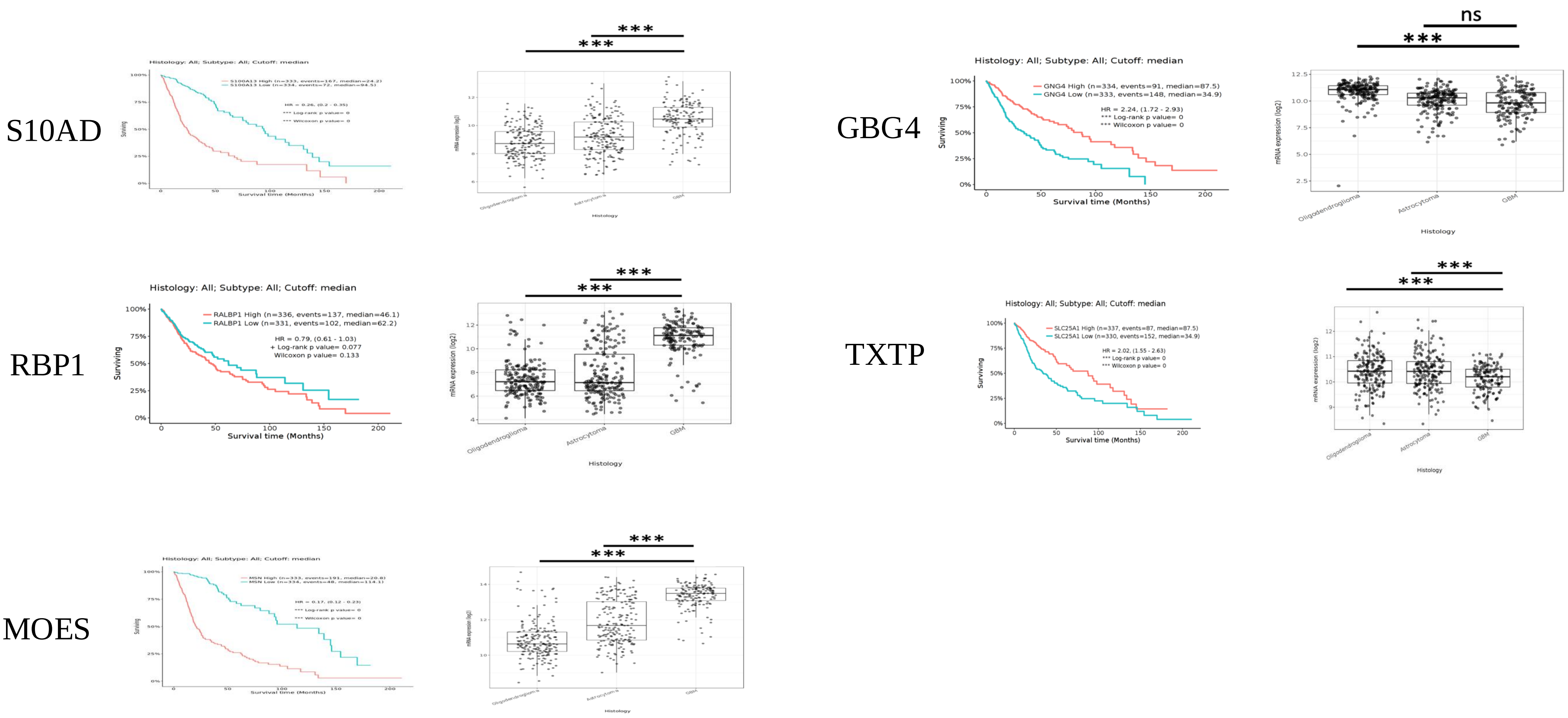


**Supplementary Figure 4:** TCGA data using the TCGA GBM-LGG cohort showing the mRNA expression levels of the DEPs most over- and underexpressed in GB MES compared to LGG, and association of high and low mRNA levels of these DEPs with survival. There were no results available for mRNA expression of MET15 in glioma (www.tcga.gov).
