## Supplementary Figure 5 for "Quantitative proteomic analysis reveals different functional subtypes among IDH-wildtype glioblastoma"

### DEP underexpressed in *IDH* HGG vs. LGG

COG8

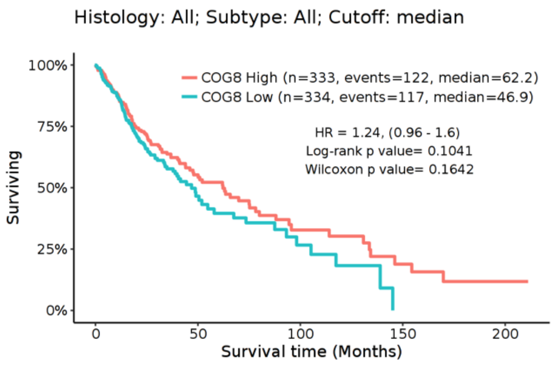


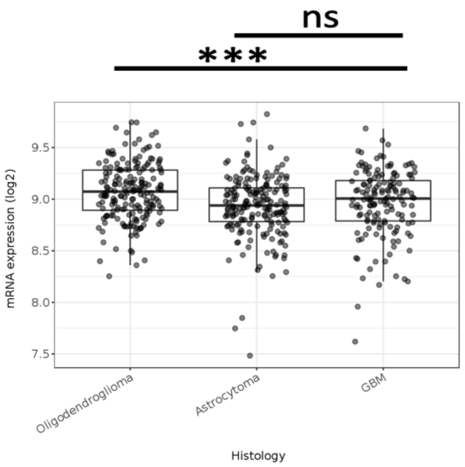


**Supplementary Figure 5:** TCGA data using the TCGA GBM-LGG cohort showing the mRNA expression level of the only DEP significantly underexpressed in IDH HGG compared to LGG, and association of high and low mRNA levels of this DEP with survival (www.tcga.gov).
