## Supplementary Figure 6 for "Quantitative proteomic analysis reveals different functional subtypes among IDH-wildtype glioblastoma"

### Inhibitor response assay with GB1107 -> targeting LEG3 – overexpressed in GB CL

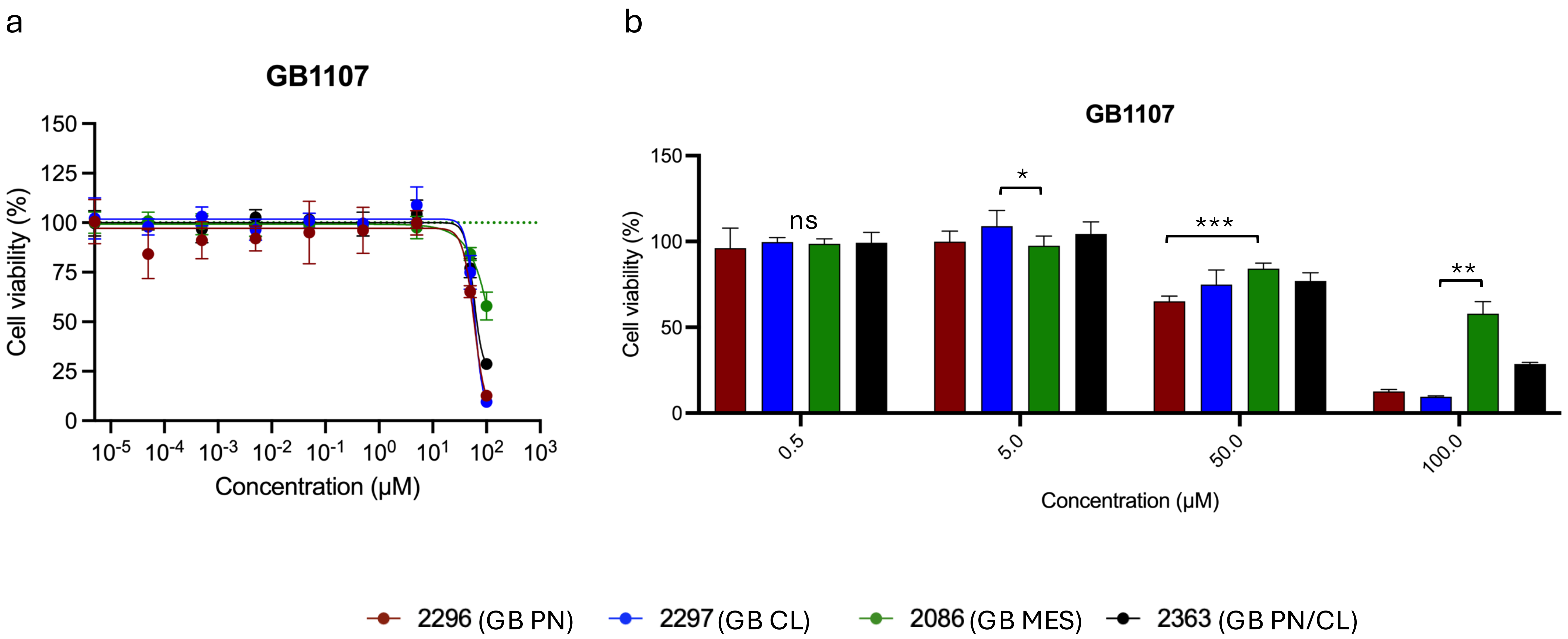


**Supplementary Figure 6: Inhibitor response assay using GB1107 targeting LEG3**

a) Dose response curves for dissociated tumor cells of patients representative of GB PN (red), GB CL (blue), GB MES (green) and GB PN/CL (black).

b) Percentage of cell viability for the four highest concentrations of GB1107 applied in inhibitor assays. Results at 500 μM not shown due to precipitation. * indicates significant difference
