## Supplementary Figure 7 for "Quantitative proteomic analysis reveals different functional subtypes among IDH-wildtype glioblastoma"

### Inhibitor response assay with Glembatumumab vedotin -> targeting GPNMB – overexpressed in GB CL

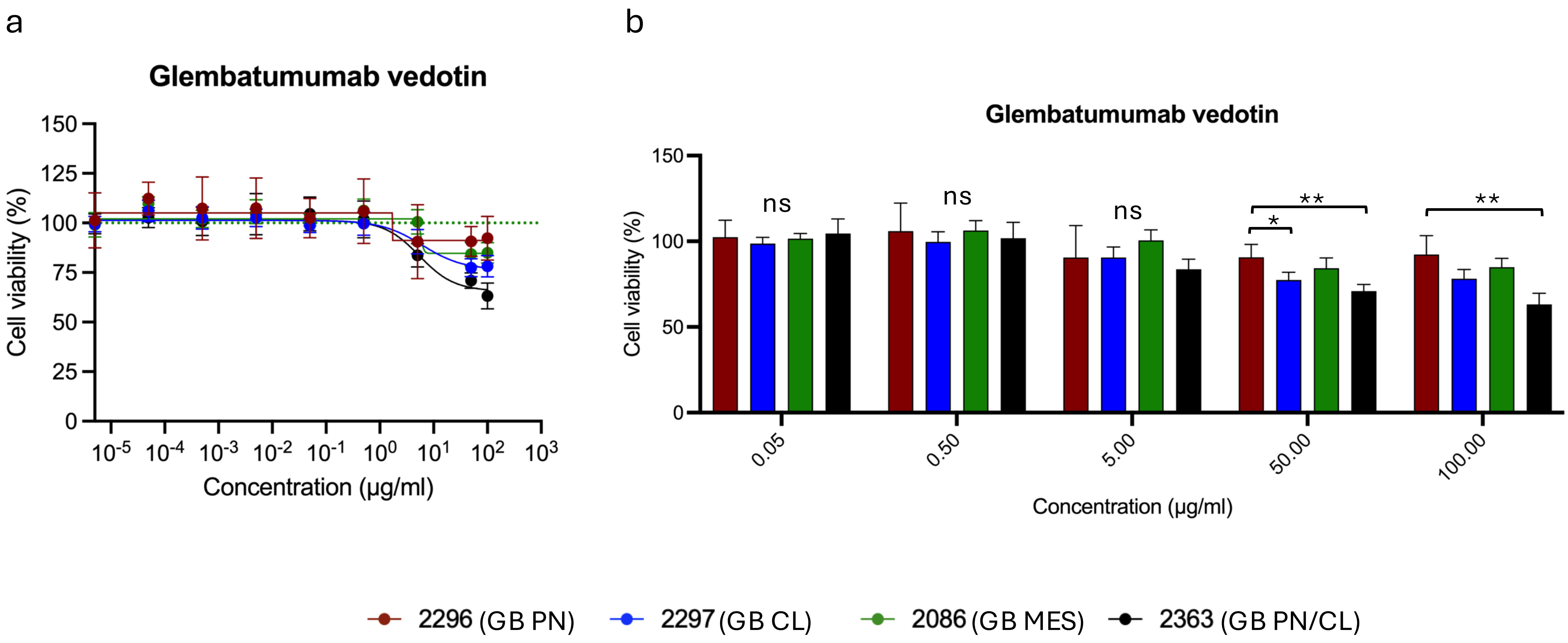


**Supplementary Figure 7: Inhibitor response assay using Glembatumumab vedotin targeting GPNMB**

a) Dose response curves for dissociated tumor cells of patients representative of GB PN (red), GB CL (blue), GB MES (green) and GB PN/CL (black).

b) Percentage of cell viability for the five highest concentrations of Glembatumumab vedotin applied in inhibitor assays. * indicates significant difference
