## Supplementary Figure 8 for "Quantitative proteomic analysis reveals different functional subtypes among IDH-wildtype glioblastoma"

### Inhibitor response assay with Vorinostat -> targeting H13 – overexpressed in GB CL

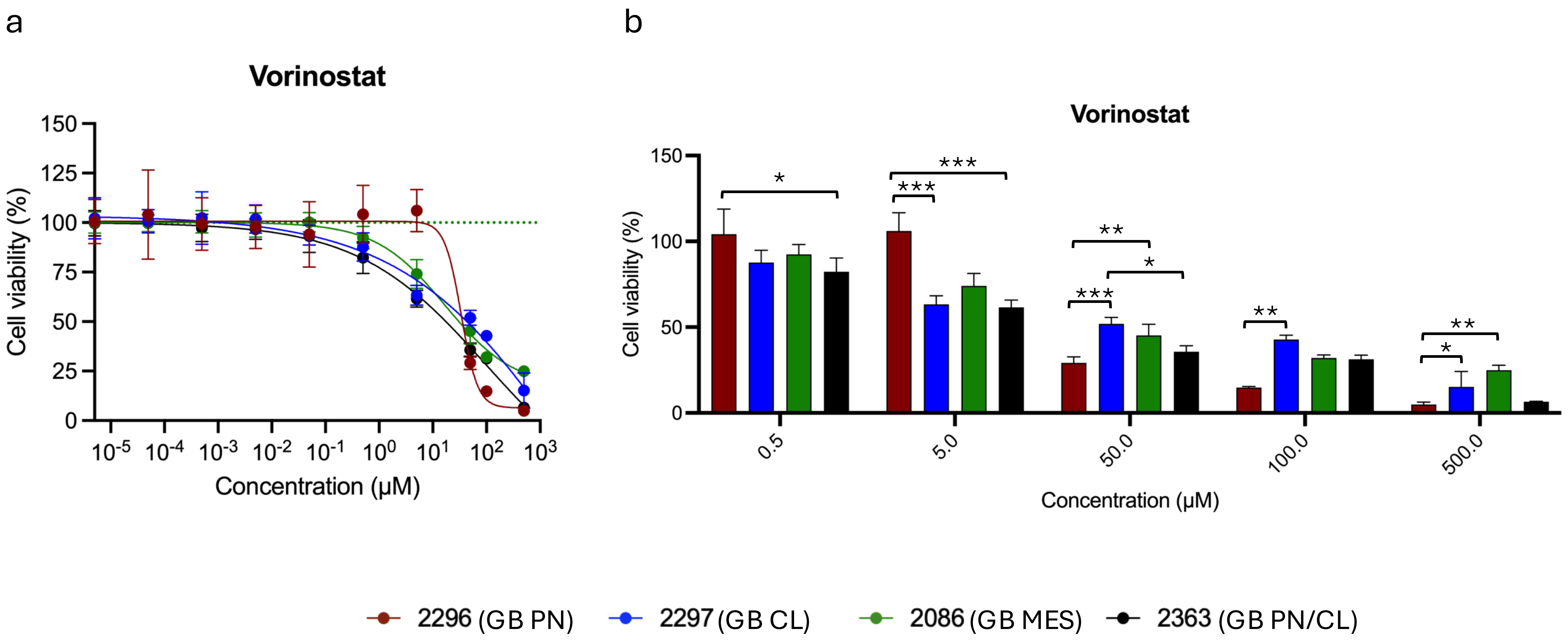


**Supplementary Figure 8: Inhibitor response assay using Vorinostat targeting H13**

a) Dose response curves for dissociated tumor cells of patients representative of GB PN (red), GB CL (blue), GB MES (green) and GB PN/CL (black).

b) Percentage of cell viability for the five highest concentrations of Vorinostat applied in inhibitor assays. * indicates significant difference
