## Supplementary Figure 9 for "Quantitative proteomic analysis reveals different functional subtypes among IDH-wildtype glioblastoma"

### Inhibitor response assay with GSK-923295 -> targeting CENPV – overexpressed in GB PN

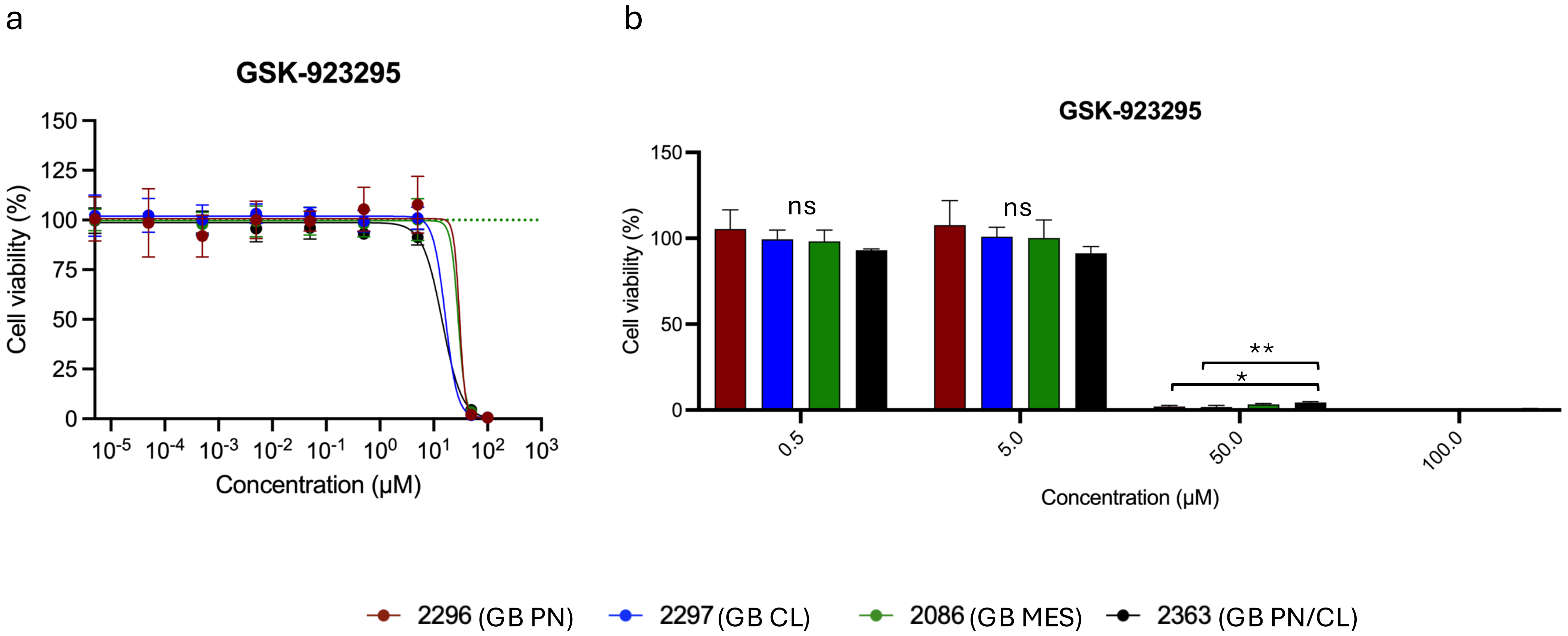


**Supplementary Figure 9: Inhibitor response assay using GSK-923295 targeting CENPV**

a) Dose response curves for dissociated tumor cells of patients representative of GB PN (red), GB CL (blue), GB MES (green) and GB PN/CL (black).

b) Percentage of cell viability for the four highest concentrations of GSK-923295 applied in inhibitor assays. Results at 500 µM not shown due to precipitation. * indicates significant difference
