## Supplementary Figure 10 for "Quantitative proteomic analysis reveals different functional subtypes among IDH-wildtype glioblastoma"

### Inhibitor response assay with Amlexanox -> targeting S10AD – overexpressed in GB MES

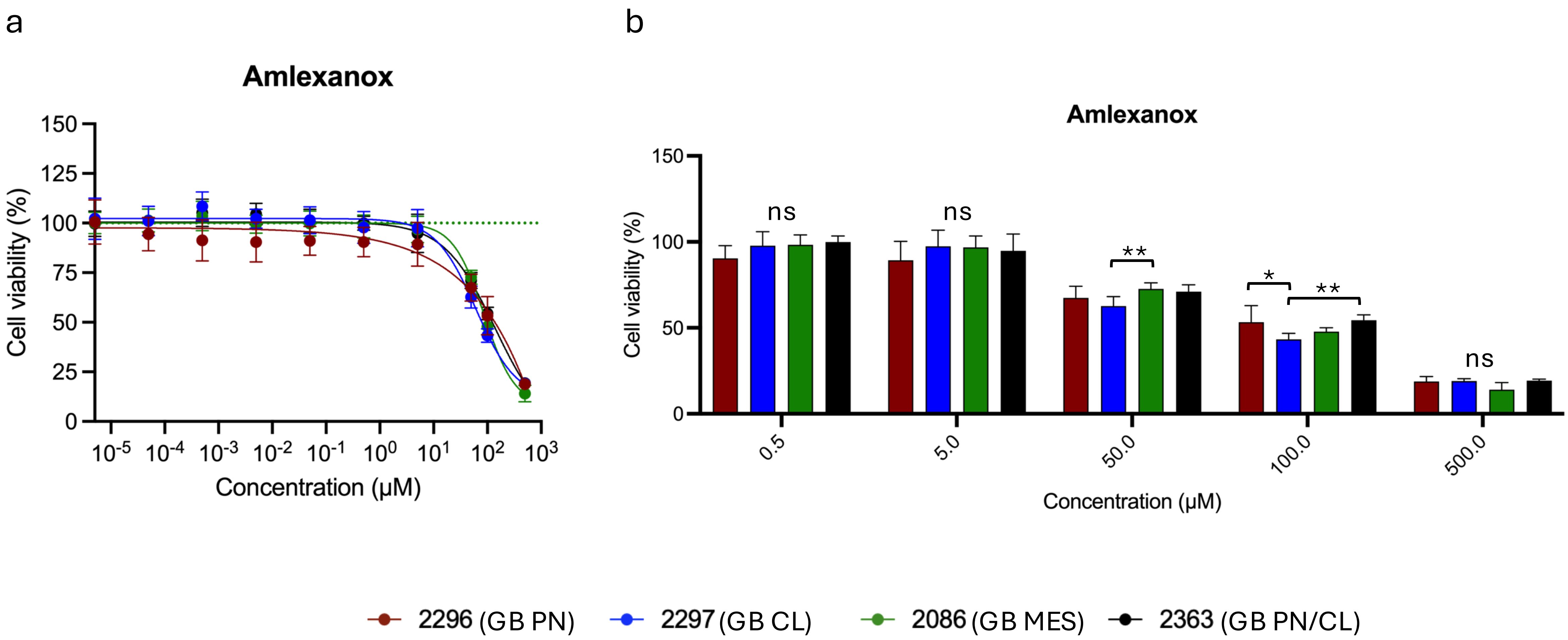


**Supplementary Figure 10: Inhibitor response assay using Amlexanox targeting S10AD**

a) Dose response curves for dissociated tumor cells of patients representative of GB PN (red), GB CL (blue), GB MES (green) and GB PN/CL (black).

b) Percentage of cell viability for the five highest concentrations of Amlexanox applied in inhibitor assays. * indicates significant difference
