## Supplementary Figure 11 for "Quantitative proteomic analysis reveals different functional subtypes among IDH-wildtype glioblastoma"

### Inhibitor response assay with NSC668394 -> targeting MOES – overexpressed in GB MES

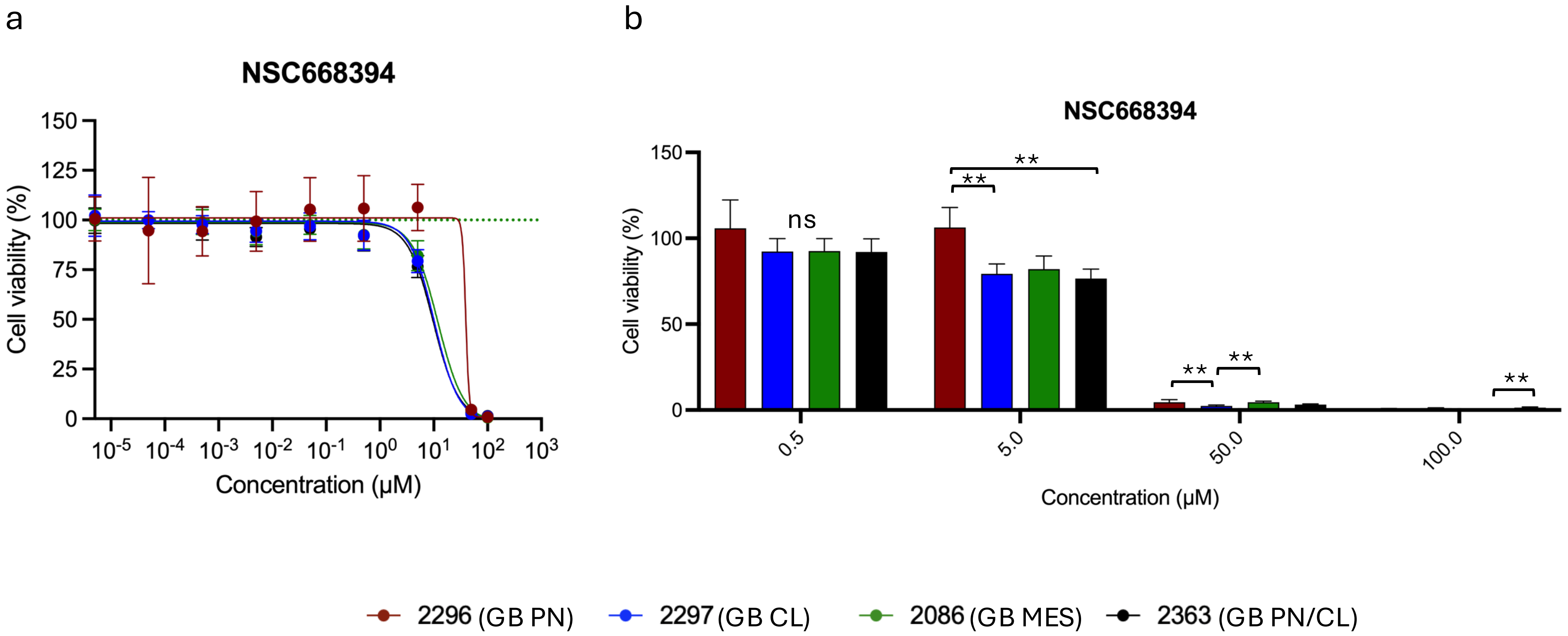


**Supplementary Figure 11: Inhibitor response assay using NSC668394 targeting MOES**

a) Dose response curves for dissociated tumor cells of patients representative of GB PN (red), GB CL (blue), GB MES (green) and GB PN/CL (black).

b) Percentage of cell viability for the four highest concentrations of NSC668394 applied in inhibitor assays. Results at 500 µM not shown due to precipitation. * indicates significant difference
