## Supplementary Figure 12 for "Quantitative proteomic analysis reveals different functional subtypes among IDH-wildtype glioblastoma"

### Inhibitor response assay with Fasudil HCl -> targeting MOES – overexpressed in GB MES

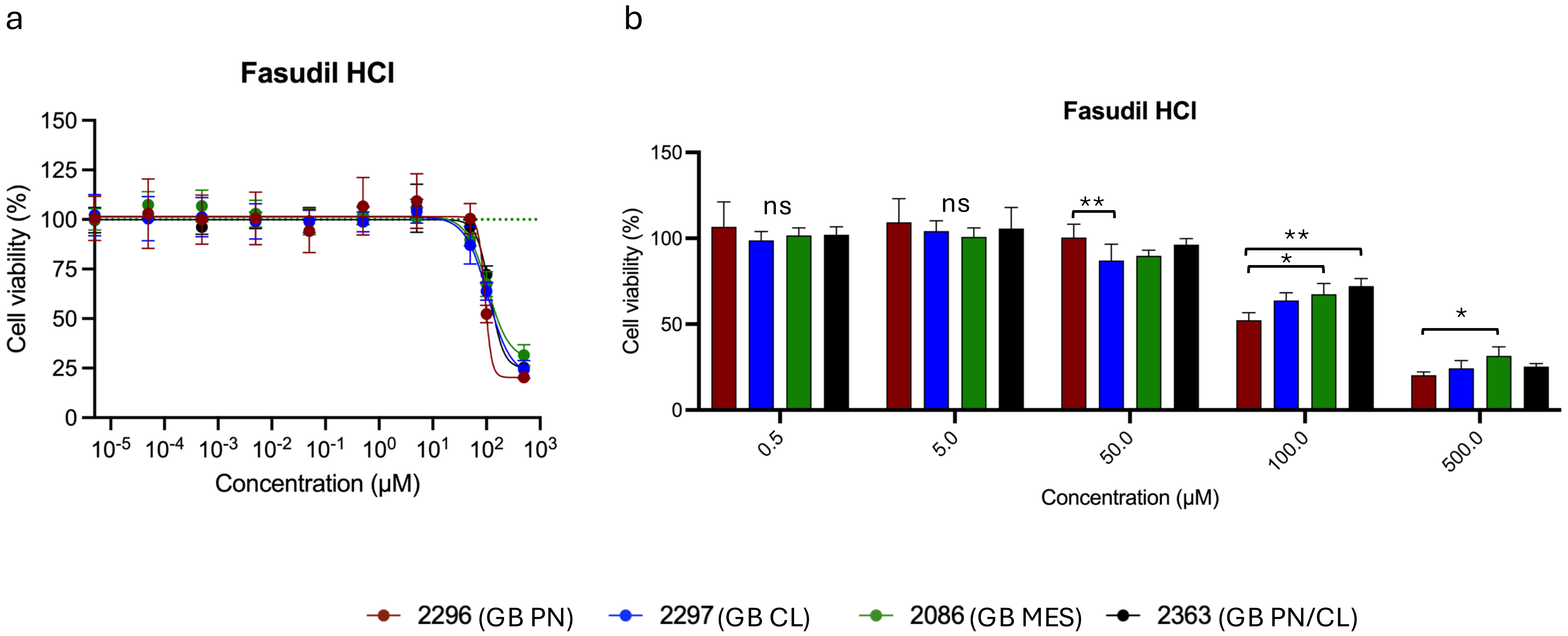


**Supplementary Figure 12: Inhibitor response assay using Fasudil HCl targeting MOES**

a) Dose response curves for dissociated tumor cells of patients representative of GB PN (red), GB CL (blue), GB MES (green) and GB PN/CL (black).

b) Percentage of cell viability for the five highest concentrations of Fasudil HCl applied in inhibitor assays. * indicates significant difference
