## Supplementary Table 1 for "Quantitative proteomic analysis reveals different functional subtypes among IDH-wildtype glioblastoma"

**Supplementary Table 1:** experimental design of the TMT proteomics experiment

| row.names | exp1 | exp2 | exp3 | exp4 | exp5 | exp6 | exp7 |
|---|---|---|---|---|---|---|---|
| 126 | IDHLGG_001 | IDHLGG_003 | IDHLGG_005 | IDHLGG_007 | IDHLGG_009 | IDHLGG_011 | IDHHGG_013 |
| 127N | IDHHGG_014 | IDHHGG_016 | IDHHGG_018 | IDHHGG_020 | IDHHGG_022 | IDHHGG_024 | IDHLGG_025 |
| 127C | IDHLGG_026 | IDHLGG_028 | IDHLGG_030 | IDHLGG_032 | IDHLGG_034 | IDHLGG_035 | IDHLGG_036 |
| 128N | IDHCODEL_03[illegible] | IDHCODEL_04[illegible] | IDHCODEL_042 | IDHCODEL_04[illegible] | IDHCODEL_04[illegible] | IDHCODEL_04[illegible] | GBPN_048 |
| 128C | GBPN_050 | GBPN_052 | GBPN_054 | GBPN_055 | GBPN_056 | GBCL_058 | GBCL_060 |
| 129N | GBCL_062 | GBCL_064 | GBCL_065 | GBCL_066 | GBCL_068 | GBCL_070 | NA |
| 129C | GBMES_074 | GBMES_075 | GBMES_076 | GBMES_078 | GBMES_080 | GBMES_082 | GBMES_084 |
| 130N | GBMES_086 | NA | NA | NA | NA | NA | pool_15 |
| 130C | IDHLGG_002 | IDHLGG_004 | IDHLGG_006 | IDHLGG_008 | IDHLGG_010 | IDHLGG_012 | pool_13 |
| 131N | IDHHGG_015 | IDHHGG_017 | IDHHGG_019 | IDHHGG_021 | IDHHGG_023 | pool_11 | pool_14 |
| 131C | NA | IDHLGG_029 | IDHLGG_031 | IDHLGG_033 | pool_9 | pool_12 | IDHCODEL_037 |
| 132N | IDHCODEL_03[illegible] | IDHCODEL_041 | IDHCODEL_043 | pool_7 | pool_10 | IDHCODEL_04[illegible] | GBPN_049 |
| 132C | GBPN_051 | GBPN_053 | pool_5 | pool_8 | GBCL_057 | GBCL_059 | NA |
| 133N | GBCL_063 | pool_3 | pool_6 | GBCL_067 | GBCL_069 | GBCL_071 | NA |
| 133C | pool_1 | pool_4 | GBMES_077 | GBMES_079 | GBMES_081 | GBMES_083 | GBMES_085 |
| 134 | pool_2 | NA | NA | NA | NA | NA | pool_16 |
