## Supplementary Table 2 for "Quantitative proteomic analysis reveals different functional subtypes among IDH-wildtype glioblastoma"

| **GB CL** | | |
|---|---|---|
| **Overexpressed differentially expressed protein (DEP)** | **Inhibitor** | **Supplier** |
| LEG3 (Galectin-3) | GB1107 | MedChemExpress |
| GPNMB (Glycoprotein NMB) | Glembatumumab Vedotin | MedChemExpress |
| H13 (Histone H1.3) | Vorinostat | Selleck Chemicals |

| **GB PN** | | |
|---|---|---|
| **Overexpressed DEP** | **Inhibitor** | **Supplier** |
| CENPV (Centromere protein V) | GSK-923295 | MedChemExpress |
| RBP1 (RalA-binding protein 1) | No inhibitor available | - |
| TYB10 (Thymosin Beta 10) | No inhibitor available | - |

| **GB MES** | | |
|---|---|---|
| **Overexpressed DEP** | **Inhibitor** | **Supplier** |
| RBP1 (RalA-binding protein 1) | No inhibitor available | - |
| S10AD (Protein S100-A13) | Amlexanox | Selleck Chemicals |
| MOES (Moesin) | NSC668394<br>Indirect inhibitor: Fasudil HCl | MedChemExpress<br>Selleck Chemicals |

**Supplementary Table 2:** Differentially expressed proteins, inhibitors and suppliers
