## Supplementary Table 4a for "Quantitative proteomic analysis reveals different functional subtypes among IDH-wildtype glioblastoma"

| **Supplementary Table 4a:** Differentially expressed proteins between GB PN and LGG | | | | | | |
| --- | --- | --- | --- | --- | --- | --- |
| **Protein** | **logFC** | **AveExpr** | **t** | **P.Value** | **adj.P.Val** | **B** |
| RBP1_HUMAN | 1.06629863 | 21.8238491 | 4.94457165 | 4.6393E-06 | 0.00645544 | 3.96663269 |
| CENPV_HUMAN | 1.03751866 | 20.3128315 | 3.69419162 | 0.00042033 | 0.0322065 | -0.0689902 |
| TYB10_HUMAN | 1.02487435 | 24.1218195 | 3.79870524 | 0.00029635 | 0.02681775 | 0.24111916 |
| H15_HUMAN | 1.02413733 | 23.3197271 | 4.02915511 | 0.00013452 | 0.0224536 | 0.94440634 |
| HMGB2_HUMAN | 0.99453991 | 22.785919 | 3.83445802 | 0.00026262 | 0.02621723 | 0.34849836 |
| FLNC_HUMAN | 0.95634114 | 25.3098693 | 3.42677061 | 0.0010013 | 0.04351329 | -0.8356413 |
| RS23_HUMAN | 0.81847001 | 21.7227685 | 3.74290293 | 0.0003574 | 0.02824017 | 0.07483552 |
| GGH_HUMAN | 0.76409545 | 21.8362546 | 3.87060537 | 0.00023227 | 0.02553477 | 0.45772062 |
| RS27_HUMAN | 0.72968772 | 20.996412 | 3.40012771 | 0.0010894 | 0.04398032 | -0.9098196 |
| TAGL2_HUMAN | 0.69762778 | 24.2074544 | 3.46816755 | 0.00087763 | 0.04220485 | -0.7195753 |
| LYAR_HUMAN | 0.68800212 | 18.4497773 | 3.74379572 | 0.00035633 | 0.02824017 | 0.0774832 |
| RBM28_HUMAN | 0.68516399 | 17.7210441 | 4.21684069 | 6.9396E-05 | 0.01746229 | 1.53585839 |
| RL19_HUMAN | 0.67081242 | 22.8290936 | 3.92105916 | 0.00019547 | 0.02410959 | 0.61126209 |
| RL38_HUMAN | 0.65938759 | 21.5391643 | 3.40357234 | 0.00107761 | 0.04397847 | -0.9002523 |
| RL37A_HUMAN | 0.65777144 | 19.1521268 | 3.60114591 | 0.00057102 | 0.0369272 | -0.3402047 |
| SC24D_HUMAN | 0.65526185 | 19.1511941 | 4.64948221 | 1.4243E-05 | 0.00828063 | 2.95676315 |
| RTL8C_HUMAN | 0.63495521 | 17.3909888 | 4.19815163 | 7.4176E-05 | 0.01746229 | 1.4762475 |
| NASP_HUMAN | 0.63472923 | 22.3870036 | 3.4763753 | 0.0008549 | 0.04156928 | -0.6964468 |
| IKIP_HUMAN | 0.6324988 | 19.2642561 | 4.18772391 | 7.698E-05 | 0.01746229 | 1.44305459 |
| RS26_HUMAN | 0.62535485 | 19.4049864 | 3.32092501 | 0.0013965 | 0.04938523 | -1.1278876 |
| UGDH_HUMAN | 0.62495464 | 21.1342944 | 4.07349906 | 0.00011521 | 0.02080799 | 1.08268362 |
| RL28_HUMAN | 0.62372336 | 22.8615836 | 4.00456635 | 0.00014652 | 0.0224536 | 0.86813176 |
| PDIA4_HUMAN | 0.61837631 | 24.6702207 | 3.89075562 | 0.00021684 | 0.02503136 | 0.51889054 |
| LDHA_HUMAN | 0.61652767 | 24.9062762 | 3.53225626 | 0.00071429 | 0.03802253 | -0.5379693 |
| RL34_HUMAN | 0.60104592 | 22.6972421 | 3.51609495 | 0.00075252 | 0.03843934 | -0.5839829 |
| RS6_HUMAN | 0.59891844 | 22.906244 | 3.40334942 | 0.00107837 | 0.04397847 | -0.9008716 |
| AN32B_HUMAN | 0.56939029 | 21.5951791 | 3.79283239 | 0.00030228 | 0.02681775 | 0.22354318 |
| NDUA5_HUMAN | 0.55955945 | 22.3125761 | 3.42237943 | 0.00101534 | 0.04351329 | -0.8478953 |
| TIF1B_HUMAN | 0.54458356 | 23.8269077 | 3.9801185 | 0.00015948 | 0.02304245 | 0.79258053 |
| DNJC8_HUMAN | 0.53291349 | 22.1189814 | 3.82457666 | 0.00027156 | 0.02621723 | 0.31875581 |
| ERF3B_HUMAN | 0.52899258 | 19.5725421 | 4.85635277 | 6.5088E-06 | 0.00645544 | 3.66157283 |
| PIHD1_HUMAN | 0.52898086 | 17.2699172 | 4.09730625 | 0.00010598 | 0.01992056 | 1.1573003 |
| RL26_HUMAN | 0.52454843 | 20.9616081 | 3.36747508 | 0.00120737 | 0.0466877 | -1.0001677 |
| DGKB_HUMAN | 0.52225541 | 20.6008286 | 3.63266992 | 0.00051498 | 0.03519279 | -0.2488396 |
| XPO5_HUMAN | 0.52173268 | 18.1886139 | 3.55217589 | 0.00066969 | 0.0372563 | -0.4810548 |
| RL22_HUMAN | 0.51939928 | 21.8500631 | 3.65323114 | 0.0004813 | 0.03428044 | -0.1889577 |
| RL9_HUMAN | 0.51517869 | 22.2326277 | 3.60073241 | 0.00057179 | 0.0369272 | -0.3413996 |
| DDX5_HUMAN | 0.51454527 | 23.5715278 | 4.16066786 | 8.4741E-05 | 0.01746229 | 1.3571585 |
| EI2BA_HUMAN | 0.51047836 | 21.6756524 | 3.64197898 | 0.00049946 | 0.03472968 | -0.2217564 |
| PXL2B_HUMAN | 0.50874556 | 20.4258639 | 3.51167583 | 0.00076331 | 0.03860048 | -0.5965393 |
| SMC2_HUMAN | 0.50691212 | 19.846373 | 3.33465971 | 0.00133798 | 0.04832963 | -1.0903366 |
| OLA1_HUMAN | 0.50453522 | 23.7619026 | 3.93022284 | 0.00018942 | 0.02410959 | 0.63928403 |
| SCYL2_HUMAN | 0.49910479 | 20.1463363 | 4.45449707 | 2.9355E-05 | 0.00927802 | 2.30698909 |
| XRCC1_HUMAN | 0.49596848 | 19.7197643 | 3.79942822 | 0.00029563 | 0.02681775 | 0.24328409 |
| PSME3_HUMAN | 0.49114877 | 20.7562091 | 3.92978349 | 0.0001897 | 0.02410959 | 0.6379396 |
| RL27_HUMAN | 0.48292209 | 21.7038646 | 3.45394529 | 0.00091839 | 0.04244162 | -0.7595613 |
| AKT1_HUMAN | 0.47485379 | 20.1685661 | 3.55183895 | 0.00067042 | 0.0372563 | -0.4820193 |
| SYPM_HUMAN | 0.46564698 | 17.5944968 | 3.33835442 | 0.00132263 | 0.0481191 | -1.0802161 |
| RRP5_HUMAN | 0.46304904 | 18.0318879 | 3.35196823 | 0.00126751 | 0.04748013 | -1.0428559 |
| RS8_HUMAN | 0.46186658 | 23.0415407 | 3.45925354 | 0.00090297 | 0.04228092 | -0.7446506 |
| RL6_HUMAN | 0.46032906 | 23.5892677 | 3.36693139 | 0.00120943 | 0.0466877 | -1.0016668 |
| KIFA3_HUMAN | 0.45970449 | 18.9477247 | 3.44232848 | 0.00095301 | 0.0426491 | -0.7921363 |
| RS7_HUMAN | 0.45930885 | 23.3293253 | 3.46273212 | 0.000893 | 0.04220485 | -0.7348707 |
| UBFD1_HUMAN | 0.45532708 | 20.6896747 | 3.31781344 | 0.00141009 | 0.04951947 | -1.1363791 |
| YTHD2_HUMAN | 0.45386938 | 19.260484 | 4.00468163 | 0.00014646 | 0.0224536 | 0.86848869 |
| PLGT3_HUMAN | 0.45293273 | 19.0726756 | 3.40962284 | 0.0010572 | 0.04397847 | -0.8834306 |
| RL14_HUMAN | 0.44746571 | 22.161616 | 3.42896645 | 0.00099434 | 0.04351329 | -0.8295094 |
| PPID_HUMAN | 0.4457125 | 21.234006 | 3.64083915 | 0.00050134 | 0.03472968 | -0.2250751 |
| RL12_HUMAN | 0.44428193 | 22.7630746 | 3.46368292 | 0.0008903 | 0.04220485 | -0.7321964 |
| TOP2B_HUMAN | 0.42967185 | 22.7934193 | 3.3285076 | 0.0013639 | 0.0487767 | -1.1071704 |
| SAE1_HUMAN | 0.42663836 | 22.3011829 | 3.44743255 | 0.00093765 | 0.04244162 | -0.7778334 |
| TMX3_HUMAN | 0.4161553 | 20.7359795 | 3.54394105 | 0.0006878 | 0.03751964 | -0.5046102 |
| RS3A_HUMAN | 0.41360809 | 24.0225472 | 3.96442661 | 0.00016837 | 0.02305355 | 0.74423943 |
| STRAP_HUMAN | 0.4045587 | 23.3571575 | 4.32681559 | 4.6744E-05 | 0.013905 | 1.88972777 |
| ERF3A_HUMAN | 0.39978359 | 21.7792447 | 3.447434 | 0.00093764 | 0.04244162 | -0.7778293 |
| RUVB1_HUMAN | 0.39729484 | 23.3593965 | 3.52172926 | 0.00073898 | 0.03813263 | -0.5679578 |
| NUP62_HUMAN | 0.39680995 | 19.6459223 | 3.59417099 | 0.00058418 | 0.0369272 | -0.3603468 |
| MYH10_HUMAN | 0.3892748 | 25.606161 | 4.16036389 | 8.4832E-05 | 0.01746229 | 1.35619535 |
| NUDC_HUMAN | 0.38875244 | 23.3326502 | 3.41613834 | 0.00103562 | 0.04397847 | -0.8652925 |
| TR150_HUMAN | 0.38733037 | 22.5734242 | 3.34123771 | 0.00131077 | 0.04807099 | -1.0723127 |
| SYAP1_HUMAN | 0.38289379 | 19.8363071 | 3.57238394 | 0.00062716 | 0.0372563 | -0.4230914 |
| NU153_HUMAN | 0.37761531 | 20.4527691 | 3.4939516 | 0.00080807 | 0.04045928 | -0.6467903 |
| TXLNA_HUMAN | 0.35295193 | 21.1136584 | 3.57154312 | 0.00062887 | 0.0372563 | -0.4255076 |
| ADPPT_HUMAN | 0.34506102 | 21.8314842 | 3.56214074 | 0.00064838 | 0.0372563 | -0.4525005 |
| CPIN1_HUMAN | 0.33412048 | 21.5428284 | 3.38391837 | 0.00114653 | 0.0456537 | -0.9547477 |
| CSK22_HUMAN | 0.3233253 | 21.3180598 | 3.75055114 | 0.00034837 | 0.02824017 | 0.09753074 |
| XPO2_HUMAN | 0.31844488 | 23.2260758 | 3.52361036 | 0.0007345 | 0.03813263 | -0.5626036 |
| ELOC_HUMAN | 0.31573515 | 22.2761692 | 3.83524427 | 0.00026193 | 0.02621723 | 0.35086706 |
| RAE1L_HUMAN | 0.30476267 | 21.4608471 | 3.35257914 | 0.00126509 | 0.04748013 | -1.0411768 |
| PRS6B_HUMAN | 0.29296987 | 23.5218036 | 3.47852801 | 0.00084902 | 0.04156928 | -0.6903744 |
| AEDO_HUMAN | -0.3038263 | 21.1041364 | -3.5676755 | 0.00063683 | 0.0372563 | -0.4366169 |
| PP1R7_HUMAN | -0.3237208 | 23.3778373 | -3.9638996 | 0.00016867 | 0.02305355 | 0.74261782 |
| TBC17_HUMAN | -0.3248017 | 21.6700258 | -3.3746768 | 0.00118036 | 0.0462719 | -0.9802945 |
| MAVS_HUMAN | -0.3769532 | 19.792478 | -3.4248351 | 0.00100746 | 0.04351329 | -0.8410438 |
| NAGA_HUMAN | -0.3912411 | 19.0216285 | -4.4628634 | 2.8467E-05 | 0.00927802 | 2.33456326 |
| BCR_HUMAN | -0.4087662 | 21.517929 | -3.5798474 | 0.0006121 | 0.0372563 | -0.4016267 |
| PRDX3_HUMAN | -0.4266066 | 23.492456 | -3.8297506 | 0.00026685 | 0.02621723 | 0.33432291 |
| DIP2C_HUMAN | -0.4297833 | 19.9916518 | -3.5537344 | 0.00066631 | 0.0372563 | -0.4765924 |
| ISCA2_HUMAN | -0.4371763 | 19.1533537 | -3.4466536 | 0.00093998 | 0.04244162 | -0.7800171 |
| KAD3_HUMAN | -0.4460977 | 23.4672288 | -3.4351926 | 0.00097487 | 0.04324511 | -0.8121078 |
| PCCA_HUMAN | -0.4462022 | 22.6587456 | -3.9248503 | 0.00019294 | 0.02410959 | 0.6228501 |
| FN3K_HUMAN | -0.449128 | 22.7899476 | -4.246421 | 6.2431E-05 | 0.01661652 | 1.63052332 |
| CIA2B_HUMAN | -0.4521412 | 19.0717356 | -3.3521263 | 0.00126689 | 0.04748013 | -1.0424213 |
| CPT1A_HUMAN | -0.4662798 | 21.6067761 | -3.5240589 | 0.00073344 | 0.03813263 | -0.5613266 |
| MTCH1_HUMAN | -0.4687806 | 21.7851445 | -3.5715481 | 0.00062886 | 0.0372563 | -0.4254933 |
| MICU1_HUMAN | -0.4764536 | 18.7167242 | -3.4078817 | 0.00106303 | 0.04397847 | -0.8882735 |
| SAT2_HUMAN | -0.4888197 | 19.1211098 | -3.6876335 | 0.00042957 | 0.03224294 | -0.088258 |
| PCCB_HUMAN | -0.4936923 | 22.8452437 | -4.1554314 | 8.6327E-05 | 0.01746229 | 1.34057218 |
| CDS2_HUMAN | -0.4940321 | 20.4547827 | -3.5330729 | 0.0007124 | 0.03802253 | -0.5356404 |
| RHG12_HUMAN | -0.494214 | 20.0507928 | -5.8771331 | 1.1212E-07 | 0.00056701 | 7.32689561 |
| SPTCS_HUMAN | -0.4970796 | 17.6702787 | -3.3982699 | 0.00109581 | 0.04398032 | -0.9149766 |
| THTM_HUMAN | -0.5015065 | 22.7335259 | -3.8453037 | 0.00025314 | 0.02621723 | 0.38120037 |
| PTGR3_HUMAN | -0.5032076 | 20.5047612 | -4.641965 | 1.465E-05 | 0.00828063 | 2.9314432 |
| KITM_HUMAN | -0.5052948 | 20.3972292 | -3.7715912 | 0.00032465 | 0.02736295 | 0.16012182 |
| ALR_HUMAN | -0.5082401 | 17.8299251 | -3.3271588 | 0.00136964 | 0.0487767 | -1.1108581 |
| DUT_HUMAN | -0.5083465 | 21.9693001 | -4.3003182 | 5.1438E-05 | 0.01445116 | 1.80398914 |
| COQ8A_HUMAN | -0.5113889 | 19.8987929 | -3.5429529 | 0.00069 | 0.03751964 | -0.5074342 |
| MVD1_HUMAN | -0.5138142 | 21.3193267 | -3.7766645 | 0.00031917 | 0.02735666 | 0.17524822 |
| HBAP1_HUMAN | -0.5147487 | 16.4662698 | -3.779918 | 0.0003157 | 0.02735666 | 0.18495584 |
| ACS2L_HUMAN | -0.5281079 | 21.3338424 | -3.8894734 | 0.00021779 | 0.02503136 | 0.51499213 |
| KT3K_HUMAN | -0.5296527 | 22.8728097 | -3.3466478 | 0.00128879 | 0.04792223 | -1.0574698 |
| SYVN1_HUMAN | -0.5428708 | 19.5456331 | -4.0962834 | 0.00010636 | 0.01992056 | 1.15408898 |
| ACBD5_HUMAN | -0.5490867 | 20.8937427 | -4.5261227 | 2.2546E-05 | 0.00880474 | 2.5439593 |
| AP5B1_HUMAN | -0.5499438 | 16.6051674 | -3.594449 | 0.00058365 | 0.0369272 | -0.3595446 |
| MBOA2_HUMAN | -0.5564526 | 17.8855624 | -3.4878472 | 0.00082405 | 0.04085499 | -0.6640562 |
| MOCS1_HUMAN | -0.5886112 | 18.3018596 | -4.57435 | 1.8853E-05 | 0.00866744 | 2.70465203 |
| GIMA1_HUMAN | -0.6067866 | 18.9777867 | -3.7314922 | 0.00037128 | 0.02888567 | 0.04103209 |
| TINAL_HUMAN | -0.6093074 | 20.1282214 | -3.404421 | 0.00107473 | 0.04397847 | -0.8978941 |
| IGS21_HUMAN | -0.6159384 | 20.3938459 | -3.5960119 | 0.00058068 | 0.0369272 | -0.3550333 |
| RT06_HUMAN | -0.6164106 | 19.4855993 | -3.9837012 | 0.00015751 | 0.02304245 | 0.80363425 |
| MPV17_HUMAN | -0.619161 | 20.3120953 | -3.5718857 | 0.00062817 | 0.0372563 | -0.4245231 |
| K1671_HUMAN | -0.6258964 | 19.3902024 | -4.0589523 | 0.00012123 | 0.02113998 | 1.03722093 |
| MINY1_HUMAN | -0.6428593 | 18.7405402 | -3.3409861 | 0.0013118 | 0.04807099 | -1.0730025 |
| JMY_HUMAN | -0.6433924 | 18.46726 | -3.8724857 | 0.00023079 | 0.02553477 | 0.46342027 |
| SCMC2_HUMAN | -0.6448827 | 19.8885181 | -3.7930574 | 0.00030205 | 0.02681775 | 0.22421614 |
| TMLH_HUMAN | -0.6622907 | 20.0617199 | -4.6121884 | 1.6375E-05 | 0.00828063 | 2.83135483 |
| TXTP_HUMAN | -0.6673553 | 22.228104 | -3.9041637 | 0.00020712 | 0.02493859 | 0.55970507 |
| GBG4_HUMAN | -0.7047842 | 18.9326108 | -3.6012423 | 0.00057084 | 0.0369272 | -0.3399261 |
| THTR_HUMAN | -0.7110007 | 21.9781555 | -4.8638427 | 6.3251E-06 | 0.00645544 | 3.68737178 |
| CDIPT_HUMAN | -0.7205232 | 18.7998176 | -3.6848441 | 0.00043356 | 0.03224294 | -0.0964465 |
| MITOK_HUMAN | -0.7298916 | 18.7007069 | -4.5250636 | 2.2634E-05 | 0.00880474 | 2.54044055 |
| SERC1_HUMAN | -0.7399721 | 18.6646444 | -3.3552815 | 0.00125443 | 0.04748013 | -1.0337469 |
| ACOD_HUMAN | -0.7457719 | 17.5450739 | -3.7504364 | 0.00034851 | 0.02824017 | 0.09718991 |
| SLIK2_HUMAN | -0.7490408 | 18.4968248 | -3.5525012 | 0.00066898 | 0.0372563 | -0.4801234 |
| F241B_HUMAN | -0.8113322 | 18.9971091 | -3.3785071 | 0.00116623 | 0.04607506 | -0.9697122 |
| DJC30_HUMAN | -0.8123685 | 17.4312145 | -3.6614041 | 0.0004685 | 0.03384551 | -0.1650918 |
| DHB8_HUMAN | -0.8308712 | 21.0509211 | -5.1696895 | 1.9325E-06 | 0.00488631 | 4.75633716 |
| GHC2_HUMAN | -0.8617913 | 17.9966741 | -4.0118122 | 0.00014288 | 0.0224536 | 0.8905785 |
| SYVM_HUMAN | -0.8721812 | 17.4619009 | -3.8295072 | 0.00026707 | 0.02621723 | 0.33359037 |
| LRP4_HUMAN | -0.8859938 | 18.559531 | -4.458026 | 2.8977E-05 | 0.00927802 | 2.31861637 |
| GSTT1_HUMAN | -0.8941856 | 19.7467145 | -3.8211025 | 0.00027477 | 0.02621723 | 0.30831037 |
| SE6L1_HUMAN | -0.9028831 | 20.5832654 | -4.6214929 | 1.5815E-05 | 0.00828063 | 2.86259423 |
| AIG1_HUMAN | -1.3885222 | 17.7129956 | -4.8136957 | 7.6592E-06 | 0.00645544 | 3.51500586 |
| REN3A_HUMAN | -1.4972617 | 15.0499783 | -3.6620475 | 0.0004675 | 0.03384551 | -0.1632117 |
