## Supplementary Table 4b for "Quantitative proteomic analysis reveals different functional subtypes among IDH-wildtype glioblastoma"

| **Supplementary Table 4b:** Differentially expressed proteins with the highest and lowest log2FC values between GB PN and LGG | | | | | | | | |
| --- | --- | --- | --- | --- | --- | --- | --- | --- |
| **Proteins with highest log2FC values in GB PN (compared to LGG):** | | | | | | | | |
| **Protein Name** | **Protein** | **Gene** | **Locus** | **Function** | **Pathway** | **Glioma** | **Biological process** | **Molecular function** |
| RBP1_HUMAN | RalA-binding protein 1 | RALBP1 | 18p11 | Multifunctional protein that functions as a downstream effector of RALA and RALB | Signaling by Rho GTPases, RAC1 GTPase cycle, Doxorubicin Pathway, Pharmacokinetics, GPCR Pathway, ERK Signaling | Expression of RET1 correlated with tumor proliferation. It was shown to be an independent prognostic marker for adverse patient survival in glioma. | Positive regulation of protein phosphorylation, Endocytosis, Chemotaxis, Signal transduction, Small GTPase mediated signal transduction | Nucleotide binding, Protein binding, GTPase activator activity, ATP binding, ABC-type xenobiotic transporter activity |
| CENPV_HUMAN | Centromere protein V | CENPV | 17p11 | Required for distribution of pericentromeric heterochromatin in interphase nuclei and for centromere formation and organization, chromosome alignment and cytokinesis. | n/a | no results on pubmed | Ameboidal-type cell migration, Cell cycle, Pericentric heterochromatin formation, Positive regulation of cytokinesis, Regulation of chromosome organization | Molecular function, Protein binding, Carbon-sulfur lyase activity, Metal ion binding |
| TYB10_HUMAN | Thymosin Beta 10 | TMSB10 | 2p11 | Involved in organisation of the cytoskeleton and regulation of cell migration. | Extrafollicular and follicular B cell activation by SARS-CoV-2, VEGFA-VEGFR2 signaling | TMSB10 may play a key role in the malignant progression of glioma, in the promotion of macrophage infiltration and immunosuppressive polarization | Actin filament organization, Regulation of cell migration, Sequestering of actin monomers | Actin binding, Actin monomer binding, Protein binding |
| **Proteins with lowest log2FC values in GB PN (compared to LGG):** | | | | | | | | |
| **Protein Name** | **Protein** | **Gene** | **Locus** | **Function** | **Pathway** | **Glioma** | **Biological process** | **Molecular function** |
| SE6L1_HUMAN | Seizure 6-like protein | SEZ6L | 22q12 | May contribute to specialized endoplasmic reticulum functions in neurons. | n/a | no results on pubmed | Synapse maturation, Regulation of protein kinase C signaling | Protein binding |
| AIG1_HUMAN | Androgen-induced gene 1 protein | AIG1 | 6q24 | Hydrolyzes bioactive fatty-acid esters of hydroxy-fatty acids (FAHFAs), but not other major classes of lipids | n/a | no results on pubmed | Lipid metabolism | Protein binding, enables hydrolase activity |
| REN3A_HUMAN | Regulator of nonsense transcripts 3A | UPF3A | 13q34 | Involved in nonsense-mediated decay (NMD) of mRNAs containing premature stop codons by associating with the nuclear exon junction complex (EJC) and serving as link between the EJC core and NMD machinery. | Nervous system development, Peptide chain elongation, Regulation of expression of SLITs and ROBOs | no results on pubmed | mRNA transport, Nonsense-mediated mRNA decay, Transport | RNA binding, Nucleid acid binding |
