## Supplementary Table 5 for "Quantitative proteomic analysis reveals different functional subtypes among IDH-wildtype glioblastoma"

| **Supplementary Table 5:** Over representation analysis (ORA) using Gene ontology (GO) enrichment and Kyoto Encyclopedia of Genes and Genomes (KEGG) Pathway analysis showing the pathway and functions with highest numbers of over- and underexpressed proteins in the four HGG subgroups (compared to LGG). | | | | | | | | |
| --- | --- | --- | --- | --- | --- | --- | --- | --- |
| **Proteins overexpressed in GB PN (compared to LGG)** | | |  |  |  |  |  |  |
| GO biological processes: | |  |  |  |  |  |  |  |
| ID | Description | GeneRatio | BgRatio | pvalue | p.adjust | qvalue | geneID | Count |
| GO:0002181 | cytoplasmic translation | 20/77 | 130/4804 | 4.098E-15 | 6.6797E-12 | 6.40145E-12 | RS7/RS3A/RL6/RL12/RL14/RL19/RS6/RL27/RS23/RL38/RL34/RL28/RL22/RS8/RS27/RL26/YTHD2/RL9/RS26/RL37A | 20 |
| GO:0042254 | ribosome biogenesis | 15/77 | 130/4804 | 1.1548E-09 | 7.0066E-07 | 6.71477E-07 | RS7/RS3A/RL6/RL14/RS6/RL27/RS23/RL38/LYAR/RRP5/RS8/RS27/RL26/YTHD2/PIHD1 | 15 |
| GO:0006412 | translation | 25/77 | 405/4804 | 1.2896E-09 | 7.0066E-07 | 6.71477E-07 | EI2BA/RS7/RS3A/RL6/RL12/RL14/RL19/RS6/ERF3B/RL27/RS23/ERF3A/RL38/SYPM/RL34/RL28/RL22/RS8/RS27/RL26/YTHD2/AKT1/RL9/RS26/RL37A | 25 |
| GO:0043043 | peptide biosynthetic process | 25/77 | 416/4804 | 2.271E-09 | 9.2544E-07 | 8.86889E-07 | EI2BA/RS7/RS3A/RL6/RL12/RL14/RL19/RS6/ERF3B/RL27/RS23/ERF3A/RL38/SYPM/RL34/RL28/RL22/RS8/RS27/RL26/YTHD2/AKT1/RL9/RS26/RL37A | 25 |
| GO:0043604 | amide biosynthetic process | 25/77 | 468/4804 | 2.5952E-08 | 8.3787E-06 | 8.02974E-06 | EI2BA/RS7/RS3A/RL6/RL12/RL14/RL19/RS6/ERF3B/RL27/RS23/ERF3A/RL38/SYPM/RL34/RL28/RL22/RS8/RS27/RL26/YTHD2/AKT1/RL9/RS26/RL37A | 25 |
| GO:0006518 | peptide metabolic process | 25/77 | 472/4804 | 3.0842E-08 | 8.3787E-06 | 8.02974E-06 | EI2BA/RS7/RS3A/RL6/RL12/RL14/RL19/RS6/ERF3B/RL27/RS23/ERF3A/RL38/SYPM/RL34/RL28/RL22/RS8/RS27/RL26/YTHD2/AKT1/RL9/RS26/RL37A | 25 |
| GO:0022613 | ribonucleoprotein complex biogenesis | 17/77 | 223/4804 | 4.8123E-08 | 1.1206E-05 | 1.0739E-05 | RUVB1/RS7/RS3A/RL6/RL14/RS6/RL27/RS23/RL38/LYAR/STRAP/RRP5/RS8/RS27/RL26/YTHD2/PIHD1 | 17 |
| GO:0006364 | rRNA processing | 11/77 | 84/4804 | 6.2824E-08 | 1.28E-05 | 1.22673E-05 | RS7/RL14/RS6/RL27/LYAR/RRP5/RS8/RS27/RL26/YTHD2/PIHD1 | 11 |
| GO:0034660 | ncRNA metabolic process | 16/77 | 213/4804 | 1.5725E-07 | 2.848E-05 | 2.72935E-05 | DDX5/RS7/XPO5/RL14/RS6/RL27/LYAR/SYPM/ELOC/RRP5/RS8/RS27/RL26/YTHD2/AKT1/PIHD1 | 16 |
| GO:0016072 | rRNA metabolic process | 11/77 | 98/4804 | 3.168E-07 | 5.1638E-05 | 4.94874E-05 | RS7/RL14/RS6/RL27/LYAR/RRP5/RS8/RS27/RL26/YTHD2/PIHD1 | 11 |
| GO:0034470 | ncRNA processing | 13/77 | 145/4804 | 3.5165E-07 | 5.2108E-05 | 4.99373E-05 | DDX5/RS7/RL14/RS6/RL27/LYAR/RRP5/RS8/RS27/RL26/YTHD2/AKT1/PIHD1 | 13 |
| GO:0042274 | ribosomal small subunit biogenesis | 7/77 | 44/4804 | 4.9117E-06 | 0.00066717 | 0.000639378 | RS7/RS3A/RS6/RS23/RL38/RS8/RS27 | 7 |
| GO:0051276 | chromosome organization | 12/77 | 207/4804 | 9.2083E-05 | 0.01154576 | 0.011064838 | RUVB1/NASP/NUP62/TIF1B/TOP2B/NUDC/HMGB2/SMC2/H15/CENPV/RAE1L/XRCC1 | 12 |
| GO:0006396 | RNA processing | 17/77 | 409/4804 | 0.00018991 | 0.02211101 | 0.021190018 | DDX5/RS7/TR150/RL14/RS6/RBM28/RL27/LYAR/STRAP/RRP5/RS8/RS27/RL26/YTHD2/AKT1/RS26/PIHD1 | 17 |

| GO molecular functions: | |  |  |  |  |  |  |  |
| --- | --- | --- | --- | --- | --- | --- | --- | --- |
| ID | Description | GeneRatio | BgRatio | pvalue | p.adjust | qvalue | geneID | Count |
| GO:0003735 | structural constituent of ribosome | 19/79 | 113/4871 | 5.2828E-15 | 1.4211E-12 | 1.3791E-12 | RS7/RS3A/RL6/RL12/RL14/RL19/RS6/RL27/RS23/RL38/RL34/RL28/RL22/RS8/RS27/RL26/RL9/RS26/RL37A | 19 |
| GO:0005198 | structural molecule activity | 22/79 | 341/4871 | 9.8789E-09 | 1.3287E-06 | 1.2895E-06 | RS7/NUP62/RS3A/RL6/RL12/RL14/RL19/RS6/RL27/RS23/H15/NU153/RL38/RL34/RL28/RL22/RS8/RS27/RL26/RL9/RS26/RL37A | 22 |
| GO:0048027 | mRNA 5'-UTR binding | 4/79 | 18/4871 | 0.00016503 | 0.01479775 | 0.01436057 | MYH10/RS7/RS3A/RL26 | 4 |

| KEGG pathway analysis: | |  |  |  |  |  |  |  |  |
| --- | --- | --- | --- | --- | --- | --- | --- | --- | --- |
| ID | Description | GeneRatio | BgRatio | pvalue | p.adjust | qvalue | geneID | Count | core_enrichment |
| hsa03010 | Ribosome | 19/51 | 101/2742 | 1.9117E-15 | 2.8484E-13 | 2.8484E-13 | P62081/P61247/Q02878/P30050/P50914/P84098/P62753/P61353/P62266/P63173/P49207/P46779/P35268/P62241/P42677/P61254/P32969/P62854/P61513 | 19 | RS7/RS3A/RL6/RL12/RL14/RL19/RS6/RL27/RS23/RL38/RL34/RL28/RL22/RS8/RS27/RL26/RL9/RS26/RL37A |
| hsa05171 | Coronavirus disease - COVID-19 | 19/51 | 140/2742 | 1.0198E-12 | 7.5978E-11 | 7.5978E-11 | P62081/P61247/Q02878/P30050/P50914/P84098/P62753/P61353/P62266/P63173/P49207/P46779/P35268/P62241/P42677/P61254/P32969/P62854/P61513 | 19 | RS7/RS3A/RL6/RL12/RL14/RL19/RS6/RL27/RS23/RL38/RL34/RL28/RL22/RS8/RS27/RL26/RL9/RS26/RL37A |

| **Proteins underexpressed in GB PN (compared to LGG)** | | | |  |  |  |  |  |
| --- | --- | --- | --- | --- | --- | --- | --- | --- |
| GO biological processes: | |  |  |  |  |  |  |  |
| ID | Description | GeneRatio | BgRatio | pvalue | p.adjust | qvalue | geneID | Count |
| GO:0044255 | cellular lipid metabolic process | 14/60 | 331/4804 | 3.9074E-05 | 0.04591187 | 0.04491446 | DHB8/CDS2/CPT1A/ACS2L/PCCB/PCCA/AIG1/PTGR3/ACBD5/CDIPT/MVD1/SERC1/ACOD/MBOA2 | 14 |
| GO:0006631 | fatty acid metabolic process | 9/60 | 154/4804 | 0.00010316 | 0.04983846 | 0.04875574 | DHB8/CPT1A/ACS2L/PCCB/PCCA/AIG1/PTGR3/ACBD5/ACOD | 9 |
| GO:0090407 | organophosphate biosynthetic process | 12/60 | 278/4804 | 0.00012725 | 0.04983846 | 0.04875574 | CDS2/TXTP/KITM/DJC30/ACS2L/DUT/CDIPT/KAD3/MVD1/SERC1/MOCS1/MBOA2 | 12 |

| **Proteins overexpressed in GB CL (compared to LGG)** | | |  |  |  |  |  |  |
| --- | --- | --- | --- | --- | --- | --- | --- | --- |
| GO biological processes: | |  |  |  |  |  |  |  |
| ID | Description | GeneRatio | BgRatio | pvalue | p.adjust | qvalue | geneID | Count |
| GO:0034975 | protein folding in endoplasmic reticulum | 4/86 | 9/4804 | 1.127E-05 | 0.02536847 | 0.02379722 | ENPL/CALR/BIP/PDIA1 | 4 |

| GO molecular functions: | |  |  |  |  |  |  |  |
| --- | --- | --- | --- | --- | --- | --- | --- | --- |
| ID | Description | GeneRatio | BgRatio | pvalue | p.adjust | qvalue | geneID | Count |
| GO:0050839 | cell adhesion molecule binding | 21/89 | 363/4871 | 1.368E-06 | 0.00051028 | 0.00046082 | MACF1/ACTN4/MOES/RUVB1/MYH9/LDHA/CLIC1/CAPG/IQGA1/EGFR/CALR/CALD1/PRDX1/SWP70/BIP/EP15R/CNN3/PDIA1/TAGL2/FBP1L/GPNMB | 21 |
| GO:0045296 | cadherin binding | 16/89 | 257/4871 | 1.2323E-05 | 0.00229828 | 0.00207549 | MACF1/RUVB1/MYH9/LDHA/CLIC1/CAPG/IQGA1/EGFR/CALD1/PRDX1/SWP70/BIP/EP15R/CNN3/TAGL2/FBP1L | 16 |
| GO:0005509 | calcium ion binding | 14/89 | 214/4871 | 2.6623E-05 | 0.00331009 | 0.00298922 | MACF1/ACTN4/CAB45/ANXA5/MYL6/ENPL/IQGA1/GLU2B/CALR/SWP70/BIP/EP15R/RCN2/CALU | 14 |
| GO:0003677 | DNA binding | 17/89 | 359/4871 | 0.00021101 | 0.01921453 | 0.01735191 | ACTN4/ANM1/EGFR/TIF1B/CALR/TIF1A/SWP70/HMGB2/H13/H15/HMGN2/NU153/PDCD5/LYAR/XRCC1/MYT1L/HDAC1 | 17 |
| GO:0003779 | actin binding | 13/89 | 235/4871 | 0.00029235 | 0.01921453 | 0.01735191 | MACF1/ACTN4/MOES/MYH9/CAPG/IQGA1/EGFR/CALD1/CNN3/PDIA1/TAGL2/FHL3/SSH3 | 13 |
| GO:0042056 | chemoattractant activity | 3/89 | 8/4871 | 0.00030908 | 0.01921453 | 0.01735191 | HMGB2/LEG3/GPNMB | 3 |
| GO:0031490 | chromatin DNA binding | 5/89 | 41/4871 | 0.00081155 | 0.04324378 | 0.03905182 | ACTN4/H13/H15/HMGN2/HDAC1 | 5 |

| KEGG pathway analysis: | |  |  |  |  |  |  |  |  |  |  |
| --- | --- | --- | --- | --- | --- | --- | --- | --- | --- | --- | --- |
| category | subcategory | ID | Description | GeneRatio | BgRatio | pvalue | p.adjust | qvalue | geneID | Count | core_enrichment |
| Organismal Systems | Endocrine system | hsa04922 | Glucagon signaling pathway | 6/51 | 50/2742 | 0.0003 | 0.0413773 | 0.0354 | Q99873/P06737/P00338/P15259/Q13557/P31749 | 6 | ANM1/PYGL/LDHA/PGAM2/KCC2D/AKT1 |
| Human Diseases | Cancer: overview | hsa05230 | Central carbon metabolism in cancer | 5/51 | 36/2742 | 0.0004 | 0.0413773 | 0.0354 | P00338/P00533/P15259/P36507/P31749 | 5 | LDHA/EGFR/PGAM2/MP2K2/AKT1 |
| Metabolism | Metabolism of cofactors and vitamins | hsa00770 | Pantothenate and CoA biosynthesis | 3/51 | 10/2742 | 0.0007 | 0.0413773 | 0.0354 | P54687/Q12882/Q9HAB8 | 3 | BCAT1/DPYD/PPCS |

| **Proteins underexpressed in GB CL (compared to LGG)** | | |  |  |  |  |  |  |
| --- | --- | --- | --- | --- | --- | --- | --- | --- |
| GO biological processes: | |  |  |  |  |  |  |  |
| ID | Description | GeneRatio | BgRatio | pvalue | p.adjust | qvalue | geneID | Count |
| GO:0090151 | establishment of protein localization to mitochondrial membrane | 5/70 | 28/4804 | 4.3076E-05 | 0.02295785 | 0.02215914 | MTCH1/TIM29/MTCH2/BCS1/MTX1 | 5 |
| GO:0007006 | mitochondrial membrane organization | 7/70 | 68/4804 | 4.9233E-05 | 0.02295785 | 0.02215914 | MTCH1/HIP1R/TIM29/MTCH2/BCS1/MTX1/DJC11 | 7 |
| GO:0051205 | protein insertion into membrane | 6/70 | 48/4804 | 5.8367E-05 | 0.02295785 | 0.02215914 | MTCH1/TIM29/MTCH2/BCS1/MTX1/EMC6 | 6 |
| GO:0090150 | establishment of protein localization to membrane | 9/70 | 144/4804 | 0.00020825 | 0.04453149 | 0.04298223 | MTCH1/TIM29/MTCH2/BCS1/MTX1/VAMP3/EMC6/ATAD1/ZDH14 | 9 |
| GO:0032787 | monocarboxylic acid metabolic process | 12/70 | 260/4804 | 0.00031172 | 0.04453149 | 0.04298223 | DHB8/PYC/CPT1A/DCMC/ALDOC/ACSF3/GABT/ABD12/PCCB/PCCA/PTGR3/ACBD5 | 12 |
| GO:0019752 | carboxylic acid metabolic process | 16/70 | 428/4804 | 0.00032225 | 0.04453149 | 0.04298223 | DHB8/PYC/CPT1A/DCMC/ALDOC/ACSF3/GABT/THTR/ABD12/ACY1/PCCB/PCCA/PTGR3/SYEM/ACBD5/SYVM | 16 |
| GO:0006631 | fatty acid metabolic process | 9/70 | 154/4804 | 0.00034511 | 0.04453149 | 0.04298223 | DHB8/CPT1A/DCMC/ACSF3/ABD12/PCCB/PCCA/PTGR3/ACBD5 | 9 |
| GO:0006082 | organic acid metabolic process | 16/70 | 436/4804 | 0.00039784 | 0.04453149 | 0.04298223 | DHB8/PYC/CPT1A/DCMC/ALDOC/ACSF3/GABT/THTR/ABD12/ACY1/PCCB/PCCA/PTGR3/SYEM/ACBD5/SYVM | 16 |
| GO:0043436 | oxoacid metabolic process | 16/70 | 436/4804 | 0.00039784 | 0.04453149 | 0.04298223 | DHB8/PYC/CPT1A/DCMC/ALDOC/ACSF3/GABT/THTR/ABD12/ACY1/PCCB/PCCA/PTGR3/SYEM/ACBD5/SYVM | 16 |
| GO:0006790 | sulfur compound metabolic process | 8/70 | 125/4804 | 0.00040971 | 0.04453149 | 0.04298223 | DCMC/ACSF3/TXTP/THTR/KIME/CIA2B/B3GA3/ISCA2 | 8 |
| GO:0051204 | protein insertion into mitochondrial membrane | 4/70 | 25/4804 | 0.00041512 | 0.04453149 | 0.04298223 | MTCH1/TIM29/MTCH2/MTX1 | 4 |
| GO:0006839 | mitochondrial transport | 7/70 | 97/4804 | 0.00046683 | 0.04590455 | 0.04430752 | CPT1A/MTCH1/HIP1R/TIM29/MTCH2/BCS1/MTX1 | 7 |

| GO molecular functions: | |  |  |  |  |  |  |  |
| --- | --- | --- | --- | --- | --- | --- | --- | --- |
| ID | Description | GeneRatio | BgRatio | pvalue | p.adjust | qvalue | geneID | Count |
| GO:0016885 | ligase activity, forming carbon-carbon bonds | 3/69 | 6/4871 | 5.2792E-05 | 0.01277563 | 0.01277563 | PYC/PCCB/PCCA | 3 |

| KEGG pathway analysis: | |  |  |  |  |  |  |  |  |  |  |
| --- | --- | --- | --- | --- | --- | --- | --- | --- | --- | --- | --- |
| category | subcategory | ID | Description | GeneRatio | BgRatio | pvalue | p.adjust | qvalue | geneID | Count | core_enrichment |
| Metabolism | Carbohydrate metabolism | hsa00640 | Propanoate metabolism | 4/34 | 26/2742 | 0.00024309 | 0.01944739 | 0.01842384 | O95822/P80404/P05166/P05165 | 4 | DCMC/GABT/PCCB/PCCA |

| **Proteins overexpressed in GB MES (compared to LGG)** | |  |  |  |  |  |  |  |
| --- | --- | --- | --- | --- | --- | --- | --- | --- |
| GO biological processes: | |  |  |  |  |  |  |  |
| ID | Description | GeneRatio | BgRatio | pvalue | p.adjust | qvalue | geneID | Count |
| GO:0006120 | mitochondrial electron transport, NADH to ubiquinone | 3/14 | 35/4804 | 0.0001221 | 0.04538594 | 0.03590519 | NDUA5/NDUA8/NDUA7 | 3 |
| GO:0042776 | proton motive force-driven mitochondrial ATP synthesis | 3/14 | 50/4804 | 0.00035631 | 0.04538594 | 0.03590519 | NDUA5/NDUA8/NDUA7 | 3 |
| GO:0015986 | proton motive force-driven ATP synthesis | 3/14 | 53/4804 | 0.00042366 | 0.04538594 | 0.03590519 | NDUA5/NDUA8/NDUA7 | 3 |
| GO:0019646 | aerobic electron transport chain | 3/14 | 59/4804 | 0.00058188 | 0.04538594 | 0.03590519 | NDUA5/NDUA8/NDUA7 | 3 |
| GO:0006754 | ATP biosynthetic process | 3/14 | 61/4804 | 0.00064197 | 0.04538594 | 0.03590519 | NDUA5/NDUA8/NDUA7 | 3 |
| GO:0042773 | ATP synthesis coupled electron transport | 3/14 | 63/4804 | 0.0007059 | 0.04538594 | 0.03590519 | NDUA5/NDUA8/NDUA7 | 3 |
| GO:0042775 | mitochondrial ATP synthesis coupled electron transport | 3/14 | 63/4804 | 0.0007059 | 0.04538594 | 0.03590519 | NDUA5/NDUA8/NDUA7 | 3 |
| GO:0009206 | purine ribonucleoside triphosphate biosynthetic process | 3/14 | 66/4804 | 0.00080923 | 0.04538594 | 0.03590519 | NDUA5/NDUA8/NDUA7 | 3 |
| GO:0009145 | purine nucleoside triphosphate biosynthetic process | 3/14 | 67/4804 | 0.0008457 | 0.04538594 | 0.03590519 | NDUA5/NDUA8/NDUA7 | 3 |
| GO:0009201 | ribonucleoside triphosphate biosynthetic process | 3/14 | 70/4804 | 0.00096137 | 0.0464344 | 0.03673464 | NDUA5/NDUA8/NDUA7 | 3 |

| GO molecular functions: | |  |  |  |  |  |  |  |
| --- | --- | --- | --- | --- | --- | --- | --- | --- |
| ID | Description | GeneRatio | BgRatio | pvalue | p.adjust | qvalue | geneID | Count |
| GO:0008137 | NADH dehydrogenase (ubiquinone) activity | 3/14 | 30/4871 | 7.3331E-05 | 0.00190547 | 0.00132776 | NDUA5/NDUA8/NDUA7 | 3 |
| GO:0050136 | NADH dehydrogenase (quinone) activity | 3/14 | 31/4871 | 8.105E-05 | 0.00190547 | 0.00132776 | NDUA5/NDUA8/NDUA7 | 3 |
| GO:0003954 | NADH dehydrogenase activity | 3/14 | 32/4871 | 8.9283E-05 | 0.00190547 | 0.00132776 | NDUA5/NDUA8/NDUA7 | 3 |
| GO:0003955 | NAD(P)H dehydrogenase (quinone) activity | 3/14 | 34/4871 | 0.00010735 | 0.00190547 | 0.00132776 | NDUA5/NDUA8/NDUA7 | 3 |
| GO:0016655 | oxidoreductase activity, acting on NAD(P)H, quinone or similar compound as acceptor | 3/14 | 41/4871 | 0.00018898 | 0.00268345 | 0.00186986 | NDUA5/NDUA8/NDUA7 | 3 |
| GO:0015453 | oxidoreduction-driven active transmembrane transporter activity | 3/14 | 47/4871 | 0.00028453 | 0.003367 | 0.00234617 | NDUA5/NDUA8/NDUA7 | 3 |
| GO:0016651 | oxidoreductase activity, acting on NAD(P)H | 3/14 | 50/4871 | 0.00034218 | 0.00347073 | 0.00241844 | NDUA5/NDUA8/NDUA7 | 3 |
| GO:0009055 | electron transfer activity | 3/14 | 73/4871 | 0.00104418 | 0.0092671 | 0.00645743 | NDUA5/NDUA8/NDUA7 | 3 |
| GO:0015399 | primary active transmembrane transporter activity | 3/14 | 78/4871 | 0.00126638 | 0.00999035 | 0.0069614 | NDUA5/NDUA8/NDUA7 | 3 |
| GO:0022804 | active transmembrane transporter activity | 3/14 | 133/4871 | 0.0058097 | 0.0412489 | 0.02874275 | NDUA5/NDUA8/NDUA7 | 3 |

| KEGG pathway analysis: | |  |  |  |  |  |  |  |  |  |  |
| --- | --- | --- | --- | --- | --- | --- | --- | --- | --- | --- | --- |
| category | subcategory | ID | Description | GeneRatio | BgRatio | pvalue | p.adjust | qvalue | geneID | Count | core_enrichment |
| Human Diseases | Cardiovascular disease | hsa05415 | Diabetic cardiomyopathy | 4/7 | 123/2742 | 0.00012 | 0.00583675 | 0.00409596 | Q16718/Q13557/P51970/O95182 | 4 | NDUA5/KCC2D/NDUA8/NDUA7 |
| Human Diseases | Neurodegenerative disease | hsa05012 | Parkinson disease | 4/7 | 166/2742 | 0.00039 | 0.00883106 | 0.00619724 | Q16718/Q13557/P51970/O95182 | 4 | NDUA5/KCC2D/NDUA8/NDUA7 |
| Organismal Systems | Nervous system | hsa04723 | Retrograde endocannabinoid signaling | 3/7 | 80/2742 | 0.00077 | 0.00883106 | 0.00619724 | Q16718/P51970/O95182 | 3 | NDUA5/NDUA8/NDUA7 |
| Human Diseases | Endocrine and metabolic disease | hsa04932 | Non-alcoholic fatty liver disease | 3/7 | 84/2742 | 0.00089 | 0.00883106 | 0.00619724 | Q16718/P51970/O95182 | 3 | NDUA5/NDUA8/NDUA7 |
| Metabolism | Energy metabolism | hsa00190 | Oxidative phosphorylation | 3/7 | 85/2742 | 0.00092 | 0.00883106 | 0.00619724 | Q16718/P51970/O95182 | 3 | NDUA5/NDUA8/NDUA7 |
| Human Diseases | Neurodegenerative disease | hsa05022 | Pathways of neurodegeneration - multiple diseases | 4/7 | 261/2742 | 0.00223 | 0.01683404 | 0.01181336 | Q16718/Q13557/P51970/O95182 | 4 | NDUA5/KCC2D/NDUA8/NDUA7 |
| Organismal Systems | Environmental adaptation | hsa04714 | Thermogenesis | 3/7 | 119/2742 | 0.00245 | 0.01683404 | 0.01181336 | Q16718/P51970/O95182 | 3 | NDUA5/NDUA8/NDUA7 |
| Human Diseases | Cancer: overview | hsa05208 | Chemical carcinogenesis - reactive oxygen species | 3/7 | 127/2742 | 0.00296 | 0.01777285 | 0.01247218 | Q16718/P51970/O95182 | 3 | NDUA5/NDUA8/NDUA7 |
| Human Diseases | Neurodegenerative disease | hsa05020 | Prion disease | 3/7 | 178/2742 | 0.00775 | 0.04133779 | 0.02900898 | Q16718/P51970/O95182 | 3 | NDUA5/NDUA8/NDUA7 |
| Human Diseases | Neurodegenerative disease | hsa05016 | Huntington disease | 3/7 | 189/2742 | 0.00917 | 0.04402497 | 0.03089471 | Q16718/P51970/O95182 | 3 | NDUA5/NDUA8/NDUA7 |

| **Proteins underexpressed in IDH HGG (compared to LGG)** | | |  |  |  |  |  |  |
| --- | --- | --- | --- | --- | --- | --- | --- | --- |
| GO biological processes: | |  |  |  |  |  |  |  |
| ID | Description | GeneRatio | BgRatio | pvalue | p.adjust | qvalue | geneID | Count |
| GO:0000301 | retrograde transport, vesicle recycling within Golgi | 1/1 | 7/4804 | 0.00145712 | 0.00874271 | 0.00153381 | COG8 | 1 |
| GO:0006891 | intra-Golgi vesicle-mediated transport | 1/1 | 26/4804 | 0.00541216 | 0.01623647 | 0.002848503 | COG8 | 1 |
| GO:0070085 | glycosylation | 1/1 | 43/4804 | 0.00895087 | 0.01790175 | 0.003140658 | COG8 | 1 |
| GO:0007030 | Golgi organization | 1/1 | 83/4804 | 0.01727727 | 0.0259159 | 0.00454665 | COG8 | 1 |
| GO:0048193 | Golgi vesicle transport | 1/1 | 185/4804 | 0.03850958 | 0.04621149 | 0.008107279 | COG8 | 1 |
