## Supplementary Table 6a for "Quantitative proteomic analysis reveals different functional subtypes among IDH-wildtype glioblastoma"

| **Supplementary Table 6a:** Differentially expressed proteins between GB CL and LGG | | | | | | |
| --- | --- | --- | --- | --- | --- | --- |
| **Protein** | **logFC** | **AveExpr** | **t** | **P.Value** | **adj.P.Val** | **B** |
| H13_HUMAN | 1.31845448 | 21.142592 | 4.48132304 | 2.6599E-05 | 0.00987517 | 2.3987697 |
| GPNMB_HUMAN | 1.18501054 | 17.6545716 | 4.39282224 | 3.6783E-05 | 0.00987517 | 2.10699854 |
| LEG3_HUMAN | 1.08465239 | 22.6514627 | 4.82166016 | 7.4303E-06 | 0.00626254 | 3.54906291 |
| H15_HUMAN | 1.04457947 | 23.3197271 | 4.33594627 | 4.5225E-05 | 0.01039555 | 1.92119796 |
| PGAM2_HUMAN | 1.03546752 | 20.7854034 | 4.36977567 | 4.0002E-05 | 0.00987517 | 2.03154721 |
| TENA_HUMAN | 0.99734435 | 25.7381482 | 3.79543624 | 0.00029964 | 0.02405127 | 0.22833254 |
| EGFR_HUMAN | 0.97277054 | 24.1904915 | 3.36858391 | 0.00120317 | 0.04309593 | -1.0035106 |
| MOES_HUMAN | 0.95432348 | 25.6833345 | 5.34531903 | 9.6526E-07 | 0.00222221 | 5.39534808 |
| HSPB6_HUMAN | 0.94725208 | 21.2710527 | 3.96369711 | 0.00016879 | 0.01778877 | 0.74044034 |
| HMGN2_HUMAN | 0.94562435 | 21.5156931 | 3.38036852 | 0.00115942 | 0.04309593 | -0.9708787 |
| CAH3_HUMAN | 0.94033407 | 20.9431335 | 3.53655733 | 0.00070442 | 0.03475742 | -0.5308083 |
| ANXA5_HUMAN | 0.91793424 | 26.4886649 | 3.56173472 | 0.00064924 | 0.03350212 | -0.4585756 |
| NEST_HUMAN | 0.9082769 | 26.1175939 | 3.79188323 | 0.00030324 | 0.02405127 | 0.21767321 |
| APOBR_HUMAN | 0.89707291 | 17.8912617 | 4.00761952 | 0.00014498 | 0.0172756 | 0.87641982 |
| HMGB2_HUMAN | 0.88226614 | 22.785919 | 3.58895488 | 0.0005942 | 0.03097821 | -0.3800866 |
| VIME_HUMAN | 0.86811576 | 28.238721 | 3.53502062 | 0.00070793 | 0.03475742 | -0.5352056 |
| FCGRN_HUMAN | 0.78254374 | 16.9813867 | 3.37577779 | 0.00117628 | 0.04309593 | -0.9836002 |
| TAGL2_HUMAN | 0.77067908 | 24.2074544 | 4.04237396 | 0.00012846 | 0.0172756 | 0.98467197 |
| TIF1A_HUMAN | 0.76883831 | 16.1484552 | 3.66665124 | 0.00046045 | 0.02752009 | -0.1538186 |
| PRDX1_HUMAN | 0.76223017 | 26.1219569 | 3.30155746 | 0.00148312 | 0.04707982 | -1.1875475 |
| BCAT1_HUMAN | 0.75805494 | 20.6218819 | 3.98438046 | 0.00015714 | 0.0172756 | 0.80435791 |
| CSRP2_HUMAN | 0.75686511 | 21.496724 | 3.75921994 | 0.00033841 | 0.02480185 | 0.11998663 |
| CD109_HUMAN | 0.75598114 | 20.4885068 | 3.36921769 | 0.00120078 | 0.04309593 | -1.0017577 |
| RET1_HUMAN | 0.69494903 | 21.8238491 | 3.40008201 | 0.00108956 | 0.04309593 | -0.9161097 |
| SERPH_HUMAN | 0.69340911 | 23.3336935 | 4.04575019 | 0.00012695 | 0.0172756 | 0.99521872 |
| HDAC1_HUMAN | 0.68681917 | 19.2204408 | 4.70069696 | 1.175E-05 | 0.00660219 | 3.13530245 |
| CNN3_HUMAN | 0.66993463 | 23.6591625 | 3.7109353 | 0.0003976 | 0.02604994 | -0.0233984 |
| TIF1B_HUMAN | 0.6656231 | 23.8269077 | 5.13270708 | 2.234E-06 | 0.00282434 | 4.63566078 |
| MYT1L_HUMAN | 0.65218036 | 18.0226957 | 3.51918855 | 0.00074505 | 0.03588323 | -0.580432 |
| CBX3_HUMAN | 0.64985238 | 21.449533 | 3.73947409 | 0.00036152 | 0.02508084 | 0.0612018 |
| GGH_HUMAN | 0.6373851 | 21.8362546 | 3.40658951 | 0.00106739 | 0.04283944 | -0.8979804 |
| OS9_HUMAN | 0.63666382 | 18.6086215 | 3.76909439 | 0.00032739 | 0.02434696 | 0.14945992 |
| PLIN2_HUMAN | 0.63642988 | 19.9990879 | 3.53519186 | 0.00070754 | 0.03475742 | -0.5347156 |
| CDN2C_HUMAN | 0.61618131 | 20.6596095 | 3.74212974 | 0.00035832 | 0.02508084 | 0.06909599 |
| CAPG_HUMAN | 0.61527202 | 22.9827687 | 4.0251211 | 0.00013642 | 0.0172756 | 0.93086135 |
| DPYD_HUMAN | 0.60448183 | 19.8681189 | 3.77518924 | 0.00032076 | 0.02434621 | 0.16767713 |
| MYL6_HUMAN | 0.60107514 | 23.8272701 | 4.75864257 | 9.4403E-06 | 0.00660219 | 3.33285765 |
| SSH3_HUMAN | 0.59641405 | 18.3293201 | 4.58318975 | 1.8244E-05 | 0.00922577 | 2.73848465 |
| CALD1_HUMAN | 0.59527134 | 24.13089 | 3.45420665 | 0.00091763 | 0.04035164 | -0.7645776 |
| CALR_HUMAN | 0.58907738 | 25.3808815 | 3.3657268 | 0.00121401 | 0.04309593 | -1.0114097 |
| LYAR_HUMAN | 0.58508342 | 18.4497773 | 3.35912967 | 0.00123939 | 0.04309593 | -1.0296306 |
| IQGA1_HUMAN | 0.58286287 | 24.3358402 | 5.77880009 | 1.6782E-07 | 0.00084865 | 6.98043895 |
| CLIC1_HUMAN | 0.57826631 | 22.6017085 | 3.81110664 | 0.00028421 | 0.02395422 | 0.27542255 |
| PDIA1_HUMAN | 0.57356704 | 25.3963023 | 4.29352938 | 5.2712E-05 | 0.01158972 | 1.78352402 |
| BUD31_HUMAN | 0.56013575 | 19.9975741 | 3.40784165 | 0.00106317 | 0.04283944 | -0.8944893 |
| LDHA_HUMAN | 0.55176289 | 24.9062762 | 3.33532939 | 0.00133518 | 0.04328219 | -1.0951521 |
| PDIA4_HUMAN | 0.55026746 | 24.6702207 | 3.65293176 | 0.00048177 | 0.02752009 | -0.1940106 |
| BIP_HUMAN | 0.53485419 | 26.1082323 | 3.43807889 | 0.00096597 | 0.04091931 | -0.8099072 |
| CHM2A_HUMAN | 0.52042838 | 21.2622534 | 4.46799083 | 2.7936E-05 | 0.00987517 | 2.35461224 |
| SAE1_HUMAN | 0.51780016 | 22.3011829 | 4.41453201 | 3.3981E-05 | 0.00987517 | 2.1782751 |
| XRCC1_HUMAN | 0.5123525 | 19.7197643 | 4.14113743 | 9.0806E-05 | 0.01670467 | 1.29539477 |
| FHL3_HUMAN | 0.49779405 | 19.5591559 | 3.34233822 | 0.00130627 | 0.04328219 | -1.0758918 |
| MYDGF_HUMAN | 0.49723824 | 21.9098406 | 3.47874538 | 0.00084843 | 0.03792056 | -0.6953217 |
| CALU_HUMAN | 0.49130326 | 22.949929 | 3.2974028 | 0.00150235 | 0.04718872 | -1.1988669 |
| ACTN4_HUMAN | 0.4749971 | 25.1522312 | 3.94226342 | 0.00018173 | 0.01875562 | 0.67442368 |
| IKBIP | 0.47028539 | 19.2642561 | 3.28523501 | 0.00156002 | 0.04869774 | -1.2319588 |
| PDCD5_HUMAN | 0.46816866 | 22.771286 | 3.60340041 | 0.00056683 | 0.030173 | -0.3382672 |
| RL21_HUMAN | 0.46149792 | 22.0576487 | 3.34599543 | 0.00129142 | 0.04328219 | -1.0658302 |
| ENPL_HUMAN | 0.45982718 | 25.7250545 | 3.52390453 | 0.00073381 | 0.03568141 | -0.5669749 |
| PYGL_HUMAN | 0.45540281 | 23.1005552 | 3.66668639 | 0.0004604 | 0.02752009 | -0.1537155 |
| NP1L5_HUMAN | 0.45406304 | 20.3570469 | 3.38077286 | 0.00115794 | 0.04309593 | -0.9697576 |
| FBP1L_HUMAN | 0.44790315 | 19.4676984 | 3.53836194 | 0.00070033 | 0.03475742 | -0.5256427 |
| PPCS_HUMAN | 0.44723978 | 20.3816811 | 4.24870022 | 6.1923E-05 | 0.01304777 | 1.63886452 |
| KCC2D_HUMAN | 0.44719734 | 23.7337268 | 3.60546536 | 0.00056301 | 0.030173 | -0.3322799 |
| RAVR1_HUMAN | 0.44485604 | 20.1319361 | 3.92537239 | 0.0001926 | 0.01909739 | 0.62255716 |
| AKT1_HUMAN | 0.44309951 | 20.1685661 | 3.49688416 | 0.00080049 | 0.03713845 | -0.6439081 |
| RUVB1_HUMAN | 0.42680227 | 23.3593965 | 3.99168597 | 0.00015322 | 0.0172756 | 0.82698352 |
| RS7_HUMAN | 0.41978206 | 23.3293253 | 3.33906267 | 0.00131971 | 0.04328219 | -1.0848967 |
| SPSY_HUMAN | 0.41521499 | 21.8500465 | 4.05940495 | 0.00012104 | 0.0172756 | 1.03792879 |
| MYH9_HUMAN | 0.41393475 | 26.3985125 | 4.03343147 | 0.00013253 | 0.0172756 | 0.95676323 |
| EMC8_HUMAN | 0.40896395 | 19.1800036 | 3.64803876 | 0.0004896 | 0.02752009 | -0.2083204 |
| HEBP2_HUMAN | 0.40787965 | 21.7678258 | 3.43383604 | 0.00097909 | 0.04091931 | -0.8218075 |
| PLOD1_HUMAN | 0.40771674 | 20.4296998 | 3.73667155 | 0.00036492 | 0.02508084 | 0.05287499 |
| ANKL2_HUMAN | 0.40699399 | 18.6738296 | 3.39252403 | 0.00111586 | 0.04309593 | -0.9371345 |
| SWP70_HUMAN | 0.40338928 | 21.2333829 | 3.88981443 | 0.00021754 | 0.02053629 | 0.51383173 |
| SCYL2_HUMAN | 0.40073083 | 20.1463363 | 3.773517 | 0.00032256 | 0.02434621 | 0.16267697 |
| CAB45_HUMAN | 0.3913925 | 19.0944362 | 3.3417009 | 0.00130888 | 0.04328219 | -1.0776444 |
| DC1L1_HUMAN | 0.38983263 | 22.6930614 | 3.35831781 | 0.00124255 | 0.04309593 | -1.0318712 |
| GLU2B_HUMAN | 0.36544669 | 24.2684954 | 3.6453442 | 0.00049396 | 0.02752009 | -0.2161952 |
| ANM1_HUMAN | 0.36418271 | 21.3000378 | 3.55597649 | 0.00066149 | 0.03378929 | -0.4751268 |
| RCN2_HUMAN | 0.34724852 | 22.5460243 | 3.47639416 | 0.00085484 | 0.03792056 | -0.7019725 |
| NU153_HUMAN | 0.34375575 | 20.4527691 | 3.35586035 | 0.00125216 | 0.04309593 | -1.0386508 |
| RANB3_HUMAN | 0.3413436 | 19.9071054 | 3.44644481 | 0.0009406 | 0.04077399 | -0.7864121 |
| GT251_HUMAN | 0.33326873 | 19.9241415 | 3.8817868 | 0.00022358 | 0.02053629 | 0.48937273 |
| RHG05_HUMAN | 0.32270503 | 21.3328811 | 3.32993181 | 0.00135786 | 0.0437368 | -1.1099648 |
| MP2K2_HUMAN | 0.29338012 | 21.8538209 | 4.71559586 | 1.1108E-05 | 0.00660219 | 3.18598154 |
| XPO2_HUMAN | 0.28987353 | 23.2260758 | 3.38414356 | 0.00114572 | 0.04309593 | -0.9604083 |
| EP15R_HUMAN | 0.25103878 | 22.707772 | 3.90700929 | 0.00020511 | 0.01994742 | 0.56633007 |
| MACF1_HUMAN | 0.24578482 | 25.232869 | 3.59847537 | 0.00057602 | 0.03034315 | -0.3525379 |
| RHG12_HUMAN | -0.2732411 | 20.0507928 | -3.4283345 | 0.00099634 | 0.04129908 | -0.8372227 |
| PP1R7_HUMAN | -0.3039848 | 23.3778373 | -3.9272681 | 0.00019135 | 0.01909739 | 0.62837109 |
| LRC8A_HUMAN | -0.309764 | 20.7359205 | -3.3846728 | 0.00114381 | 0.04309593 | -0.9589396 |
| RBNS5_HUMAN | -0.310117 | 21.6249464 | -3.6448332 | 0.00049479 | 0.02752009 | -0.2176882 |
| NIT1_HUMAN | -0.3154973 | 21.69254 | -3.3717611 | 0.00119122 | 0.04309593 | -0.994721 |
| ABD12_HUMAN | -0.3186912 | 20.9251531 | -3.8680305 | 0.00023432 | 0.02078852 | 0.44753425 |
| ARFG2_HUMAN | -0.3247478 | 21.0104495 | -3.7506085 | 0.00034831 | 0.02508084 | 0.09432456 |
| AEDO_HUMAN | -0.3292676 | 21.1041364 | -4.079393 | 0.00011286 | 0.0172756 | 1.1006066 |
| ACSF3_HUMAN | -0.32933 | 21.0918497 | -4.1025393 | 0.00010404 | 0.0172756 | 1.17342083 |
| DCMC_HUMAN | -0.337931 | 18.5546184 | -3.3609704 | 0.00123226 | 0.04309593 | -1.0245492 |
| BCS1_HUMAN | -0.3383124 | 20.1610217 | -3.646074 | 0.00049278 | 0.02752009 | -0.2140628 |
| CHMP6_HUMAN | -0.3424074 | 20.4112177 | -3.4396558 | 0.00096114 | 0.04091931 | -0.8054817 |
| ACY1_HUMAN | -0.3595406 | 21.5653263 | -3.3680748 | 0.0012051 | 0.04309593 | -1.0049184 |
| KIME_HUMAN | -0.3659543 | 19.3033579 | -3.3217887 | 0.00139275 | 0.04457674 | -1.1322792 |
| DUT_HUMAN | -0.3739137 | 21.9693001 | -3.3373272 | 0.00132688 | 0.04328219 | -1.0896652 |
| SOGA3_HUMAN | -0.3783262 | 22.5238813 | -3.3603196 | 0.00123478 | 0.04309593 | -1.0263461 |
| GHDC_HUMAN | -0.3819216 | 20.1195833 | -3.9903216 | 0.00015394 | 0.0172756 | 0.82275596 |
| MTX1_HUMAN | -0.3829348 | 21.0312928 | -3.6101584 | 0.00055443 | 0.03014801 | -0.3186638 |
| NRX1A_HUMAN | -0.3844763 | 20.6752336 | -3.707773 | 0.0004018 | 0.02604994 | -0.0327463 |
| CORO7_HUMAN | -0.3938282 | 22.5863661 | -3.6524335 | 0.00048256 | 0.02752009 | -0.1954683 |
| ISCA2_HUMAN | -0.4031595 | 19.1533537 | -3.3535476 | 0.00126126 | 0.04309593 | -1.0450279 |
| PCCB_HUMAN | -0.4120307 | 22.8452437 | -3.659114 | 0.00047205 | 0.02752009 | -0.175912 |
| PCCA_HUMAN | -0.4120574 | 22.6587456 | -3.8241567 | 0.00027195 | 0.02330901 | 0.31473426 |
| RT05_HUMAN | -0.4130393 | 18.954755 | -3.5095606 | 0.00076852 | 0.03598539 | -0.6078665 |
| PYC_HUMAN | -0.4173614 | 24.218858 | -4.0563035 | 0.00012236 | 0.0172756 | 1.0282203 |
| PMGT2_HUMAN | -0.4202209 | 18.436008 | -3.445526 | 0.00094336 | 0.04077399 | -0.7889945 |
| ATAD1_HUMAN | -0.423915 | 21.0865855 | -4.1359307 | 9.2492E-05 | 0.01670467 | 1.2789012 |
| ACBD5_HUMAN | -0.4242016 | 20.8937427 | -3.6893032 | 0.0004272 | 0.02700456 | -0.0872375 |
| DJC11_HUMAN | -0.4258526 | 21.3444493 | -3.4362436 | 0.00097162 | 0.04091931 | -0.815056 |
| PKP4_HUMAN | -0.4293621 | 21.0832093 | -3.7974549 | 0.00029761 | 0.02405127 | 0.23439154 |
| SPRE_HUMAN | -0.4299805 | 20.9482768 | -3.3535703 | 0.00126117 | 0.04309593 | -1.0449653 |
| PITM1_HUMAN | -0.4339686 | 18.7973213 | -3.3414962 | 0.00130972 | 0.04328219 | -1.0782071 |
| CIA2B_HUMAN | -0.4359352 | 19.0717356 | -3.4100037 | 0.00105592 | 0.04283944 | -0.8884589 |
| B3GA3_HUMAN | -0.4423016 | 19.6927656 | -3.7907667 | 0.00030439 | 0.02405127 | 0.21432478 |
| PGRC1_HUMAN | -0.4496297 | 22.2174615 | -3.644571 | 0.00049522 | 0.02752009 | -0.2184541 |
| THTR_HUMAN | -0.4572341 | 21.9781555 | -3.3001578 | 0.00148957 | 0.04707982 | -1.191362 |
| PRDX3_HUMAN | -0.4606269 | 23.492456 | -4.3629359 | 4.1008E-05 | 0.00987517 | 2.00919732 |
| PBLD_HUMAN | -0.4611939 | 19.8165056 | -3.3926222 | 0.00111551 | 0.04309593 | -0.9368616 |
| RM50_HUMAN | -0.4632517 | 17.693039 | -3.5121341 | 0.00076218 | 0.03598539 | -0.6005385 |
| MTCH1_HUMAN | -0.4823735 | 21.7851445 | -3.8775458 | 0.00022684 | 0.02053629 | 0.47646392 |
| SAT2_HUMAN | -0.4824453 | 19.1211098 | -3.8400226 | 0.00025772 | 0.02247019 | 0.36264575 |
| KAD3_HUMAN | -0.4915298 | 23.4672288 | -3.9935363 | 0.00015224 | 0.0172756 | 0.83271831 |
| SFXN4_HUMAN | -0.4984621 | 17.7937926 | -3.6543318 | 0.00047955 | 0.02752009 | -0.1899137 |
| CPT1A_HUMAN | -0.5251009 | 21.6067761 | -4.187222 | 7.7118E-05 | 0.01559941 | 1.44191724 |
| MACD1_HUMAN | -0.5344428 | 20.8478458 | -3.7349571 | 0.00036701 | 0.02508084 | 0.0477832 |
| SYEM_HUMAN | -0.5372474 | 19.9801795 | -3.5160008 | 0.00075275 | 0.03591179 | -0.5895213 |
| PTGR3_HUMAN | -0.5411289 | 20.5047612 | -5.2667423 | 1.3183E-06 | 0.00222221 | 5.11309553 |
| COQ8A_HUMAN | -0.5460879 | 19.8987929 | -3.9917492 | 0.00015318 | 0.0172756 | 0.8271796 |
| HDHD5_HUMAN | -0.5492134 | 20.7548145 | -4.074942 | 0.00011463 | 0.0172756 | 1.0866331 |
| PHIPL_HUMAN | -0.5543722 | 23.9555587 | -3.6345838 | 0.00051175 | 0.02812982 | -0.2476033 |
| FAH2A_HUMAN | -0.5565325 | 18.7584763 | -4.8910342 | 5.6995E-06 | 0.00576444 | 3.78866736 |
| TMLH_HUMAN | -0.5591677 | 20.0617199 | -4.1085356 | 0.00010187 | 0.0172756 | 1.19232473 |
| DHB8_HUMAN | -0.5670146 | 21.0509211 | -3.7223017 | 0.00038283 | 0.0258132 | 0.01024468 |
| RT34_HUMAN | -0.5745226 | 19.5295922 | -3.4125043 | 0.0010476 | 0.04283944 | -0.881481 |
| RT14_HUMAN | -0.5764895 | 17.7859461 | -3.963601 | 0.00016885 | 0.01778877 | 0.74014393 |
| TIM29_HUMAN | -0.5820759 | 18.7282511 | -4.1629315 | 8.4064E-05 | 0.01635035 | 1.36456756 |
| CSPG5_HUMAN | -0.5830398 | 19.2374178 | -3.9970338 | 0.0001504 | 0.0172756 | 0.84356236 |
| MTCH2_HUMAN | -0.5994248 | 22.3281964 | -4.3891461 | 3.7279E-05 | 0.00987517 | 2.09494849 |
| EMC6_HUMAN | -0.6006611 | 17.7372615 | -3.6902766 | 0.00042583 | 0.02700456 | -0.0843703 |
| VAMP3_HUMAN | -0.6060097 | 18.8185677 | -3.3438202 | 0.00130024 | 0.04328219 | -1.0718157 |
| HIP1R_HUMAN | -0.6307167 | 23.0670899 | -3.4901978 | 0.00081786 | 0.03726044 | -0.662882 |
| ABCB6_HUMAN | -0.6397958 | 18.3074337 | -3.3947978 | 0.00110789 | 0.04309593 | -0.9308127 |
| ALDOC_HUMAN | -0.6569845 | 25.8089786 | -3.4823804 | 0.00083861 | 0.03786491 | -0.6850334 |
| GABT_HUMAN | -0.6701549 | 24.889071 | -4.4004816 | 3.577E-05 | 0.00987517 | 2.13212309 |
| MITOK_HUMAN | -0.6762711 | 18.7007069 | -4.4235788 | 3.2876E-05 | 0.00987517 | 2.20803461 |
| SE6L1_HUMAN | -0.6869706 | 20.5832654 | -3.7100142 | 0.00039882 | 0.02604994 | -0.0261219 |
| GBG4_HUMAN | -0.7007514 | 18.9326108 | -3.7778682 | 0.00031788 | 0.02434621 | 0.17569056 |
| TXTP_HUMAN | -0.7277961 | 22.228104 | -4.4922843 | 2.5546E-05 | 0.00987517 | 2.43512783 |
| COX2_HUMAN | -0.7405879 | 22.5522566 | -3.356743 | 0.0012487 | 0.04309593 | -1.0362162 |
| MET15_HUMAN | -0.8277389 | 16.6010139 | -3.4927379 | 0.00081122 | 0.03726044 | -0.6556768 |
| SYVM_HUMAN | -0.8368573 | 17.4619009 | -3.8768073 | 0.00022741 | 0.02053629 | 0.47421707 |
| TM245_HUMAN | -0.8836359 | 20.1065892 | -4.0634403 | 0.00011934 | 0.0172756 | 1.05056746 |
| ZDH14_HUMAN | -1.1110076 | 17.4618704 | -4.4312213 | 3.1969E-05 | 0.00987517 | 2.23320069 |
