## Supplementary Table 6b for "Quantitative proteomic analysis reveals different functional subtypes among IDH-wildtype glioblastoma"

| **Supplementary Table 6b:** Differentially expressed proteins with the highest and lowest log2FC values between GB CL and LGG | | | | | | | | |
| --- | --- | --- | --- | --- | --- | --- | --- | --- |
| **Proteins with highest log2FC values in GB CL (compared to LGG):** | | | | |  |  |  |  |
| **Protein Name** | **Protein** | **Gene** | **Locus** | **Function** | **Pathway** | **Glioma** | **Biological process** | **Molecular function** |
| H13_HUMAN | Histone H1.3 | H1-3 | 6p22 | Forming the macromolecular structure of DNA known as the chromatin fiber. | Cellular responses to stimuli and Programmed Cell Death | none | Negative regulation of transcription by RNA polymerase II, Chromatin organization, Nucleosome assembly | DNA-binding, RNA binding, Structural constituent of chromatin |
| GPNMB_HUMAN | Transmembrane glycoprotein NMB | GPNMB | 7p15 | This transmembrane protein is involved in cell adhesion. | Signaling by PTK6, Signal Transduction. | GPNMB-overexpression is associated with unfavorable prognosis in glioma | Negative regulation of cytokine production, Positive regulation of protein phosphorylation, Cell adhesion, Signal transduction | Integrin binding, Protein binding, Chemoattractant activity |
| LEG3_HUMAN | Galectin-3 | LGALS3 | 14q22 | Galectin-3 is an extracellular matrix protein involved in acute inflammatory responses having chemoattractant activity2. | Gene expression (Transcription), Innate Immune System, RUNX1 regulates transcription of genes involved in differentiation of myeloid cells, Regulation of activated PAK-2p34 by proteasome mediated degradation | LEG3 has been shown as a glioma related marker and expression has been reported to correlate with WHO grade in human gliomas. | Differentiation, Immunity, Innate immunity, mRNA processing, mRNA splicing | IgE binding protein, Protein phosphatase inhibitor activity, RNA binding, Protein binding |
| **Proteins with lowest log2FC values in GB CL (compared to LGG):** | | | | |  |  |  |  |
| **Protein Name** | **Protein** | **Gene** | **Locus** | **Function** | **Pathway** | **Glioma** | **Biological process** | **Molecular function** |
| SYVM_HUMAN | Valine--tRNA ligase, mitochondrial | VARS2 | 6p21 | Catalytic activity, This gene encodes a mitochondrial aminoacyl-tRNA synthetase, which catalyzes the attachment of valine to tRNA(Val) for mitochondrial translation. | tRNA Aminoacylation, Metabolism of proteins, Peptide chain elongation | no results on pubmed | Protein biosynthesis, tRNA metabolic process, Translation, tRNA aminoacylation for protein translation | Aminoacyl-tRNA synthetase, Ligase, Nucleotide binding, Protein binding |
| TM245_HUMAN | Transmembrane protein 245 | TMEM245 | 9q31 | Transmembrane protein, serves as transporter | n/a | no results on pubmed | Biological process | Molecular function |
| ZDH14_HUMAN | Palmitoyltransferase ZDHHC14 | ZDHHC14 | 6q25 | Palmitoyltransferase that could catalyze the addition of palmitate onto various protein substrates. | n/a | no results on pubmed | Protein palmtoylation, Protein targeting to membrane, Peptidyl-L-cysteine S-palmitoylation | Acyltransferase, Transferase, Palmitoyltransferase activity, Protein-cysteine S-palmitoyltransferase activity |
