## Supplementary Table 7a for "Quantitative proteomic analysis reveals different functional subtypes among IDH-wildtype glioblastoma"

| **Supplementary Table 7a:** Differentially expressed proteins between GB MES and LGG | | | | | | |
| --- | --- | --- | --- | --- | --- | --- |
| **Protein** | **logFC** | **AveExpr** | **t** | **P.Value** | **adj.P.Val** | **B** |
| RBP1_HUMAN | 0.92136963 | 21.8238491 | 4.51528031 | 2.3468E-05 | 0.02032125 | 2.47464295 |
| S10AD_HUMAN | 0.91646948 | 22.8090432 | 3.9030064 | 0.00020795 | 0.047799 | 0.5558722 |
| MOES_HUMAN | 0.70063812 | 25.6833345 | 3.93084729 | 0.00018901 | 0.04551536 | 0.63948565 |
| NDUA5_HUMAN | 0.64561424 | 22.3125761 | 4.17307338 | 8.1093E-05 | 0.03417402 | 1.38219143 |
| KCC2D_HUMAN | 0.62695409 | 23.7337268 | 5.06305029 | 2.9321E-06 | 0.01482743 | 4.31369349 |
| NDUA7_HUMAN | 0.59632703 | 22.3277067 | 3.96842628 | 0.00016606 | 0.04551536 | 0.7529319 |
| HEBP1_HUMAN | 0.58674889 | 21.7222764 | 3.84029788 | 0.00025748 | 0.04962936 | 0.36891779 |
| NDUA8_HUMAN | 0.54310068 | 22.680657 | 4.07315915 | 0.00011535 | 0.03888787 | 1.0725933 |
| NP1L5_HUMAN | 0.51379677 | 20.3570469 | 3.83182436 | 0.00026498 | 0.04962936 | 0.34380329 |
| SSH3_HUMAN | 0.51183564 | 18.3293201 | 3.93971564 | 0.00018333 | 0.04551536 | 0.66619758 |
| SYAP1_HUMAN | 0.39990204 | 19.8363071 | 3.94306938 | 0.00018123 | 0.04551536 | 0.67630898 |
| TTC1_HUMAN | 0.39754171 | 22.218239 | 4.29140607 | 5.3116E-05 | 0.02984556 | 1.75449567 |
| GOPC_HUMAN | 0.36372291 | 19.9475772 | 3.86771729 | 0.00023457 | 0.04851979 | 0.45042781 |
| EP15R_HUMAN | 0.2982116 | 22.707772 | 4.64881795 | 1.4278E-05 | 0.01913049 | 2.91341772 |
| RHG12_HUMAN | -0.3170517 | 20.0507928 | -3.9845712 | 0.00015704 | 0.04551536 | 0.80187636 |
| PTGR3_HUMAN | -0.4217634 | 20.5047612 | -4.1117297 | 0.00010073 | 0.03888787 | 1.19158158 |
| ACY1_HUMAN | -0.4348603 | 21.5653263 | -4.0803557 | 0.00011248 | 0.03888787 | 1.09474343 |
| FXR2_HUMAN | -0.4680475 | 21.0251327 | -3.9517147 | 0.00017591 | 0.04551536 | 0.70239899 |
| COQ8A_HUMAN | -0.5306907 | 19.8987929 | -3.8855857 | 0.0002207 | 0.04851979 | 0.50374337 |
| SAT2_HUMAN | -0.5654319 | 19.1211098 | -4.5079639 | 2.4111E-05 | 0.02032125 | 2.45079582 |
| MICA1_HUMAN | -0.5669523 | 20.1785466 | -4.1943968 | 7.5175E-05 | 0.03417402 | 1.44883579 |
| MITOK_HUMAN | -0.582937 | 18.7007069 | -3.8193455 | 0.00027641 | 0.04992118 | 0.30688229 |
| HDHD5_HUMAN | -0.5996702 | 20.7548145 | -4.4566366 | 2.9125E-05 | 0.02104101 | 2.28407931 |
| MACD1_HUMAN | -0.5998509 | 20.8478458 | -4.1989635 | 7.3962E-05 | 0.03417402 | 1.46313447 |
| TMLH_HUMAN | -0.6537092 | 20.0617199 | -4.8110958 | 7.7354E-06 | 0.01913049 | 3.45523368 |
| TXTP_HUMAN | -0.6965424 | 22.228104 | -4.30645 | 5.0313E-05 | 0.02984556 | 1.80225239 |
| GBG4_HUMAN | -0.8580134 | 18.9326108 | -4.6333095 | 1.5132E-05 | 0.01913049 | 2.86212367 |
| MET15_HUMAN | -0.913546 | 16.6010139 | -3.8611568 | 0.00023986 | 0.04851979 | 0.43089167 |
