## Supplementary Table 7b for "Quantitative proteomic analysis reveals different functional subtypes among IDH-wildtype glioblastoma"

| **Supplementary Table 7b:** Differentially expressed proteins with the highest and lowest log2FC values between GB MES and LGG | | | | | | | | |
| --- | --- | --- | --- | --- | --- | --- | --- | --- |
| **Proteins with highest log2FC values in GB MES (compared to LGG):** | | | | | |  |  |  |
| **Protein Name** | **Protein** | **Gene** | **Locus** | **Function** | **Pathway** | **Glioma** | **Biological process** | **Molecular function** |
| RBP1_HUMAN | RalA-binding protein 1 | RALBP1 | 18p11 | Multifunctional protein that functions as a downstream effector of RALA and RALB | Signaling by Rho GTPases, RAC1 GTPase cycle, Doxorubicin Pathway, Pharmacokinetics, GPCR Pathway, ERK Signaling | Expression of RET1 correlated with tumor proliferation. It was shown to be an independent prognostic marker for adverse patient survival in glioma. | Positive regulation of protein phosphorylation, Endocytosis, Chemotaxis, Signal transduction, Small GTPase mediated signal transduction | Nucleotide binding, Protein binding, GTPase activator activity, ATP binding, ABC-type xenobiotic transporter activity |
| S10AD_HUMAN | Protein S100-A13 | S100A13 | 1q21 | S100 proteins are localized in the cytoplasm and/or nucleus of a wide range of cells, and involved in the regulation of a number of cellular processes such as cell cycle progression and differentiation. | n/a | overexpressed in HGG and correlates with tumor grading and microvessel density | Protein transport, Transport, Positive regulation of cytokine production, Positive regulation of cell population proliferation, Positive regulation of interleukin-1 alpha production, Positive regulation of canonical NF-kappaB signal transduction | Copper ion binding, Calcium ion binding, Protein binding, Zinc ion binding, Lipid binding |
| MOES_HUMAN | Moesin | MSN | Xq12 | Moesin is an effector of the immunological synapse. It connects the actin cytoskeleton to the plasma membrane and thereby regulates the structure and function of specific domains of the cell cortex. | Sensory processing of sound, Interleukin-12 family signaling, Nervous system development, Signaling by ALK in cancer, Blood-Brain Barrier and Immune Cell Transmigration: VCAM-1/CD106 Signaling | Moesin seems to be a driver of proliferation and invasion in glioma, especially in glioblastoma | Immunological synapse formation, Cytoskeleton organization, Leukocyte cell-cell adhesion, Host-virus interaction, Regulation of cell shape and size | Double-stranded RNA binding, Actin binding, Signaling receptor binding, Structural constituent of cytoskeleton |
| **Proteins with lowest log2FC values in GB MES (compared to LGG):** | | | | | |  |  |  |
| **Protein Name** | **Protein** | **Gene** | **Locus** | **Function** | **Pathway** | **Glioma** | **Biological process** | **Molecular function** |
| GBG4_HUMAN | Guanine nucleotide-binding protein G(I)/G(S)/G(O) subunit gamma-4 | GNG4 | 1q42 | G proteins are involved as a modulator or transducer in various transmembrane signaling systems | ADORA2B mediated anti-inflammatory cytokines production, Thromboxane signalling through TP receptor. | no results on pubmed | Transducer, Signal transduction, G protein-coupled receptor signaling pathway, Negative regulation of cell growth | Protein binding, G-protein beta-subunit binding |
| MET15_HUMAN | 12S rRNA N4-methylcytidine (m4C) methyltransferase | METTL15 | 11p14 | responsible for the methylation of position C839 in mitochondrial 12S rRNA | n/a | no results on pubmed | Methylation, rRNA base methylation | Methyltranserase, Transferase, Protein binding, rRNA (cytosine-N4-)-methyltransferase activity |
| TXTP_HUMAN | Tricarboxylate transport protein, mitochondrial | SLC25A1 | 22q11 | Mitochondrial electroneutral antiporter that exports citrate from the mitochondria into the cytosol in exchange for malate | Metabolism, Glycolysis, Gluconeogenesis, Fatty acyl-CoA biosynthesis | | Gluconeogenesis, Mitochondrial citrate transmembrane transport, Fatty-acyl-CoA biosynthetic process, Transmembrane transport | Citrate transmembrane transporter activity, Tricarboxylic acid transmembrane transporter activity, Antiporter activity, Citrate secondary active transmembrane transporter activity |
