## Supplementary Table 8a for "Quantitative proteomic analysis reveals different functional subtypes among IDH-wildtype glioblastoma"

| **Supplementary Table 8a:** Differentially expressed protein between IDH HGG and LGG | | | | | | |
| --- | --- | --- | --- | --- | --- | --- |
| **Protein** | **logFC** | **AveExpr** | **t** | **P.Value** | **adj.P.Val** | **B** |
| COG8_HUMAN | -0.5663972 | 19.3777782 | -5.0816215 | 2.7274E-06 | 0.01379265 | 4.13499846 |
