## Supplementary Table 8b for "Quantitative proteomic analysis reveals different functional subtypes among IDH-wildtype glioblastoma"

| **Supplementary Table 8b:** Differentially expressed protein between IDH HGG and LGG | | | | | | |  |  |
| --- | --- | --- | --- | --- | --- | --- | --- | --- |
| **Protein Name** | **Protein** | **Gene** | **Locus** | **Function** | **Pathway** | **Glioma** | **Biological process** | **Molecular function** |
| COG8_HUMAN | Conserved oligomeric Golgi complex subunit 8 | COG8 | 16q22 | Required for normal Golgi function. | Transport to the Golgi and subsequent modification, Vesicle-mediated transport, Metabolism of proteins, Golgi-to-ER retrograde transport, Intra-Golgi traffic | no results on pubmed | Retrograde transport, vesicle recycling within Golgi, Intra-Golgi vesicle-mediated transport, Golgi organization, Protein transport, Glycosylation | Protein binding |
