## Supplementary Table 9 for "Quantitative proteomic analysis reveals different functional subtypes among IDH-wildtype glioblastoma"

**Supplementary Table 9a:** Comparison of DEPs between individual HGG subgroups.

| Comparison | Protein | logFC | AveExpr | t | P.Value | adj.P.Val | B |
| --- | --- | --- | --- | --- | --- | --- | --- |
| IDHHG / GBPN | None |  |  |  |  |  |  |
| IDHHG / GBCL | CLIC1_HUMAN | -0.863 | 22.602 | -4.823 | 0.000007 | 0.028 | 3.484 |
|  | GPNMB_HUMAN | -1.488 | 17.655 | -4.676 | 0.000013 | 0.028 | 2.994 |
|  | PTBP3_HUMAN | -0.630 | 18.550 | -4.533 | 0.000022 | 0.028 | 2.524 |
|  | LEG3_HUMAN | -1.190 | 22.651 | -4.484 | 0.000026 | 0.028 | 2.365 |
|  | IQGA1_HUMAN | -0.532 | 24.336 | -4.475 | 0.000027 | 0.028 | 2.336 |
|  | PDIA1_HUMAN | -0.666 | 25.396 | -4.229 | 0.000067 | 0.039 | 1.552 |
|  | EMIL1_HUMAN | -0.816 | 21.419 | -4.225 | 0.000067 | 0.039 | 1.541 |
|  | GOGA2_HUMAN | -0.355 | 20.344 | -4.197 | 0.000074 | 0.039 | 1.453 |
|  | SERPH_HUMAN | -0.843 | 23.334 | -4.169 | 0.000082 | 0.039 | 1.365 |
|  | BIP_HUMAN | -0.761 | 26.108 | -4.145 | 0.000090 | 0.039 | 1.290 |
|  | ENPL_HUMAN | -0.638 | 25.725 | -4.143 | 0.000090 | 0.039 | 1.285 |
|  | CHM2A_HUMAN | -0.567 | 21.262 | -4.129 | 0.000095 | 0.039 | 1.243 |
|  | PLOD2_HUMAN | -0.820 | 19.624 | -4.116 | 0.000099 | 0.039 | 1.201 |
|  | MOES_HUMAN | -0.858 | 25.683 | -4.073 | 0.000115 | 0.040 | 1.069 |
|  | MYL6_HUMAN | -0.606 | 23.827 | -4.067 | 0.000118 | 0.040 | 1.051 |
|  | PLOD1_HUMAN | -0.511 | 20.430 | -3.968 | 0.000166 | 0.050 | 0.750 |
|  | TIF1B_HUMAN | -0.603 | 23.827 | -3.941 | 0.000183 | 0.050 | 0.668 |
|  | FKB10_HUMAN | -0.672 | 21.083 | -3.930 | 0.000190 | 0.050 | 0.634 |
|  | I2BP1_HUMAN | -0.562 | 19.234 | -3.928 | 0.000191 | 0.050 | 0.628 |
|  | BGH3_HUMAN | -1.079 | 22.763 | -3.920 | 0.000196 | 0.050 | 0.605 |
|  | PLGT3_HUMAN | -0.579 | 19.073 | -3.901 | 0.000209 | 0.050 | 0.548 |
|  | TAGL2_HUMAN | -0.875 | 24.207 | -3.892 | 0.000216 | 0.050 | 0.522 |
| IDHHG / GBMES | None |  |  |  |  |  |  |
| GBPN / GBCL | None |  |  |  |  |  |  |
| GBPN / GBMES | None |  |  |  |  |  |  |
| GBCL / GBMES | None |  |  |  |  |  |  |

**Supplementary Table 9b:** Comparison of DEPs between each HGG subgroup and the three other pooled HGG subgroups. Note: there was only one protein significantly overexpressed in GB CL (compared to the pooled HGG).

| Comparison | Protein | logFC | AveExpr | t | P.Value | adj.P.Val | B |
| --- | --- | --- | --- | --- | --- | --- | --- |
| IDHHG / High grade Glioma | None |  |  |  |  |  |  |
| GBPN / High grade Glioma | None |  |  |  |  |  |  |
| GBCL / High grade Glioma | IQGA1_HUMAN | 0.407 | 24.336 | 4.950 | 0.000005 | 0.023 | 3.614 |
| GBMES / High grade Glioma | None |  |  |  |  |  |  |
