## Supplementary Table 10 for "Quantitative proteomic analysis reveals different functional subtypes among IDH-wildtype glioblastoma"

Supplementary Table 10: Univariable Cox Regression Analysis of Differentially Expressed Proteins and Survival

| **DEP** | **HR (95% CI)** | **P-value** |
| --- | --- | --- |
| COG8 | 1 (0.49-2) | 0.99 |
| RBP1 | 0.58 (0.17-2) | 0.4 |
| CENPV | 0.9 (0.6-1.3) | 0.59 |
| TYB10 | 2.8 (1.8-4.3) | <0.001 |
| SE6L1 | 0.26 (0.14-0.47) | <0.001 |
| AIG1 | 0.56 (0.39-0.8) | 0.002 |
| REN3A | 0.66 (0.51-0.86) | 0.002 |
| H13 | 1.8 (1.3-2.4) | <0.001 |
| GPNMB | 2.4 (1.6-3.4) | <0.001 |
| LEG3 | 2.9 (2-4.1) | <0.001 |
| SYVM | 0.35 (0.18-0.66) | 0.001 |
| TM245 | 0.24 (0.12-0.49) | <0.001 |
| ZDH14 | 0.61 (0.36-1.1) | 0.075 |
| S10AD | 2.3 (1.5-3.6) | <0.001 |
| MOES | 3.1 (2.1-4.7) | <0.001 |
| TXTP | 0.21 (0.11-0.43) | <0.001 |
| GBG4 | 0.38 (0.22-0.65) | <0.001 |
| MET15 | 0.44 (0.24-0.81) | 0.008 |
